# Winter forecasting of respiratory viruses in Victoria Australia

**DOI:** 10.64898/2026.05.18.26353544

**Authors:** Alec S. Henderson, Robert Moss, Adeshina I. Adekunle, Henry Ye, Mitchell O’Hara-Wild, Oliver Eales, Katharine L. Senior, Ruarai Tobin, Saras M. Windecker, Nick Golding, Ellie Robinson, Janet Strachan, Rob J. Hyndman, Peter Dawson, James M. McCaw, Emma S. McBryde, Freya M. Shearer

**Affiliations:** Frazer Institute, Faculty of Health, Medicine and Behavioural Sciences, University of Queensland; Infectious Disease Dynamics Unit, Centre for Epidemiology and Biostatistics, Melbourne School of Population and Global Health, The University of Melbourne, Australia; Defence Science and Technology Group, Melbourne, Victoria, Australia; Department of Econometrics and Business Statistics, Monash University; School of Mathematics and Statistics, The University of Melbourne, Australia; Infectious Disease Ecology and Modelling Team, The Kids Research Institute Australia, Australia; School of Physics, Mathematics and Computing, University of Western Australia, Perth, Australia; Community and Public Health, Department of Health, Victoria, Australia; Australian Institute of Tropical Health and Medicine, James Cook University, Townsville, Queensland; Burnet Institute, Australia

## Abstract

Temperate regions of the world, such as southern Australia, often experience increased health burden from respiratory pathogens during winter. The ability to forecast short-term trends in cases of these pathogens is of significant interest to public health. Across the 2024 southern hemisphere winter period, the Australia–Aotearoa Consortium for Epidemic Forecasting and Analytics (ACEFA) ran a pilot respiratory virus forecasting initiative in collaboration with the Victorian Department of Health. Each week from the 9th of May 2024 through to 12th September 2024, the consortium solicited 28-day forecasts of daily case incidence for influenza, severe acute respiratory syndrome coronavirus 2 (SARS-CoV-2), and respiratory syncytial virus (RSV) from multiple research groups. Four component model forecasts were contributed by three different research groups, with a fourth group utilising the component forecasts to generate ensemble forecasts (making a total of six models, four component models and two ensembles). Here we statistically evaluated the performance of each forecast and a baseline model against the observed case data. The two ensemble models were found to be frequently the top performing models. All models performed worse than the baseline model around the epidemic peaks for each pathogen.

## 1 Introduction

Influenza, respiratory syncytial virus (RSV) and COVID-19, are major respiratory diseases which are monitored together in Australia [1], as advocated by the World Health Organization (WHO) [2], and cause substantial annual health burden worldwide [3]. The magnitude and timing of respiratory virus epidemics are highly variable and difficult to predict. In temperate winter seasons, they can place significant demands on hospital services, particularly if multiple epidemic peaks occur simultaneously [4].

Accurate and timely forecasts of infectious disease activity have the potential to enable a rapid understanding of the status of concurrent epidemics and inform decisions about hospital staffing, resource allocation, and public health messaging [5, 6, 7]. As part of efforts worldwide to integrate forecasting into public health outbreak response, large-scale collaborations have been used in forecasting applications to generate multi-model ensembles with component models solicited from multiple research teams [8, 9]. Multi-model ensembles, i.e., models that combine predictions from multiple different component models, have consistently been shown to outperform any single model [10, 11] and in well-established forecasting domains, such as weather forecasting, are routine practice [12, 13]. They have been used successfully during epidemics of many diseases, including influenza [14], COVID-19 [8, 9], Ebola [15], and dengue [16].

Infectious disease researchers and public health agencies are increasingly using so-called “forecasting hubs” to coordinate collaborative efforts, with prominent examples in Europe (e.g., through “RespiCast” of the European Centre for Disease Control [8]) and the United States (e.g., through the Center for Forecasting and Outbreak Analytics of the Centers for Disease Control and Prevention [9]). In Australia, real-time forecasts of influenza cases based on a single forecasting model were generated in partnership with public health agencies over 2014–19 [17]. Building on those efforts, the Australian Government funded a national consortium [18] to deliver multi-model ensemble forecasts of COVID-19 cases to national decision-making committees every week from July 2020 (with single model forecasts from April) to December 2023 [19, 20, 21].

By late 2023, following the cessation of emergency surveillance initiatives, members of what is now the Australia–Aotearoa Consortium for Epidemic Forecasting and Analytics (ACEFA) sought to develop a platform in Australia for routine, collaborative forecasting of high-burden respiratory viruses and for advancing infectious disease forecasting science more generally. In 2024 ACEFA launched the Australia–Aotearoa Forecasting Hub, drawing on opensource software tools from the hubverse [22] to facilitate efficient coordination between teams and rapid turnaround from data receipt to validated outputs. The ACEFA Forecasting Hub ran a pilot during the Southern Hemisphere winter season of 2024, soliciting weekly forecasts from multiple teams of four week-ahead incident cases for three high-burden respiratory viruses — influenza, SARS-CoV-2, and RSV — for the state of Victoria. Three teams from three different research institutions contributed four component model forecasts, with a fourth team (from another institution) generating ensemble forecasts and conducting performance evaluation. While respiratory virus activity began to increase in May 2024, due to various challenges establishing new workflows (relative to the previous COVID-19 forecasting activities) including, accommodating additional pathogens (influenza and RSV), new research groups, and non-emergency research ethics and data sharing agreements, the Hub only reported prospective “real-time” forecasts to the Victorian Department of Health on a weekly basis from 15th August 2024.

In this paper, we conduct a detailed performance analysis of weekly ensemble forecasts and component forecasts submitted by each team during the 2024 Hub pilot. Our analysis includes determining when during the season a component or ensemble model performed better/worse than a baseline model, and assessments of forecast bias and interval coverage. We use a mix of prospectively and retrospectively generated forecasts. Retrospective forecasts were used up until the 8th of August and prospective forecasts thereafter, covering the respiratory virus season which we defined as being from the 9th May to 12th September 2024. Our retrospective forecasts were generated only on data that would have reasonably been available on the forecast date (i.e. an approximation of a realtime forecast). The insights gained from our analysis informed the design of our winter forecasting program in 2025.

## 2 Data and methods

### 2.1 Data

Modelling teams were provided with target data of daily aggregate counts of laboratory-confirmed case notifications (PCR only) for each of influenza, RSV and SARS-CoV-2 for the south-eastern Australian state of Victoria (population of approximately 7 million people in 2024). De-identified data (date of notifi-cation and organism detected) were extracted from the following Department of Health Victoria notification databases [23]: Public Health Event Surveillance System (PHESS) for influenza and RSV; and the Transmission and Response Epidemiology Victoria (TREVi) for COVID-19. Only data from electronic laboratory results (ELR) were included in analyses due to the delay in manual entry of non-ELR records. In 2024, all COVID-19 notifications were by ELR and approximately 85% of influenza and RSV notifications. The time-series of daily confirmed case counts extended from 2nd January 2015 for influenza, 16th January 2022 for RSV (RSV first became notifiable in Victoria in February 2022) and 25th January 2020 for SARS-CoV-2 up to 17th September 2024. Forecasting teams generated and submitted 28-day forecast trajectories of daily case counts. Teams were permitted to utilise other data sets as they deemed appropriate (for retrospective forecasts this excluded data which would not have reasonably been available at the forecast date). A plot of the daily case notifications for each of the three pathogens is displayed in Figure 1.

**Figure 1:**
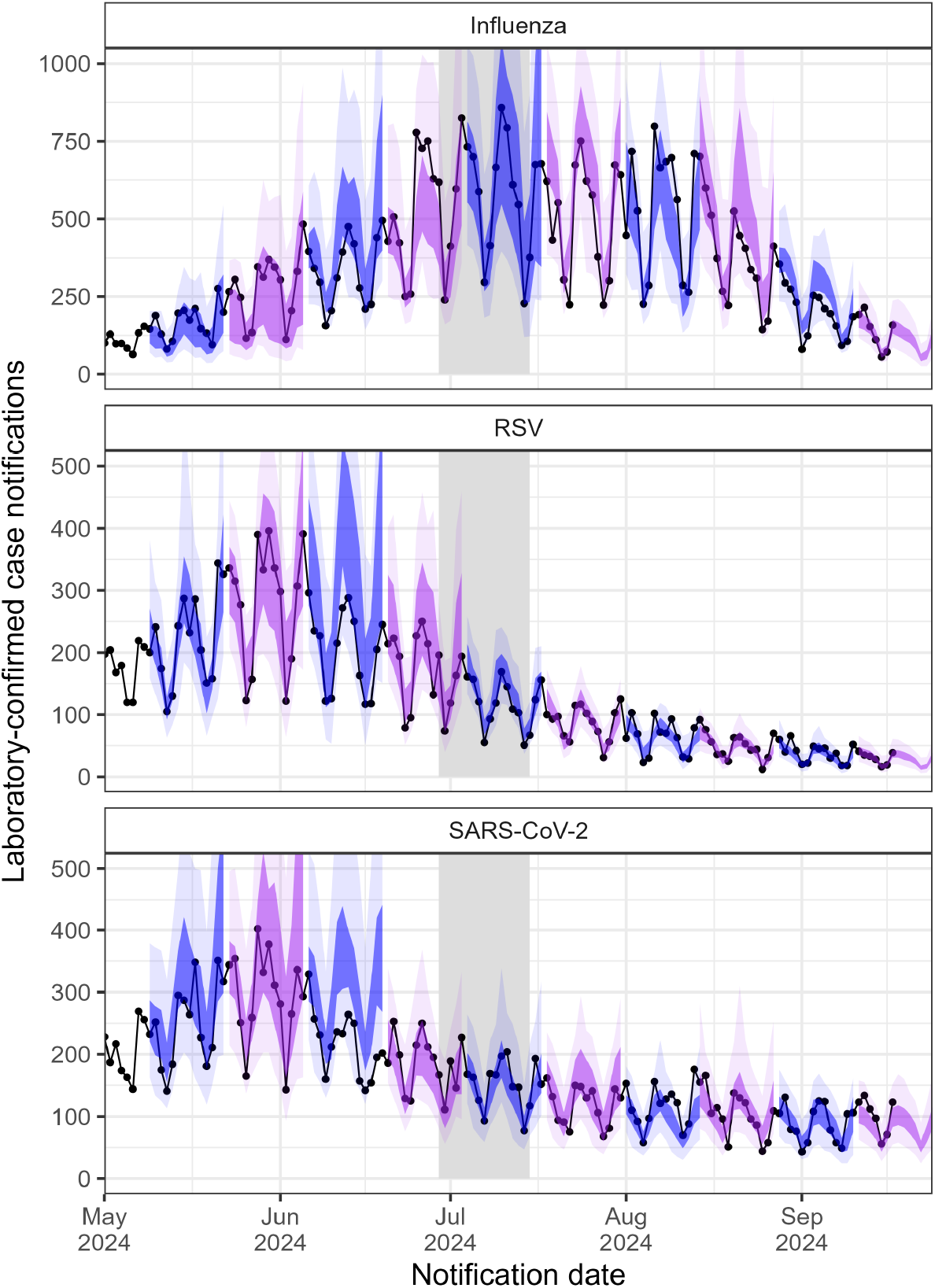
Half of the two-week ahead forecasts from the ensemble mixture model for influenza, RSV, and SARS-CoV-2 in the Australian state of Victoria in 2024. The lightest shading represents the 95% prediction interval and the darker the 50% prediction interval. Alternating colours are used to visualise forecasts from a new forecast date. Daily confirmed case notifications are overlaid (black dots and line) as reported at the end of the study period. The vertical grey shading represents the winter school holidays. Four-week ahead forecasts for each component and ensemble model are provided in the supplementary material.

When referring to the peak of an epidemic we have taken this to be the date at which the seven day sum of cases was largest (i.e. the date with the most case notifications in the weekly notification series).

### 2.2 Component models

Each week during the season (or retrospectively), four component models (from three teams) produced 28-day ahead forecasts of daily reported case incidence. These models were used as component models to form two different ensembles: one from averaging probability densities (linear pooling [24]) and the other from averaging quantiles (quantile averaging [24]). We compared each of these component and ensemble models against a baseline model. A summary of the models is presented in Table 1. Forecasts were submitted in the form of 2000 simulated case trajectories over the forecast horizon. This allowed the set of trajectories to form a predictive distribution at each date over the forecast horizon which could then be assessed following the observation of the number of notified cases (ground truth). Using half of the two-week ahead forecasts as an example, as shown in Figure 1, each day produces a set of prediction intervals which can then be scored (showing here intervals for two-week ahead predictions). For the evaluation analysis, we used weekly counts of the target data and forecast trajectories, i.e. by summing the trajectories over seven day periods beginning from the forecast date (corresponding to one-week, two-week, three-week and four-week ahead forecast horizons).

**Table 1:**
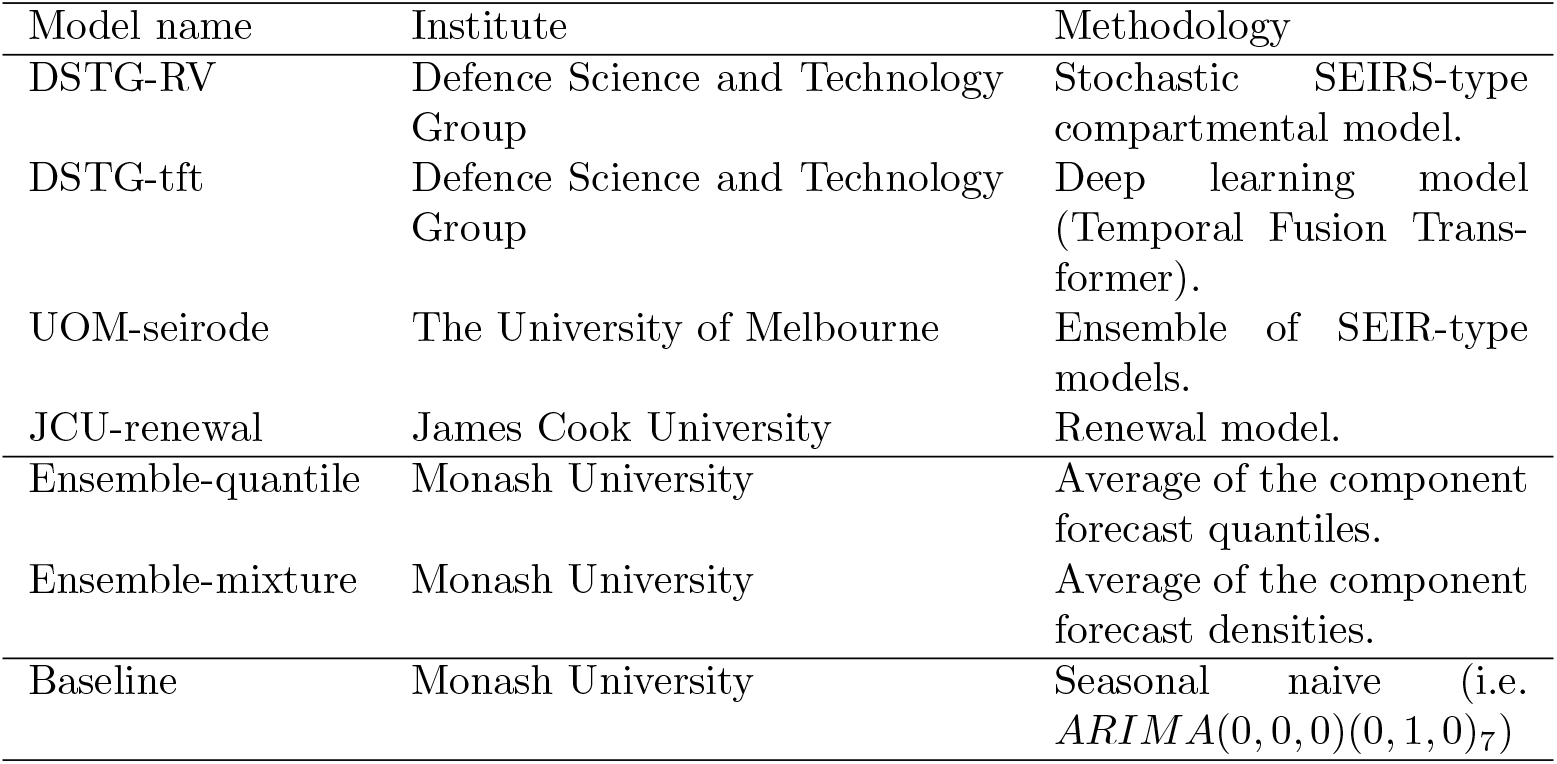
Overview of the different component, ensemble, and baseline models used in this study.

### 2.3 Ensemble models

The distributional mixture ensemble model (also known as linear pooling) is a weighted average of the component forecast densities, which produces an ensemble forecast that closely resembles the shape of the underlying component forecasts (including wider tails/extremes and potentially multimodal forecast distributions when component forecasts differ). The quantile ensemble model is a weighted average of the component forecast quantiles, producing a smoother (often unimodal) distribution with narrower tails/extremes even when component forecasts differ.

### 2.4 Scoring forecasts

To compare and contrast the different forecasts we performed formal forecast evaluations. We evaluated performance (segregated by pathogen) by forecast date, forecast horizon, and overall performance. Evaluations by forecast date provide insight into when during the season (e.g., before the peak, after the peak, or around the peak) models performed better/worse. However, since evaluating by forecast date means grouping forecasts of different horizon length, the resulting score may emphasise a different horizon depending on the epidemic phase of the forecast. That is, if target data values are orders of magnitude different across the forecasting period (e.g. due to exponential growth), and the performance metric used is scale dependant, it may place different emphasis on performance across epidemic phases. For example, during the increasing phase of an epidemic, the emphasis of overall performance may be on later forecasts (i.e. they have larger values which may dominate the scoring). Similarly, in the decreasing phase, the emphasis may be on earlier forecasts.

Evaluations by forecast horizon provide an indication of the reliability of models as they predict observations that fall at different times (i.e. horizons) into the future. However, this does not evaluate where in the epidemic phase that each horizon is most accurate (an idea of which is given through evaluations by forecast date). Hence we evaluated forecast performance across all forecast dates at one-week, two-week, three-week, and four-week horizons.

Finally, we evaluate overall performance across the entire season and all horizons. Together, our assessments provide an understanding of the absolute performance and strengths and weaknesses of each model.

#### 2.4.1 Scoring rules

The continuous ranked probability score (CRPS) was the primary score that we used to assess forecast accuracy. The CRPS is commonly used in infectious disease forecasting [25] (and many other forecasting domains) and is defined as [26]:

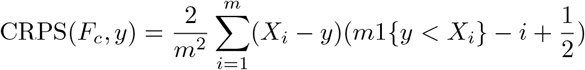

where *X*_*i*_ is the sorted samples from the predictive distribution, *m* the number of samples from the predictive distribution *F, F*_*c*_ the cumulative distribution of *F*, and *y* the observed count (notified cases). One of the advantages of the CRPS is that it is a proper scoring rule [27] so teams are incentivised to produce “honest” probabilistic forecasts (i.e. appropriately capturing uncertainty).

In our primary analysis, we applied CRPS to both untransformed daily case counts and log-transformed counts. Untransformed CRPS is scale-dependent and can be interpreted as a measure of absolute error whereas log-transformed CRPS is a measure of relative error and less dependent on the order of magnitude of the predicted quantity (i.e. is scale invariant). Bosse and colleagues [28] recently argued that log-transformed CRPS better accommodates the exponential nature of infectious disease spread, avoiding issues with forecasting scores being strongly influenced by the order of magnitude of the forecast quantity. For example, CRPS will typically assign higher (worse) scores to forecast targets with higher expected values (e.g. around the peak of an epidemic). The advantages and disadvantages of scale dependent and invariant forecast metrics for public health applications are discussed in depth by Bosse and colleagues [28] and Bracher and colleagues [25].

Other scoring rules have been popular amongst infectious disease forecasters due to the significance that different metrics place on the probability distribution [25], including the log score [29], and less frequently, the Dawid-Sebastiani score (DSS) [30]. The log score and DSS results are provided in the supplementary material, along with full definitions of all scoring rules. When choosing a scoring rule, the utilisation of the forecasts must be considered. In this analysis, we have not taken a specific end user question into account—while the paper is motivated by the public health utility of the forecasts, we are not addressing a specific public health question. Therefore, we have provided multiple proper forecasting scores as suggested in [28, 31, 32]. The log score and DSS scores are sensitive to outliers. For example, where the data fall outside the prediction interval, the log score will tend to infinity, and so we clipped log scores [29, 25] at a maximum value of 20. This property of the log score may be useful in some contexts but not others; as discussed in [25], the log score will be optimal when the predicted value is at the mode of the forecast, the CRPS will be optimal when it is at the median (a generalization of the mean absolute error) and the DSS when it is at the mean. Deciding which of these to emphasise or to use a different scoring rule all together, depends on the application.

#### 2.4.2 Baseline model

Forecasts were compared against a simple “baseline” model, here taken to be the seasonal naive model (as implemented in the *fable* package [33]). This model predicts that the next data point will be the same as its value in the previous season (or periodic cycle), here taken to be the value of the data one week prior to the forecast origin. Specifically, the baseline model is assumed to be ARIMA(0,0,0)(0,1,0)m where m (here it is 7 days) is the seasonal period. This can be expressed as: *Y*_*t*_ = *Y*_*t−m*_ + *Z*_*t*_ where *Z*_*t*_ is a normal independent and identically distributed (iid) error. As with all other models, daily predictions were made and then summed over seven days for evaluation purposes.

#### 2.4.3 Forecast skill score

To measure any gain in forecast performance compared to the baseline model described above, we utilised the skill score:

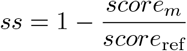

where the score is either the CRPS or log-transformed CRPS, *m* refers to the forecast model, and ref is the baseline model.

#### 2.4.4 Forecast coverage

To assess how well the forecast prediction intervals quantified the uncertainty in forecasts, we calculated forecast coverage. Coverage is the probability that a forecast credible interval will contain the observed value, and for a perfectly calibrated forecast each X% credible interval will include the observed value X% of the time.

#### 2.4.5 Forecast bias

To understand if a forecast tended to over-predict (above the observed counts) or under-predict (below the observed counts), we computed bias:

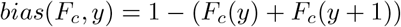

with the bias taking values between −1 and 1, inclusive, and positive values indicating over-prediction and negative values indicating under-prediction. Over/under prediction is computed as:

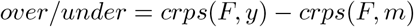

If *m* is greater than *y* then its over and if *m* is less than *y* then under with the other being 0. If they are the same then the forecast is unbiased. With the component *crps*(*F, m*) being called the dispersion.

## 3 Results

Half of the two-week ahead ensemble-mixture forecasts for influenza, RSV, and SARS-CoV-2 in the Australian state of Victoria from 9th May to 12th September 2024 are shown in Figure 1 as example forecasts, overlaid by the case notification data as reported at the end of the study period.

The RSV epidemic peaked in late May. SARS-CoV-2 also peaked in late May, with two short periods of resurgence around early August and mid-September, coinciding with the emergence of new variants (for example, JN.1-descendant lineages with the F456L spike mutation in April/May and the KP.3 descendant lineages or variants with the S31 deletion in August/September). Influenza peaked in early July, although the decline was short-lived, with a resurgence and second (lower) peak occurring in early August. The transient decline in influenza activity coincided with the timing of winter school holidays.

Our ensemble forecasts were generally in good agreement with the observed data, as demonstrated by the example forecasts. In the below sections, we formally examine forecast performance by forecast date (3.1), then horizon (3.2) and finally, overall (3.3). Additional results are included in the supplementary material, including plots of four-week ahead forecasts from each component and ensemble model by pathogen (overlaid with daily case notifications).

### 3.1 Forecast date

Skill scores of each model, relative to the baseline model, varied across models and by forecast date and pathogen, although some clear patterns emerged (Figure 2). Firstly, no model performed consistently better or worse than baseline across all forecast dates and pathogens. The ensemble models (both quantile and mixture) outperformed the component models for all three pathogens, but only marginally compared to the DSTG-tft model for SARS-CoV-2.

**Figure 2:**
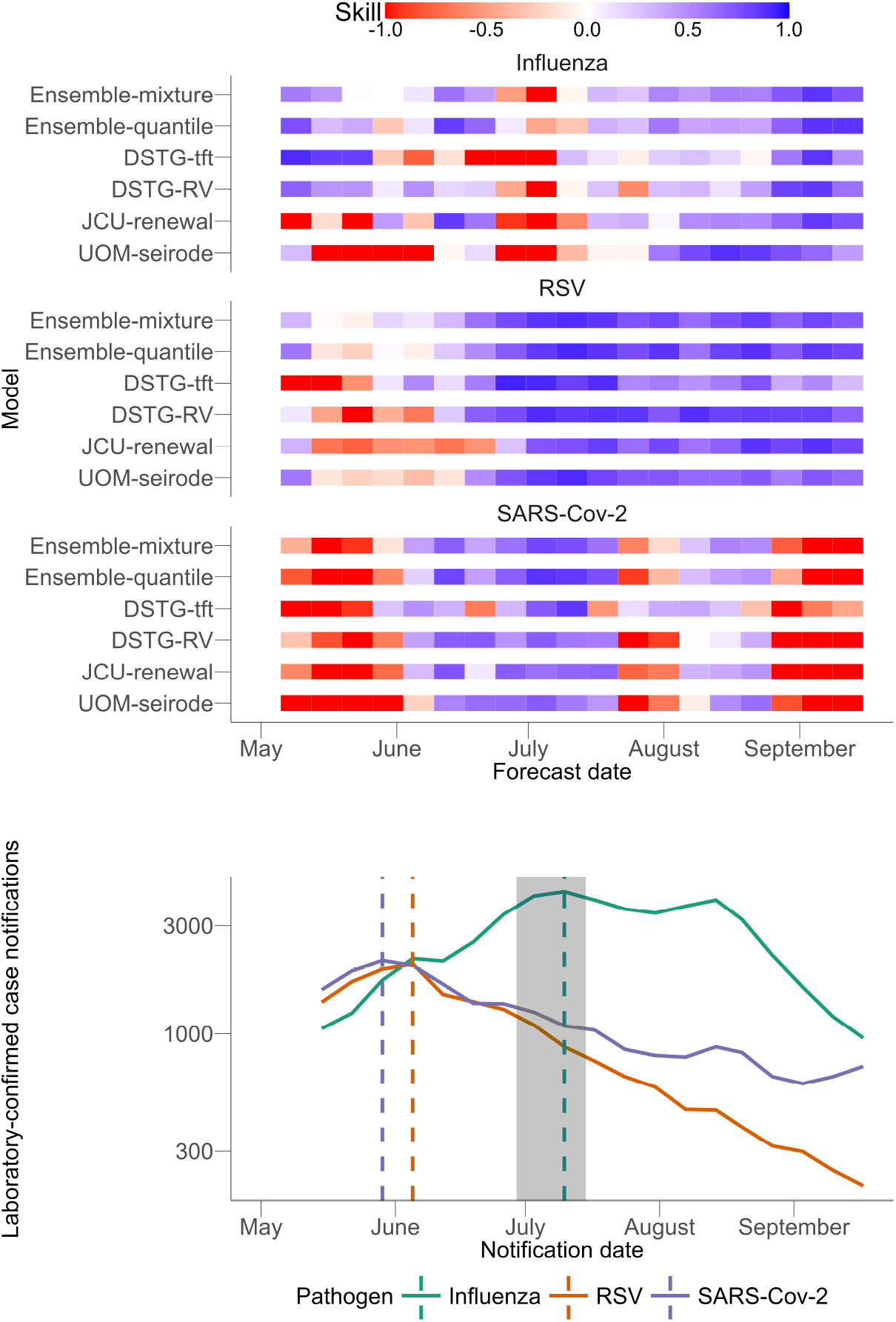
Top: Skill score (for log-transformed CRPS) comparison of each component and ensemble model relative to the baseline model by forecast date. Bottom: Weekly case notifications of each pathogen with the dashed vertical lines indicating the peak timings and the grey shaded area indicating the Victorian school holidays.

Model performance varied substantially by epidemic phase. Before the epidemic peak of influenza, performance across models was mixed. The ensemble models, DSTG-tft model, and DSTG-RV model, performed consistently better than the baseline model, while the UOM-seirode model and JCU-renewal model were worse. The UOM-seirode model tended to over predict the rate of epidemic growth preceding the peak and the magnitude of the observed peak (Table 2). The JCU-renewal model’s performance was mixed during these periods, with both under and over predictions (Figure 3).

**Table 2:** Bias (between −1 and 1 inclusive) of the component models across forecasts in which the observed peak fell within the 28 day forecasting horizon.

| Model | Influenza | RSV | SARS-CoV-2 |
| --- | --- | --- | --- |
| JCU-renewal | 0.65 | 0.92 | 0.75 |
| UOM-seirode | 0.98 | 0.79 | 1.00 |
| DSTG-RV | 0.22 | 0.87 | 0.73 |
| DSTG-tft | $-1.00$ | $-1.00$ | $-0.99$ |

**Figure 3:**
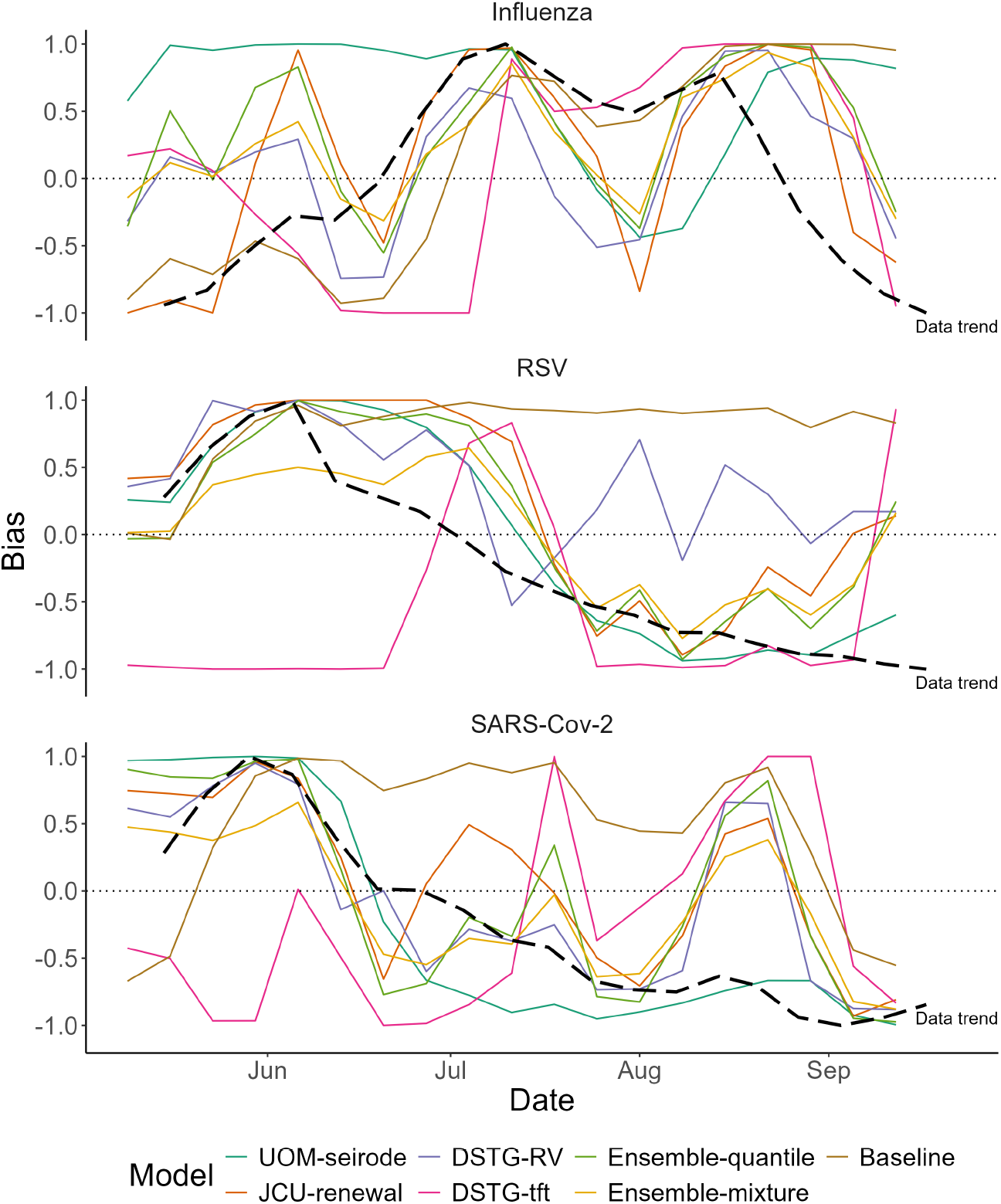
Forecast bias calculated for each component model by forecast date and pathogen (coloured lines) with the black dashed line showing a scaled version of the weekly case counts (Data trend). The horizontal dotted line indicates zero bias (positive values indicate over-prediction, negative values indicate under-prediction).

Around the epidemic peaks of each pathogen, all models performed worse than the baseline model. The JCU-renewal model, UOM-seirode model, and DSTG-RV model all had positive biases indicating that they overshot the height of the epidemic peaks (see Table 2 for biases), whereas the DSTG-tft model had a negative bias indicating that it undershot the peaks.

Post-peak for RSV, all component and ensemble models consistently performed better than the baseline model. For influenza post-peak, the ensemble models consistently outperformed the baseline model, with component models also frequently outperforming the baseline model. Results for SARS-CoV-2 post-peak were mixed (see supplement for metrics), with models performing poorly around the short periods of resurgence in early August and mid-September. The DSTG-RV model and JCU-renewal model initially under-predicted (at the onset of the resurgence) and then over-predicted (as the resurgence subsided), whereas the UOM-seirode model under-predicted during this entire phase. These results reflect a general challenge for the models to predict change points in the observed data.

### 3.2 Forecast horizon

When comparing values of CRPS and log-transformed CRPS, averaged across the season (i.e. averaged over all forecast dates), forecast performance generally decreased over the target horizon (Figure 4). As expected, observations were harder to predict the further they fell into the future. Forecast performance was best for the one-week horizon and degraded as the horizon increased, with performance poorest for the four-week horizon. This was true across across all pathogens and models, except the DSTG-tft model which exhibited no performance degradation over the two-, three-, and four-week horizons for RSV and SARS-CoV-2, and was frequently the best performing model for the four-week horizon.

**Figure 4:**
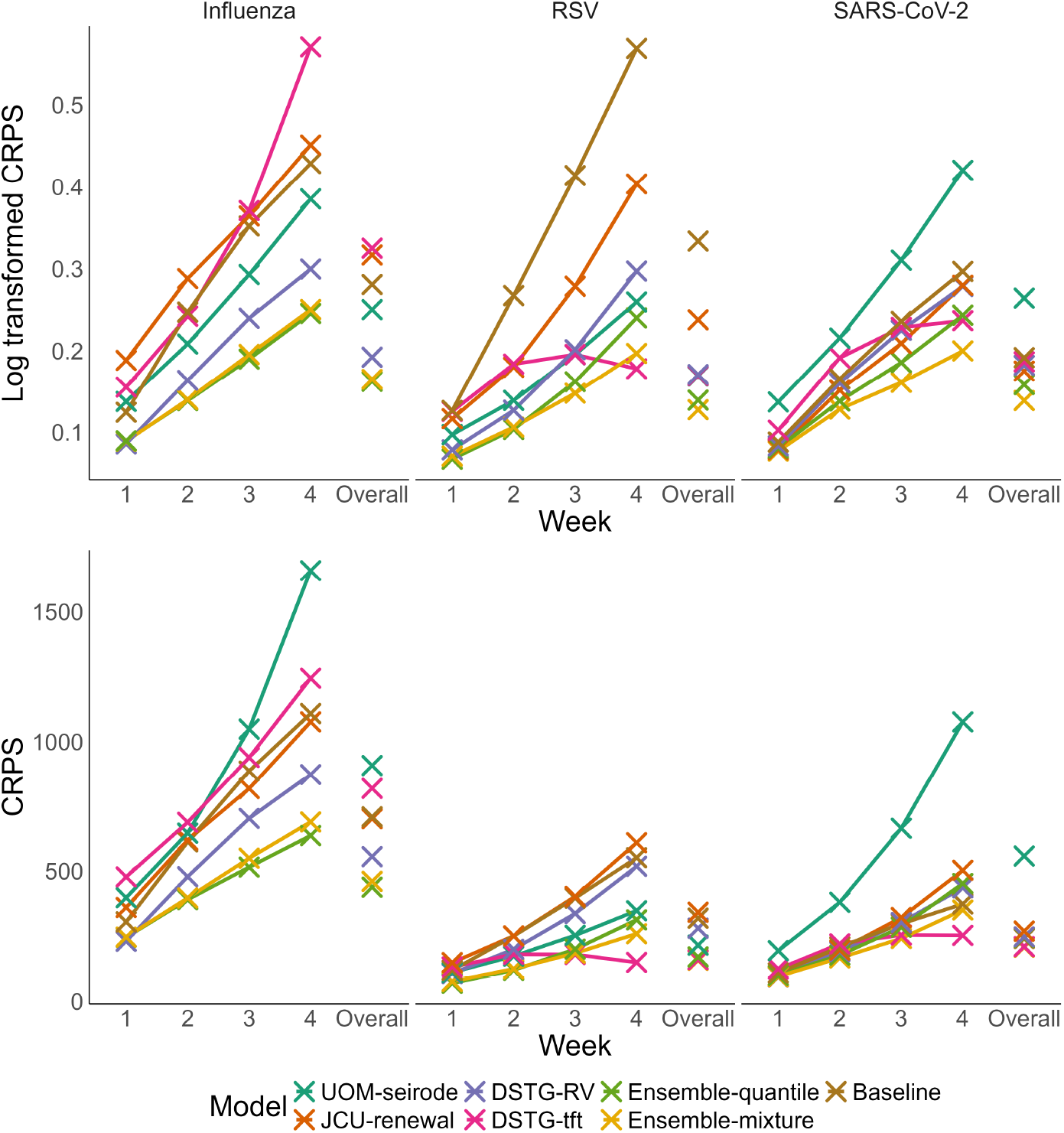
Continuous ranked probability scores (CRPS) and log-transformed CRPS by forecast horizon and overall for all pathogens.

Figure 4 gives a graphical representation of the results for both CRPS and log-transformed CRPS (other metrics are displayed in a tabular format in the supplement). No single component model dominated the top ranking across all horizons and pathogens. For log-transformed CRPS, the ensemble models were consistently high-performing models, with both the quantile-ensemble model and mixture-ensemble model ranking within the top three across all horizons and pathogens. The baseline model was not consistently the lowest-ranking model, but it always ranked in the bottom four (of seven).

The ensemble models did not exhibit a consistent performance ranking (relative to each other) based on forecast horizon. The ensemble-quantile model consistently (for both log-transformed CRPS and CRPS) performed worse than the ensemble mixture for SARS-CoV-2. For influenza, the ensemble-quantile model was better for all but the one-week horizon for log-scaled CRPS (for CRPS the ensemble quantile was better). For RSV, performance was horizon dependant with the ensemble-quantile model outperforming the ensemble-mixture model for the one- and two-week horizons, whereas the ensemble-mixture model out-performed the ensemble-quantile model for the three- and four-week horizons.

Amongst the component forecasts, the DSTG-RV model was the top performing component across all horizons for influenza, and outperformed the ensembles for the one-week horizon. For RSV, the component models exhibited inconsistent performance rankings across horizons. The DSTG-RV model was the top performing component under log-transformed CRPS for the one- and two-week horizons (both CRPS and log-transformed for one-week). The UOM-seirode model was the top performing component model for the two-week horizon for CRPS and three-week horizon for log-tranformed CRPS. Finally, the DSTG-tft model was the top performing component model for the four-week horizon (both CRPS and log-transformed CRPS), and outperformed both ensemble models.

As with RSV, component model performance rankings for SARS-CoV-2 varied by horizon. The JCU-renewal model was the top performing for one-, two- and three-week horizons under log-transformed CRPS. However, for all horizons the JCU-renewal model was never the top ranked component model under CRPS. The DSTG-tft model was the top ranked component model for the four-week horizon for both CRPS and log-transformed CRPS, and the three-week horizon for CRPS. The DSTG-RV model was the top ranked component model for one- and two-week horizons under CRPS.

### 3.3 Overall

When averaging log-transformed CRPS across horizons and forecast dates, the best performing models across all three pathogens were the two ensembles (see Figure 4). The top performing component model varied by pathogen with DSTG-RV top-ranked for influenza, DSTG-tft for RSV, and JCU-renewal for SARS-CoV-2.

When averaging CRPS across horizons and forecast dates, one of the ensembles was always the top performing model (see Figure 4). For RSV and SARS-CoV-2, the second ranked model (best component model) was the DSTG-tft model. For influenza, the DSTG-RV model was the best component model (third overall). It is note worthy that the model performance rankings sometimes changed substantially between log-transformed CRPS and CRPS. For example, the JCU-renewal model moved from the best component model (third overall) for SARS-CoV-2 when using log-transformed CRPS to second last (sixth overall) under CRPS.

We found marked differences in interval coverage across models (Figure 5). The best interval coverage was demonstrated by DSTG-RV for all three pathogens (tracking closely to the line indicating perfect calibration). DSTG-tft and UOM-seirode were both highly overconfident, with many observations falling outside 100% intervals. In general, models exhibited a tendency to be over-confident rather than underconfident. A clear difference was observed between the ensemble-quantile and the ensemble-mixture. The ensemble-mixture exhibited a tendency to be slightly overconfident whereas the ensemble-quantile was overly uncertain. Interval coverage of the ensemble-mixture was comparable to the component model with the best coverage (DSTG-RV).

**Figure 5:**
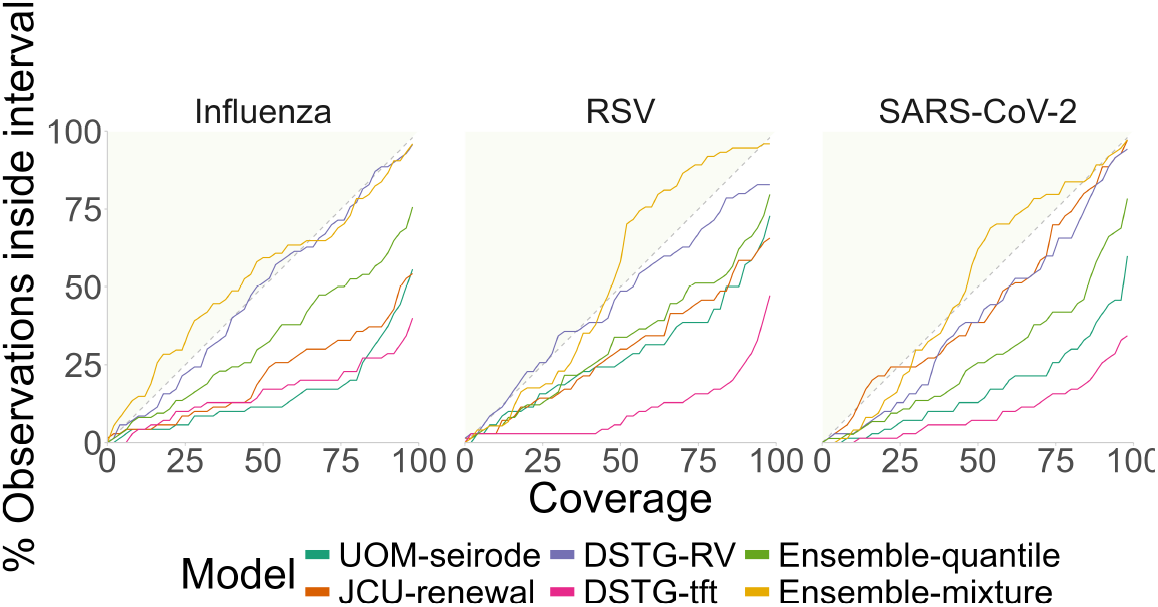
Interval coverage of each component model and ensemble forecast, with respect to the case data as reported at the end of the study period. Dashed lines indicate perfect interval coverage. Points below the dashed lines (white region) indicate overly confident/certain forecasts, points above the dashed lines (grey region) indicate overly uncertain forecasts.

## 4 Discussion

We have presented a performance evaluation of weekly forecasts of one-week, two-week, three-week and four-week ahead incident cases of influenza, SARS-CoV-2, and RSV in the Australian state of Victoria for the 2024 winter period. Our results support a growing body of literature showing that ensemble models typically outperform individual component forecasts in infectious disease prediction [10, 11]. This consistency reinforces the value of ensemble forecasting in operational public health contexts, particularly when model diversity is leveraged to capture a range of assumptions and uncertainties [15]. In our study, when evaluating the performance of forecasts by CRPS on either log-transformed or untransformed case counts, we found that the ensembles were frequently the top performing models for all pathogens. Component model performance rankings varied by forecast date, horizon, pathogen, and performance metric, indicating that each component model carried strengths and weaknesses. All component models were the top ranked component for at least one horizon pathogen combination under either CRPS or log-transformed CRPS. Notably, the DSTG–RV model was the top performing component model for all influenza forecast horizons for both CRPS and log-transformed CRPS.

A notable feature of our analysis is the change in model rankings depending on the scoring metric used. In particular, the DSTG-tft model performed relatively well under the CRPS but poorly under both the Dawid-Sebastiani Score (DSS) and log score (see supplementary material). This discrepancy highlights a broader methodological issue identified by Bosse *et al*. [28], whereby models that produce narrower predictive intervals may appear more accurate under CRPS but fail to capture the actual probability mass around the true outcome. Furthermore, the most appropriate performance metric will be context dependant and as discussed in [34] ideally motivated by the end user (in our case, public health officials).

The 2024 ensembles comprised of four models (two compartmental, one renewal, and one machine learning model). The more mechanistic type of models (compartment and renewal) tended to overshoot around epidemic peaks. This tendency may be attributed to the model’s assumptions of continuing trends i.e. that *R*_*t*_ will stay the same or drop (insufficiently for it to predict the peak) due to susceptible depletion. In contrast, the machine learning model (the DSTG-tft model) tended to undershoot the peaks. This potentially occurred due to training data of similar scenarios trending down earlier. The different types of biases exhibited by different models emphasises the importance of model diversity in forming the most accurate ensemble, compensating for the strengths and weaknesses of each component model.

Forecast performance around epidemic turning points, particularly peaks and resurgences, was poor across models. This aligns with prior findings from both FluSight [14] and the US COVID-19 Forecast Hub [35], where predictive performance tends to degrade around inflection points in observations. These periods are both highly policy-relevant and inherently challenging due to shifting transmission dynamics, behavioural changes, and possible data artifacts (e.g. reporting delays). Future ensemble designs may benefit from including models explicitly structured to detect and adapt to these turning points, as suggested by McGowan *et al*. [35]. Given this was a pilot study year for the ACEFA Forecasting Hub, such a target was not included and so has not been assessed here.

The ACEFA pilot study confirms the feasibility of a national (and expanding to trans-Tasman) collaborative forecasting hub. Tools adapted from the hubverse [22] ecosystem enabled streamlined submission, validation, and evaluation workflows, as also demonstrated in the ECDC’s RespiCast project [9]. With the infrastructure now in place, the ACEFA 2025 forecasting targets covered nine regions (Australia’s eight states/territories and New Zealand). Internationally, performance benchmarks increasingly include “epidemic” or “season” targets (e.g. onset, peak week, peak magnitude) [14, 35]. These targets were not part of the 2024 pilot season, but if included in future seasons, they could provided further insights into the effects of different modelling assumptions on predictive performance of epidemic peaks, particularly if the same models are used to predict both case trajectories and peak timing and magnitude.

Future work could include increasing the diversity of modelling methodologies (and therefore the number of models), adding extra data streams, and within season calibration monitoring. Increasing the number of models would encourage a greater diversity of methods and model structures (e.g. agent-based, machine-learning, statistical, mechanistic) as recommended by [11]. While the number of models in our 2024 ensemble is comparatively small to Forecasting Hubs in Europe (RespiCast containing more than 10 models [36]) and North America (US CDC greater than 20 models [37]), recent research has found that only four components are needed to ensure robust ensemble accuracy, with additional models producing diminishing returns [38]. Inclusion of other data sources, for example: mobility, climate, and syndromic data, or web search trends, past epidemic trends, or trends from other locations, may improve model performance. Examples of this is the utilisation of internet search queries in [39] or waste water data [40]. However, incorporating multiple data sources will not necessarily provide performance benefits over analysing a single dataset. For example, Klaassen and colleagues [40] found that adding waste water data reduced the predictive performance of forecasts compared to a model fitted to deaths data alone. Similarly, Moss and colleagues [41] found that simultaneously using data from three different influenza surveillance systems only improved (compared to using a single data source) influenza forecasts under certain circumstances and sometimes reduced forecast performance. They proposed that multi-data source data forecasting may only provide performance benefit if each dataset captures distinct, but complementary, aspects of the biological or surveillance processes. Ongoing assessment of forecast interval coverage, such as seen in Figure 5, should be used not only for evaluation but also for model development. Such an approach is proposed by Rumack and colleagues [42] to “recalibrate” a model using past forecasts to form a weighted ensemble of base recalibration methods for application to the forecasting model. Alternatively, interval coverage scores may be used for model rejection as suggested by Funk and colleagues [43].

With further model development and improvements in evaluations (including of the predicted peak timing and magnitude), we anticipate that our forecasting ensemble exercise will not only be useful in projecting viral respiratory pathogens activity and burden, but may lead to insights around the cause for poor predictions around the peak. The 2024 pilot has established ACEFA as a viable framework for collaborative, operational forecasting in Australia and Aotearoa New Zealand. Expanding geographic coverage and enhancing model robustness will be critical to supporting timely public health decision-making in future seasons.

## Data Availability

Code and data for reproducing the analysis is available at https://github.com/acefa-hubs/Winter-forecasting-of-respiratory-viruses-in-Victoria-Australia-2024

## 5 Ethics

This study received research ethics approval from the University of Melbourne Human Research Ethics Committee (application identifier 2024-26949-50575-3).

## 9 Acknowledgements

We wish to acknowledge the medical practitioners and pathology services who diagnosed and notified the cases to the Department of Health, Victoria. We also acknowledge the public health officers, medical officers, epidemiologists and systems specialists who have also contributed to the collection and curation of this dataset.

