## Supplementary file for "Winter forecasting of respiratory viruses in Victoria Australia"

### 1 Introduction

This supplement file adds more detail to the methods and results presented in the main paper. The supplements sections and subsections align with the main text (expanding on the corresponding section).

### 2 Data and methods

#### 2.1 Evaluation metrics

Here we present the metrics used in this study with $F$ being the predictive distribution $F_c$ being the cumulative distribution of $F$ and $y$ being the true number of cases. $\mu$ is the mean of $F$ and $\sigma$ the standard deviation: The log score (also sometimes called ignorance score):

$$LogS(F, y) = -log(F(y))s \tag{1}$$

The Dawid-Sebastiani Score (dss):

$$dss(F, y) = \left(\frac{y - \mu}{\sigma}\right)^2 + 2\log(\mu) \tag{2}$$

### 3 Results

#### 3.1 Four week Forecasts

Here we present the four week ahead forecasts. They are grouped by model. They show non overlapping forecasts for four weeks based on the different forecast date.

##### 3.1.1 UOM-SEIRODE

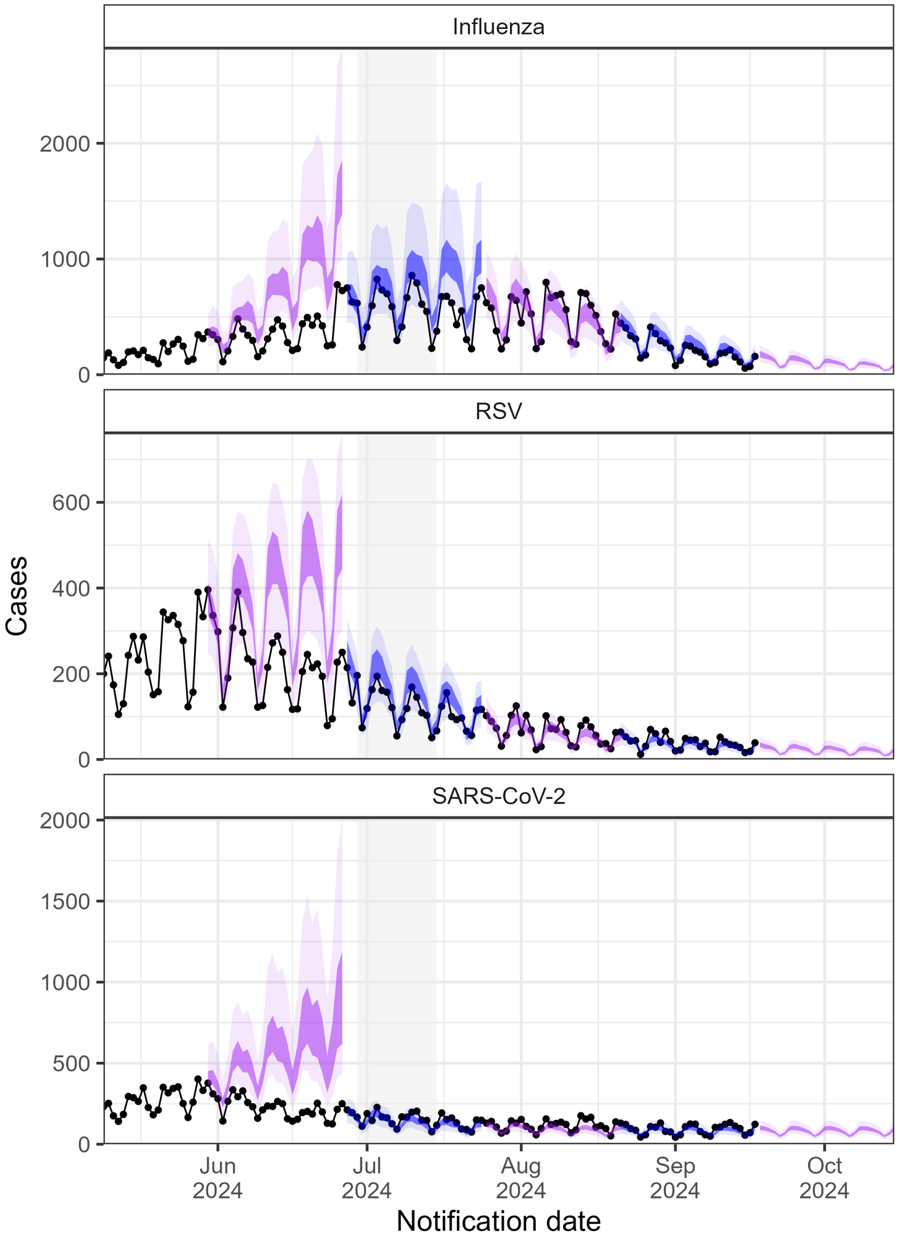

Figure 1: Four week ahead forecasts for uom-seirode. The vertical shading represents the winter two-week school holidays. The lightest shading represents the 95% prediction interval and the darker the 50% prediction interval. Alternating colours are used to make it easier to visualise a new forecast date.

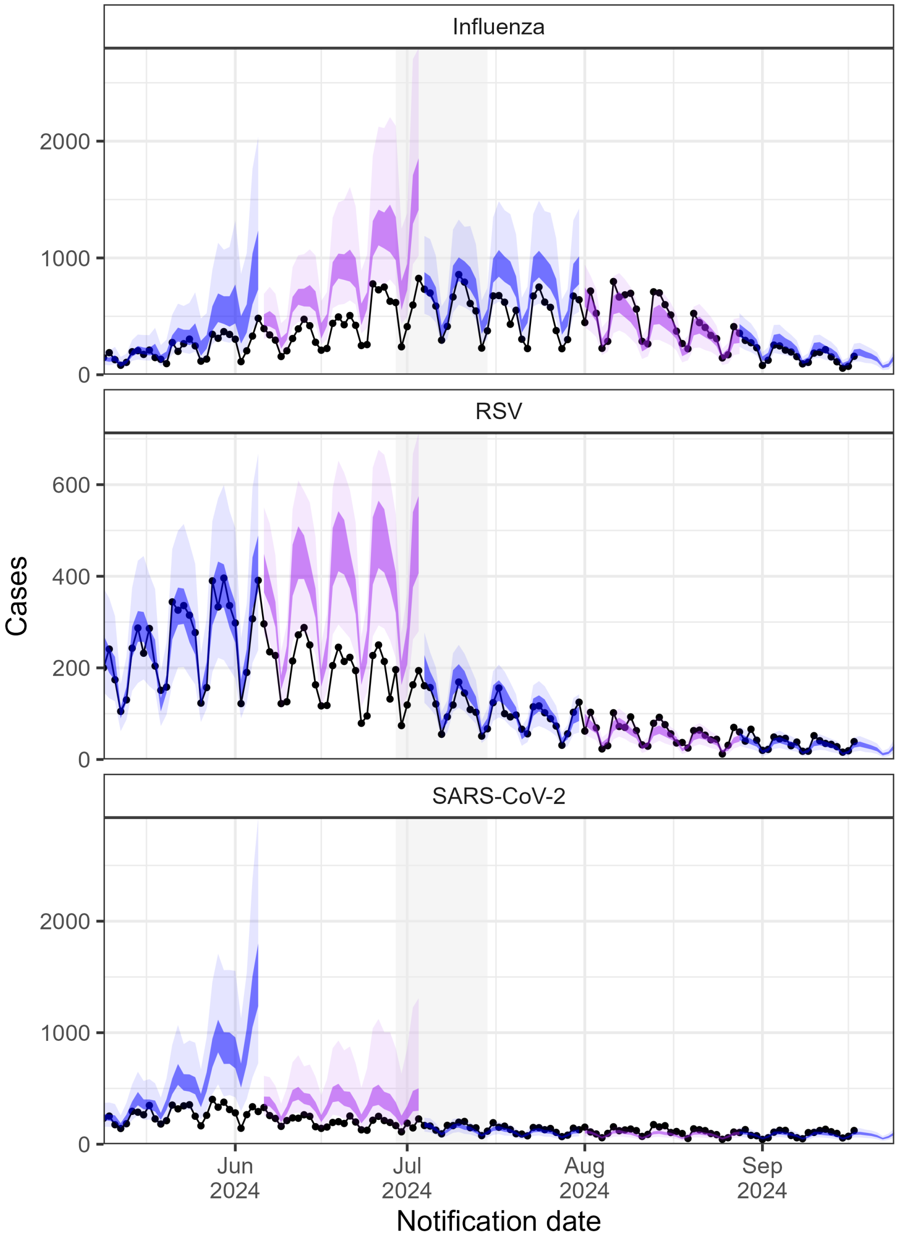

Figure 2: Four week ahead forecasts for uom-seirode. The vertical shading represents the winter two-week school holidays. The lightest shading represents the 95% prediction interval and the darker the 50% prediction interval. Alternating colours are used to make it easier to visualise a new forecast date.

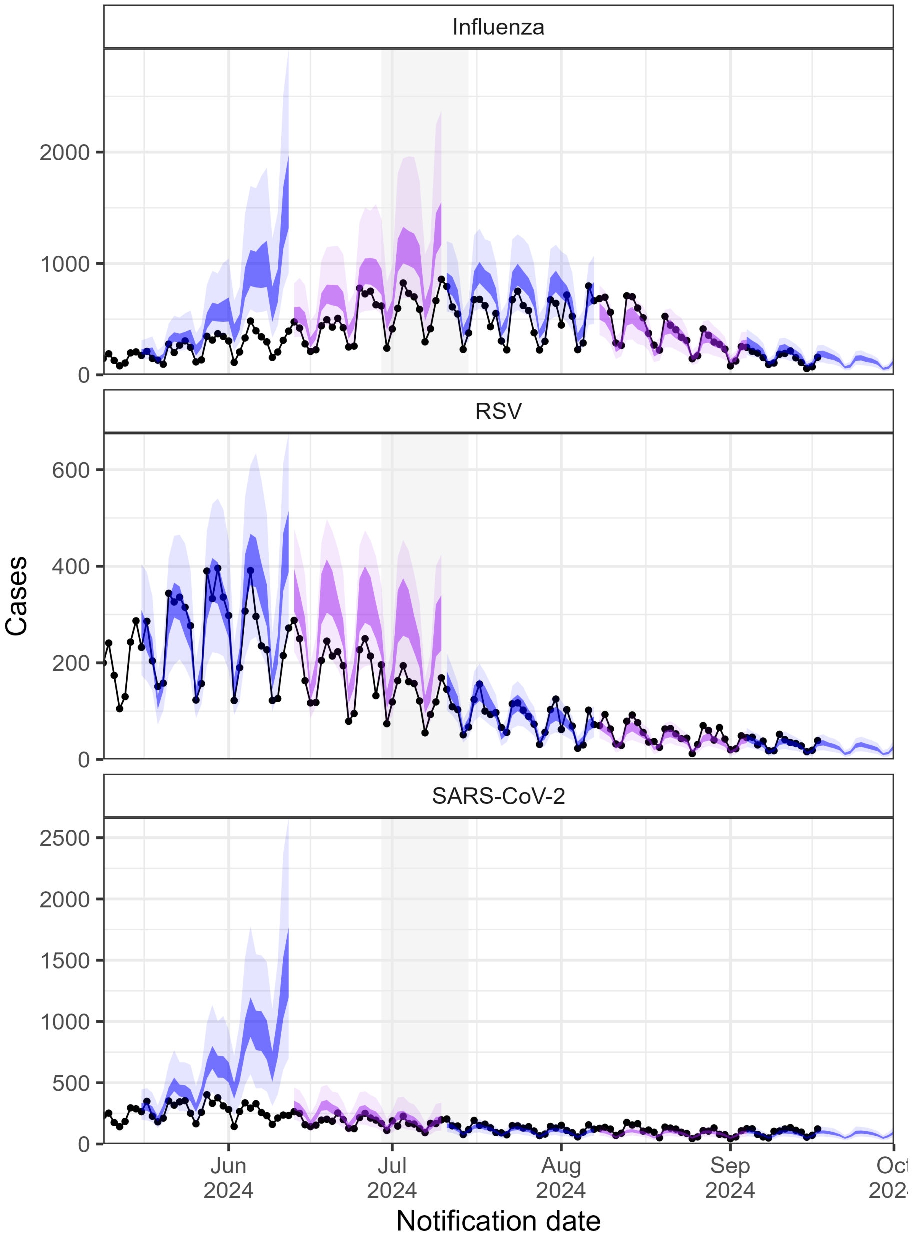

Figure 3: Four week ahead forecasts for uom-seirode. The vertical shading represents the winter two-week school holidays. The lightest shading represents the 95% prediction interval and the darker the 50% prediction interval. Alternating colours are used to make it easier to visualise a new forecast date.

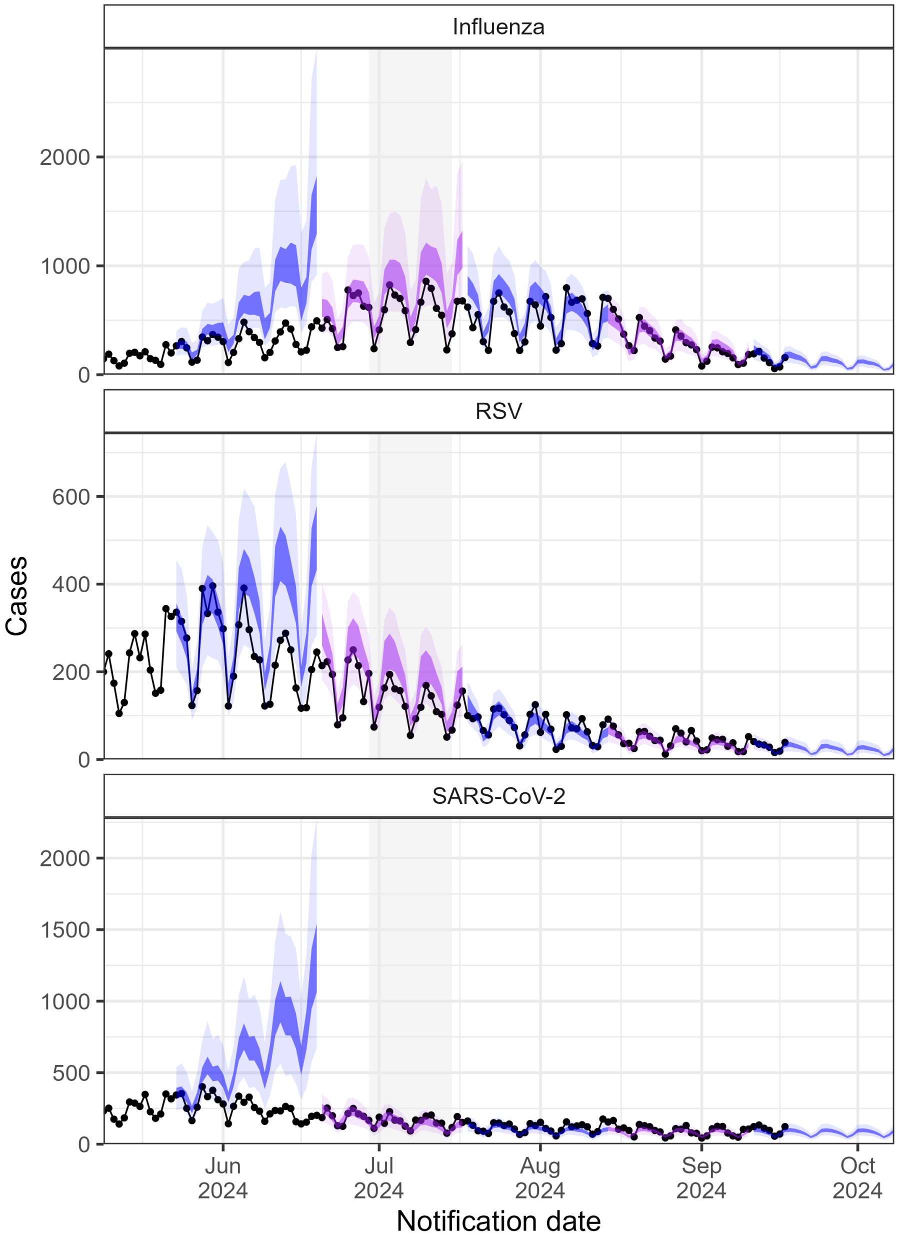

Figure 4: Four week ahead forecasts for uom-seirode. The vertical shading represents the winter two-week school holidays. The lightest shading represents the 95% prediction interval and the darker the 50% prediction interval. Alternating colours are used to make it easier to visualise a new forecast date.

##### 3.1.2 JCU-RENEWAL

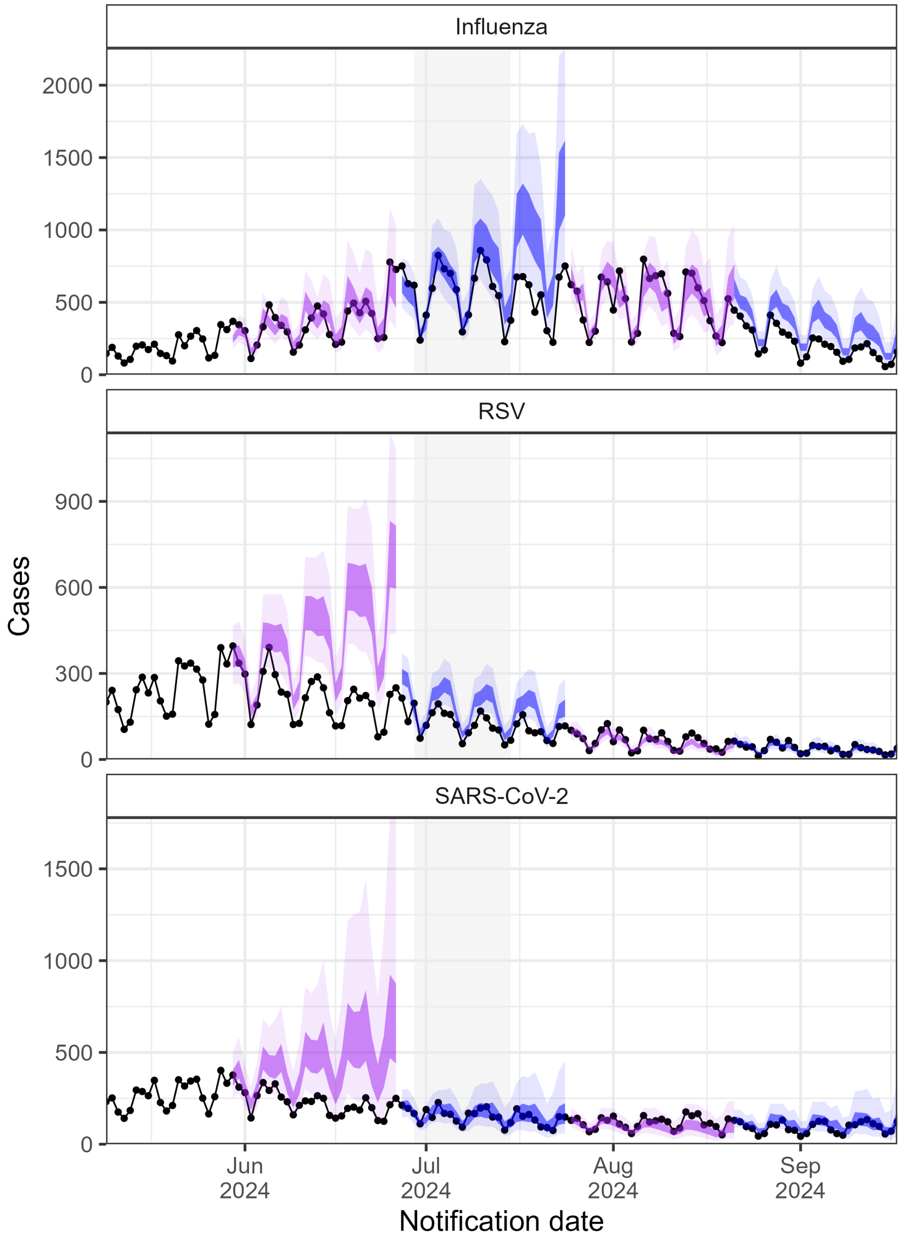

Figure 5: Four week ahead forecasts for JCU-renewal. The vertical shading represents the winter two-week school holidays. The lightest shading represents the 95% prediction interval and the darker the 50% prediction interval. Alternating colours are used to make it easier to visualise a new forecast date.

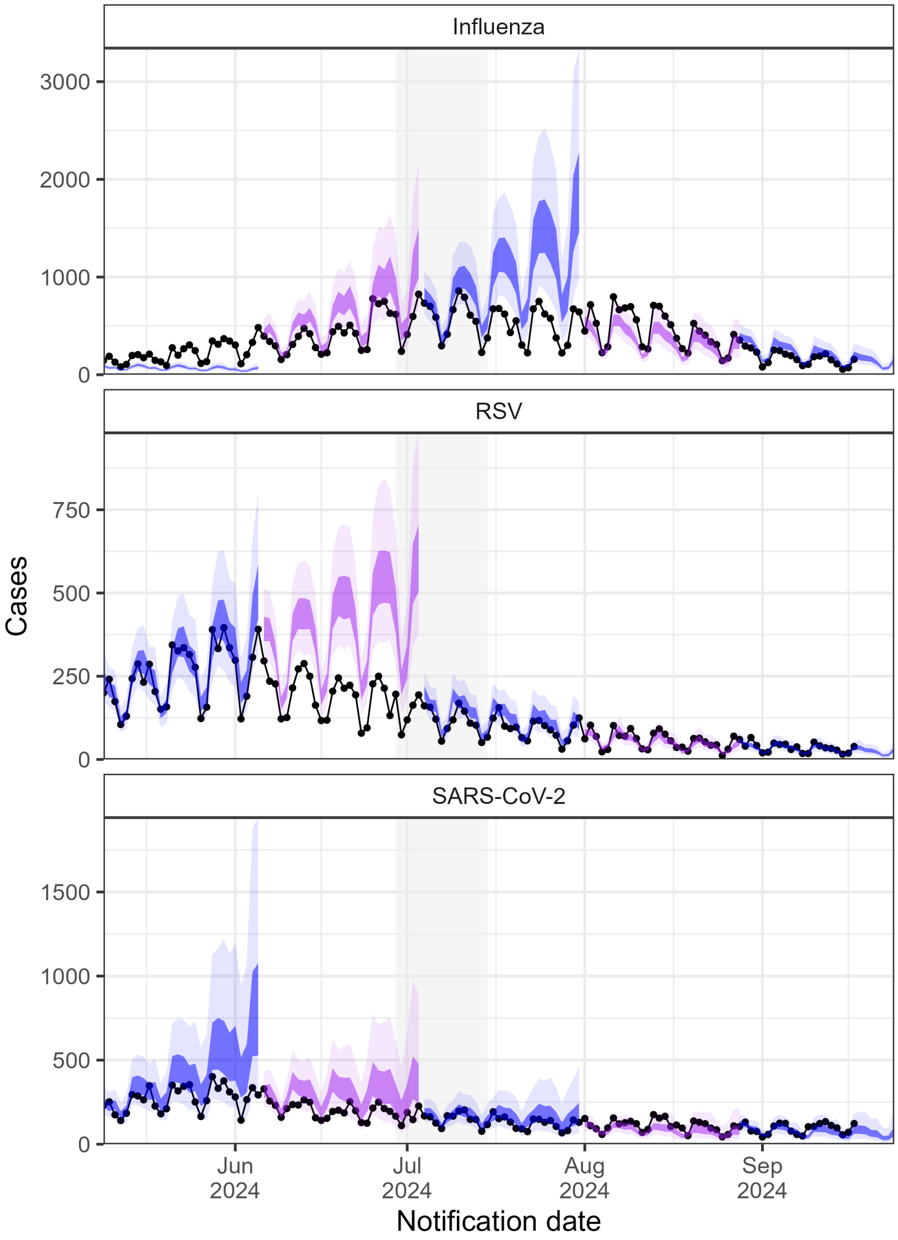

Figure 6: Four week ahead forecasts for JCU-renewal. The vertical shading represents the winter two-week school holidays. The lightest shading represents the 95% prediction interval and the darker the 50% prediction interval. Alternating colours are used to make it easier to visualise a new forecast date.

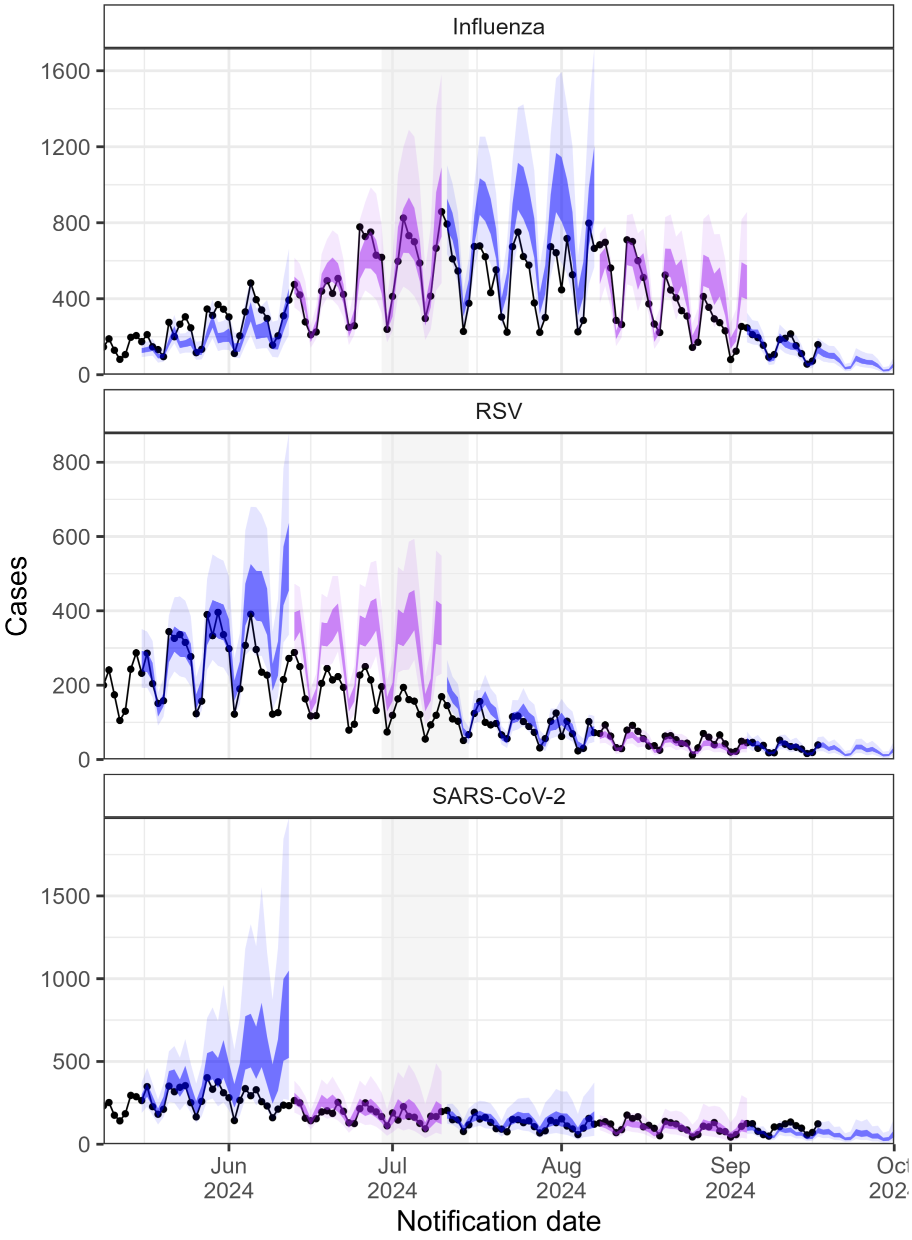

Figure 7: Four week ahead forecasts for JCU-renewal. The vertical shading represents the winter two-week school holidays. The lightest shading represents the 95% prediction interval and the darker the 50% prediction interval. Alternating colours are used to make it easier to visualise a new forecast date.

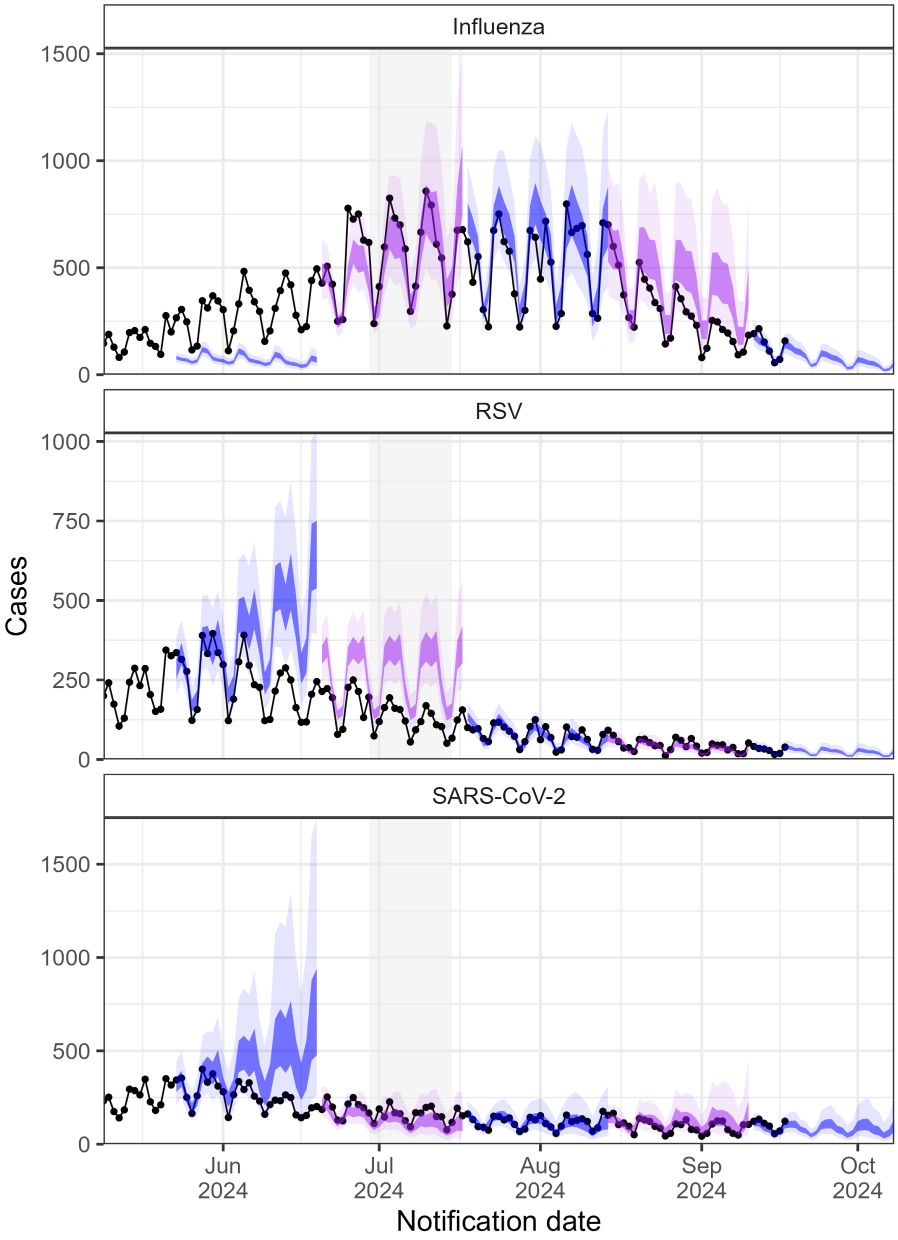

Figure 8: Four week ahead forecasts for JCU-renewal. The vertical shading represents the winter two-week school holidays. The lightest shading represents the 95% prediction interval and the darker the 50% prediction interval. Alternating colours are used to make it easier to visualise a new forecast date.

##### 3.1.3 DSTG-TFT

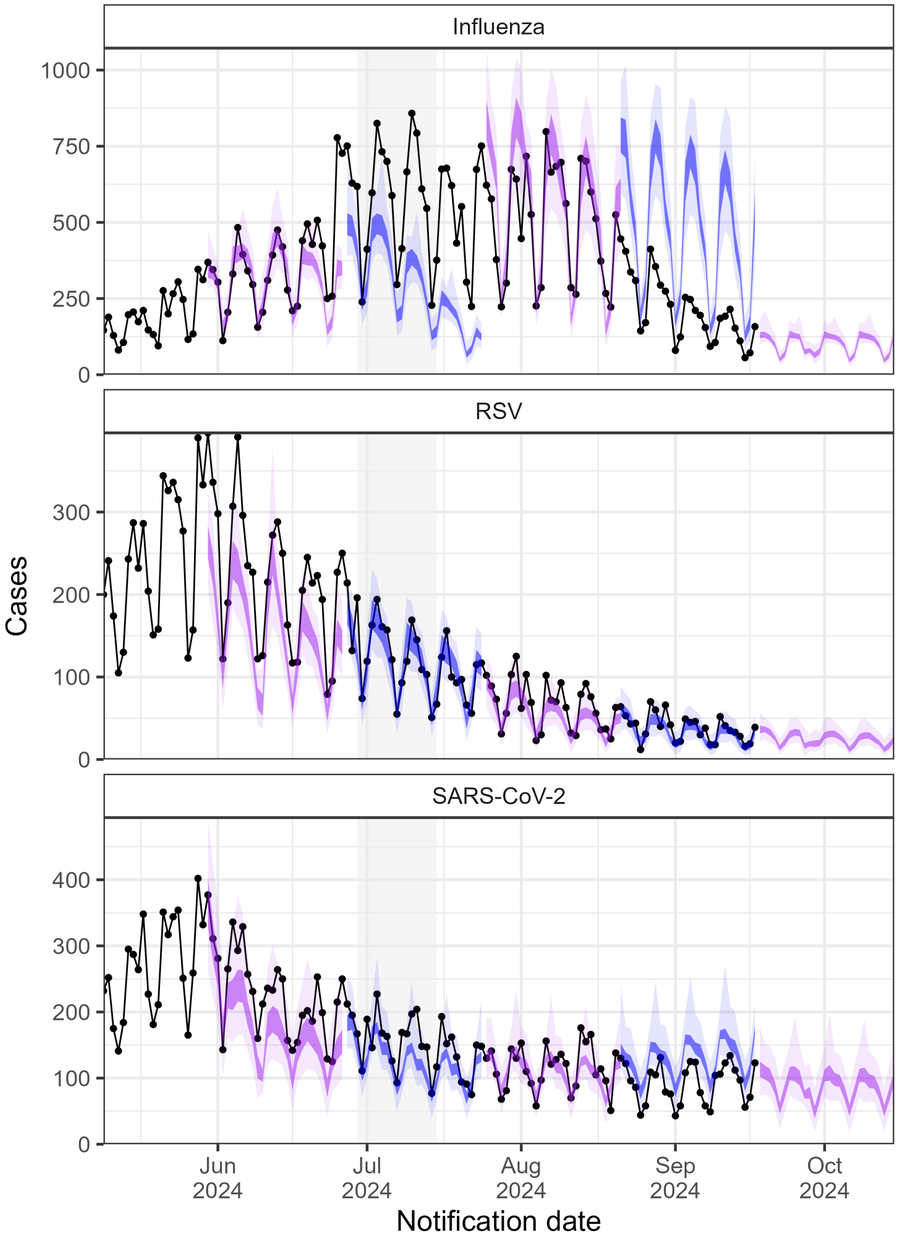

Figure 9: Four week ahead forecasts for dstg-tft. The vertical shading represents the winter two-week school holidays. The lightest shading represents the 95% prediction interval and the darker the 50% prediction interval. Alternating colours are used to make it easier to visualise a new forecast date.

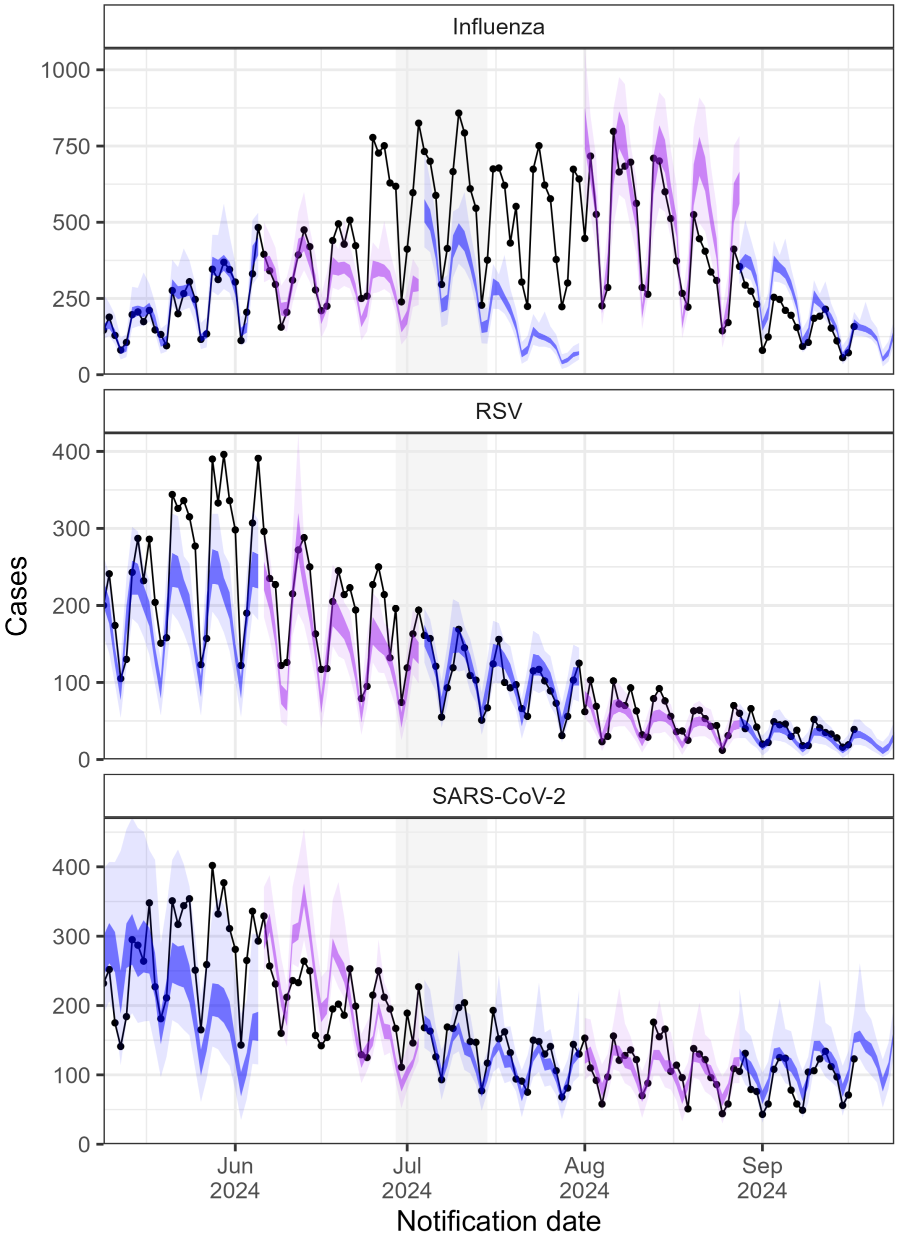

Figure 10: Four week ahead forecasts for dstg-tft. The vertical shading represents the winter two-week school holidays. The lightest shading represents the 95% prediction interval and the darker the 50% prediction interval. Alternating colours are used to make it easier to visualise a new forecast date.

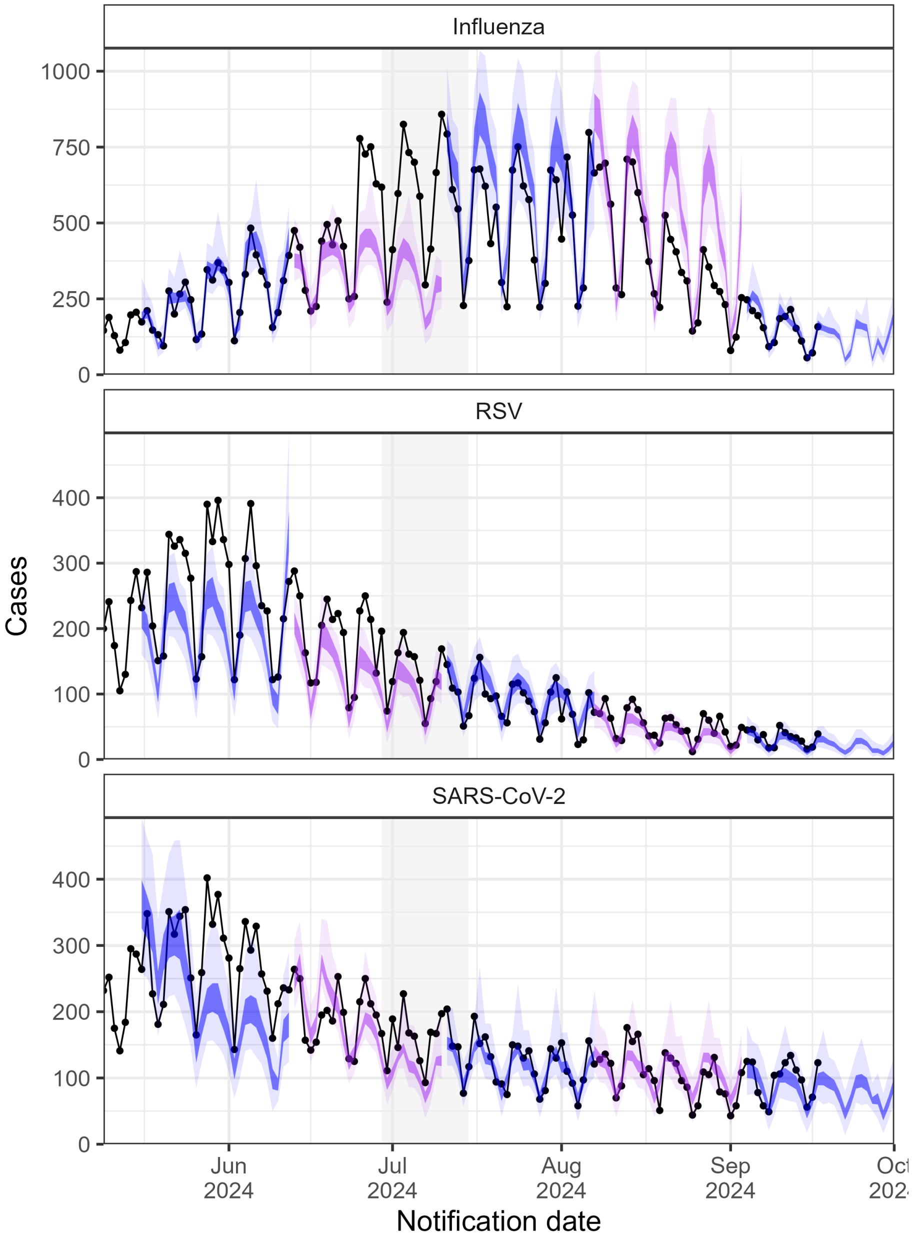

Figure 11: Four week ahead forecasts for dstg-tft. The vertical shading represents the winter two-week school holidays. The lightest shading represents the 95% prediction interval and the darker the 50% prediction interval. Alternating colours are used to make it easier to visualise a new forecast date.

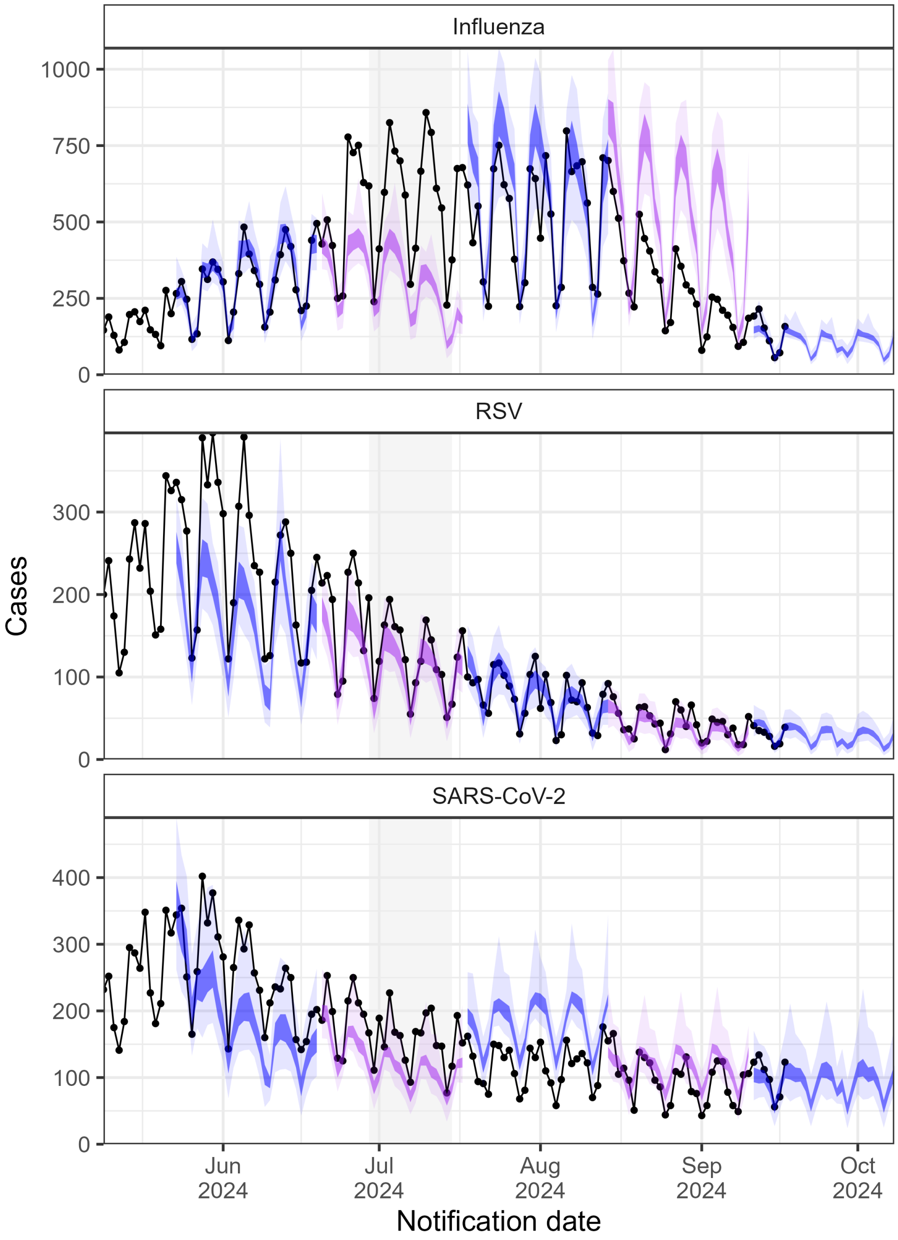

Figure 12: Four week ahead forecasts for dstg-tft. The vertical shading represents the winter two-week school holidays. The lightest shading represents the 95% prediction interval and the darker the 50% prediction interval. Alternating colours are used to make it easier to visualise a new forecast date.

###### 3.1.4 DSTG-RV

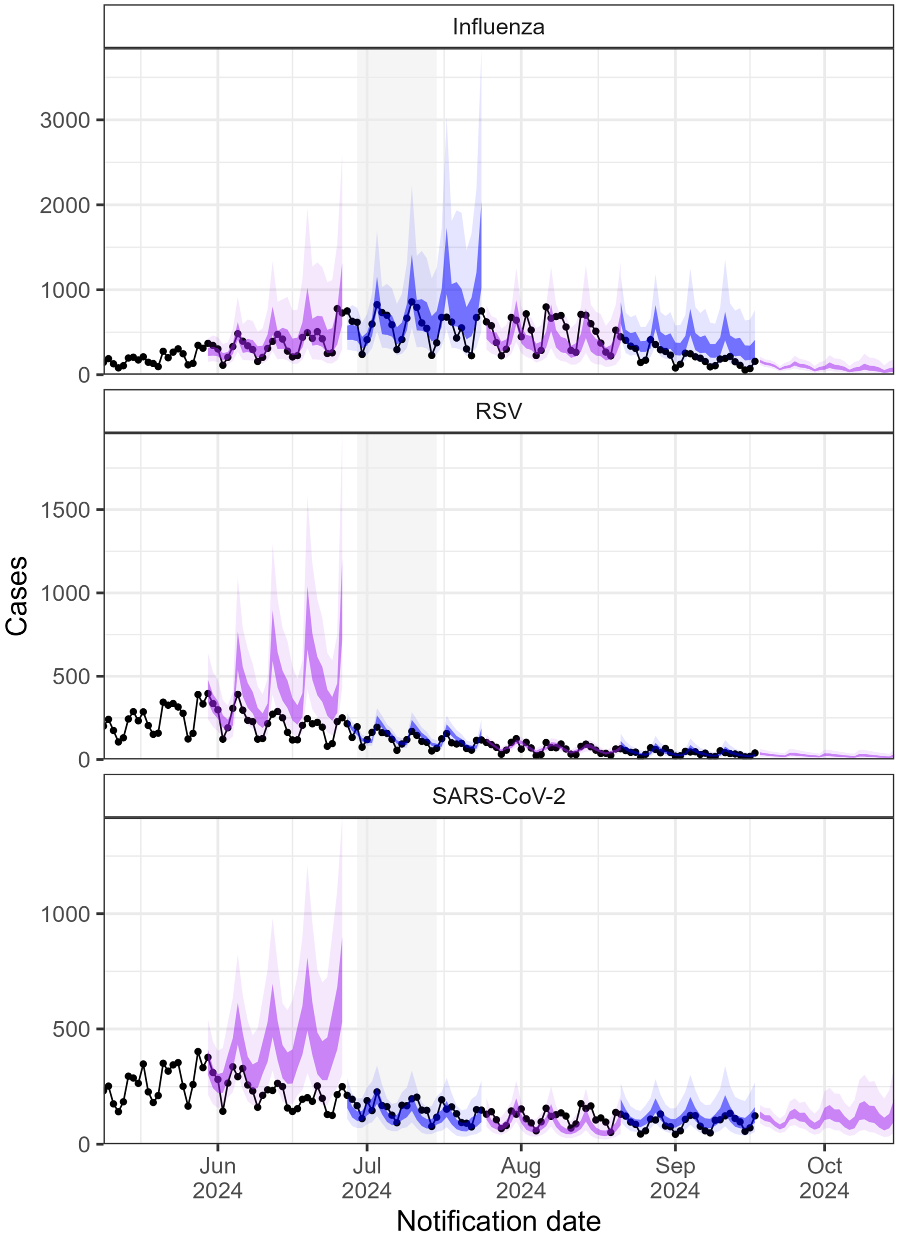

Figure 13: Four week ahead forecasts for dstg-RV. The vertical shading represents the winter two-week school holidays. The lightest shading represents the 95% prediction interval and the darker the 50% prediction interval. Alternating colours are used to make it easier to visualise a new forecast date.

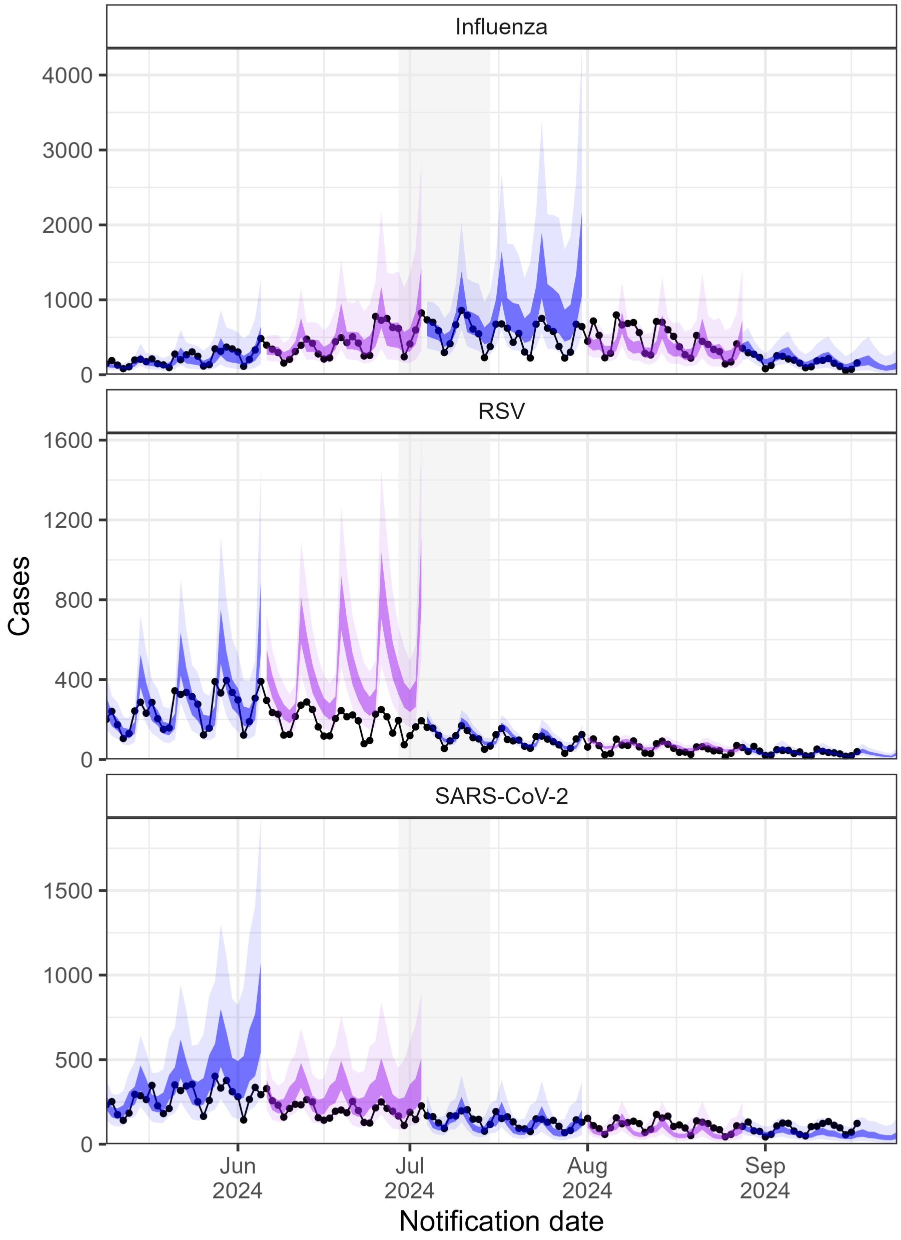

Figure 14: Four week ahead forecasts for dstg-RV. The vertical shading represents the winter two-week school holidays. The lightest shading represents the 95% prediction interval and the darker the 50% prediction interval. Alternating colours are used to make it easier to visualise a new forecast date.

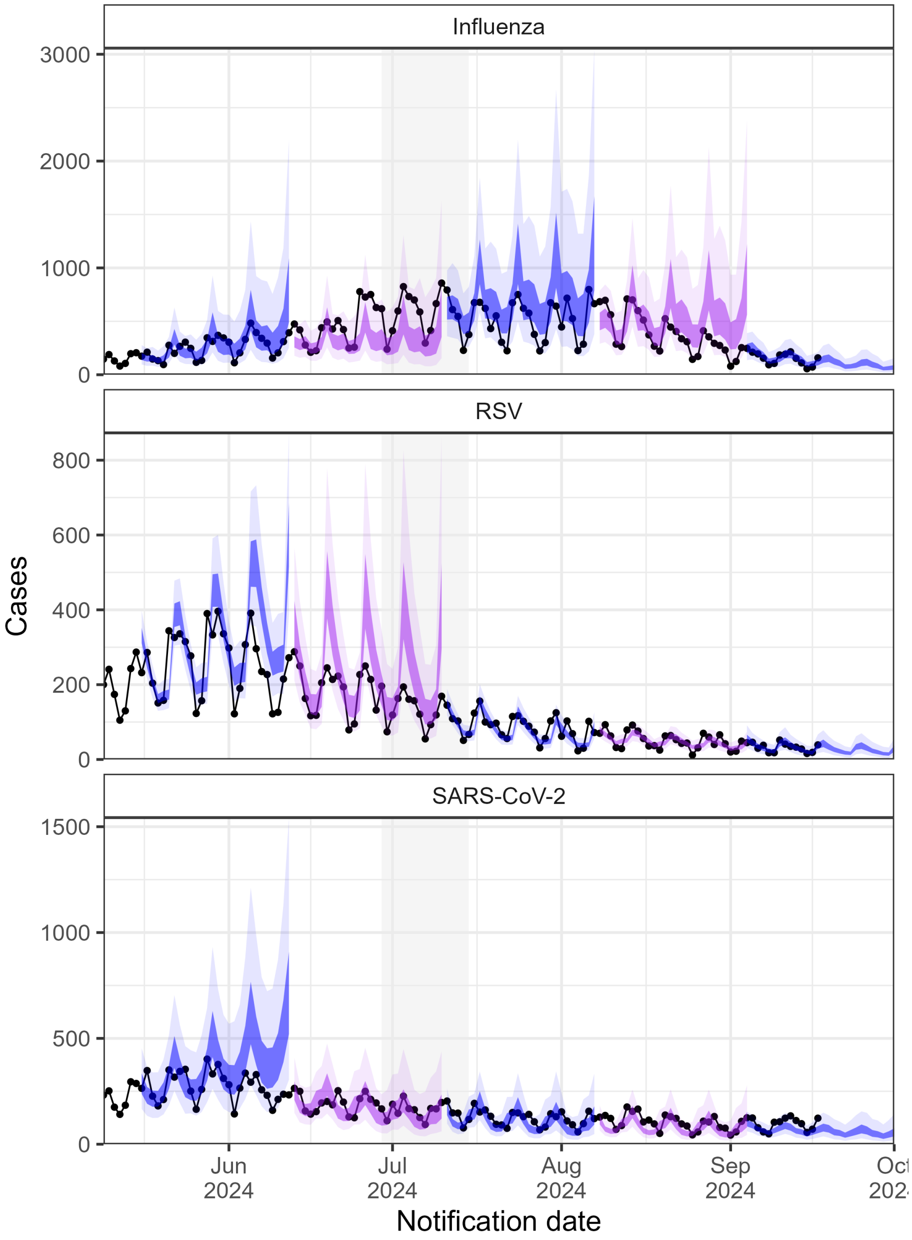

Figure 15: Four week ahead forecasts for dstg-RV. The vertical shading represents the winter two-week school holidays. The lightest shading represents the 95% prediction interval and the darker the 50% prediction interval. Alternating colours are used to make it easier to visualise a new forecast date.

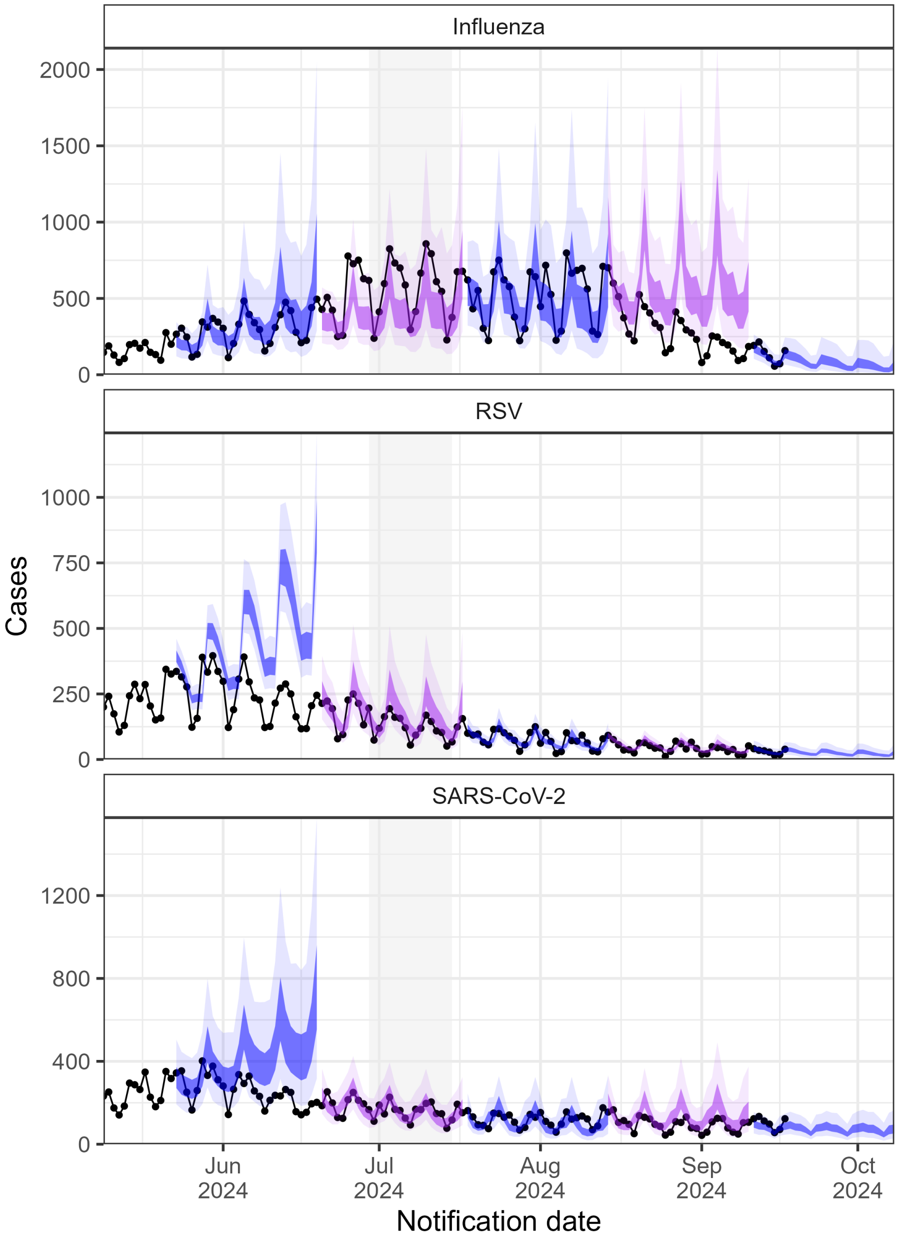

Figure 16: Four week ahead forecasts for dstg-RV. The vertical shading represents the winter two-week school holidays. The lightest shading represents the 95% prediction interval and the darker the 50% prediction interval. Alternating colours are used to make it easier to visualise a new forecast date.

###### 3.1.5 ENSEMBLE-MIXTURE

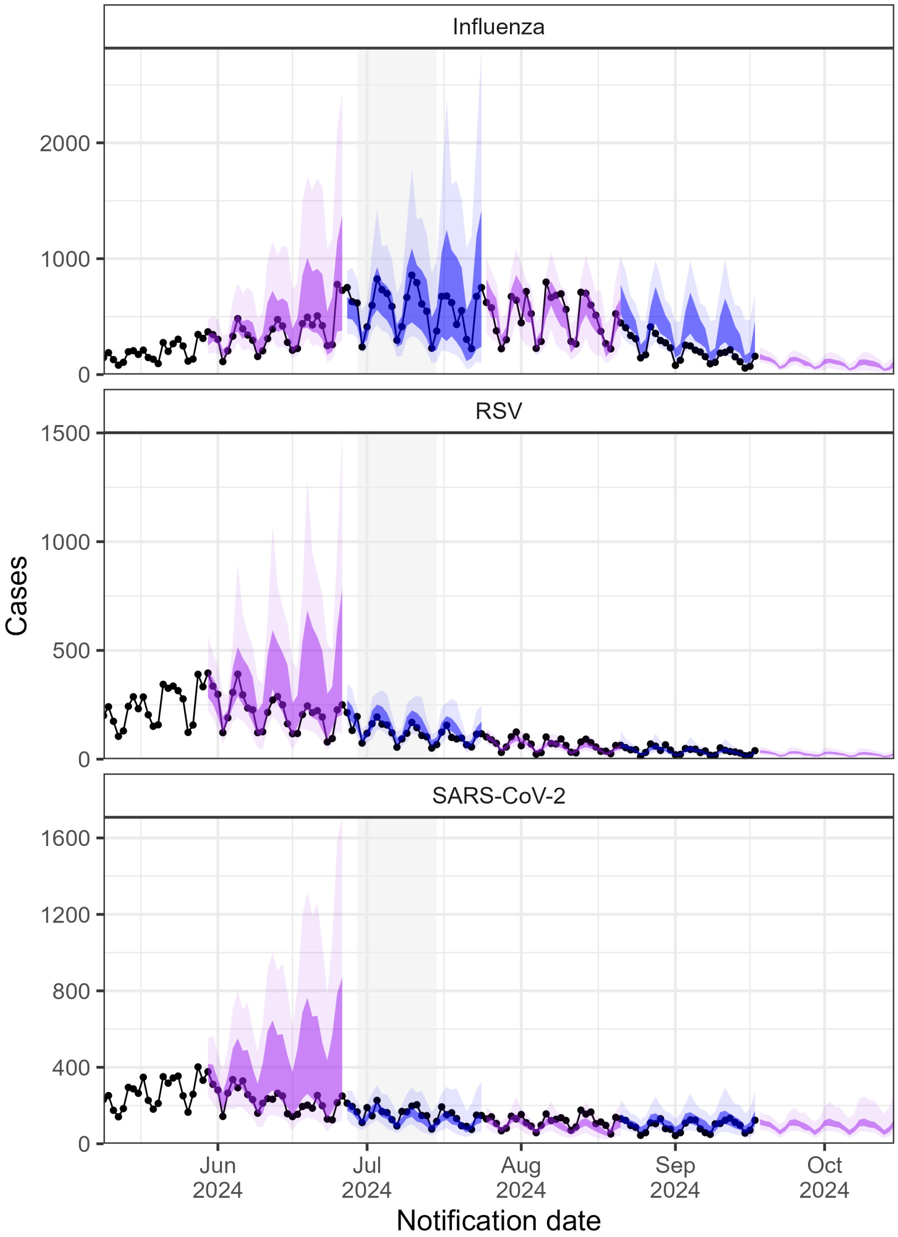

Figure 17: Four week ahead forecasts for ensemble-mixture. The vertical shading represents the winter two-week school holidays. The lightest shading represents the 95% prediction interval and the darker the 50% prediction interval. Alternating colours are used to make it easier to visualise a new forecast date.

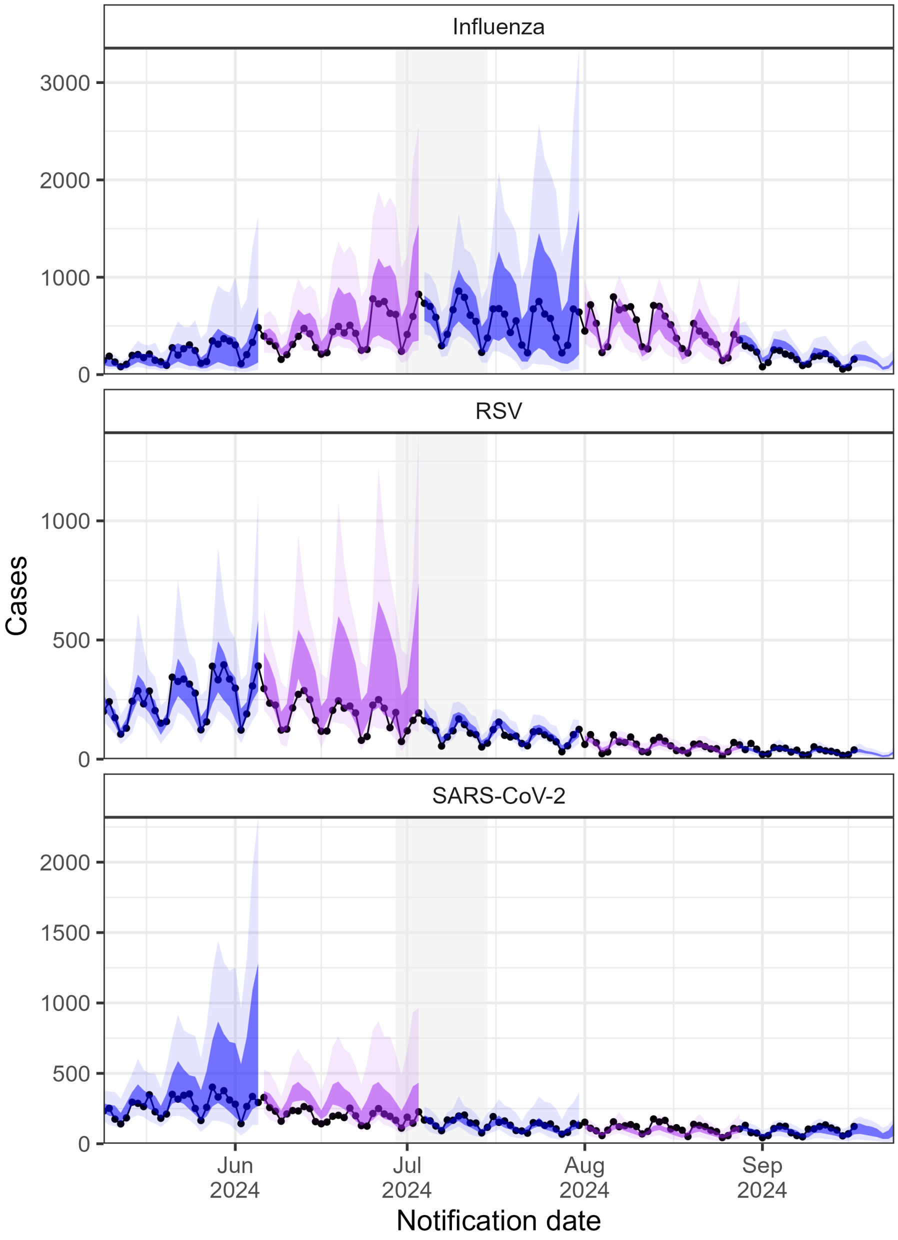

Figure 18: Four week ahead forecasts for ensemble-mixture. The vertical shading represents the winter two-week school holidays. The lightest shading represents the 95% prediction interval and the darker the 50% prediction interval. Alternating colours are used to make it easier to visualise a new forecast date.

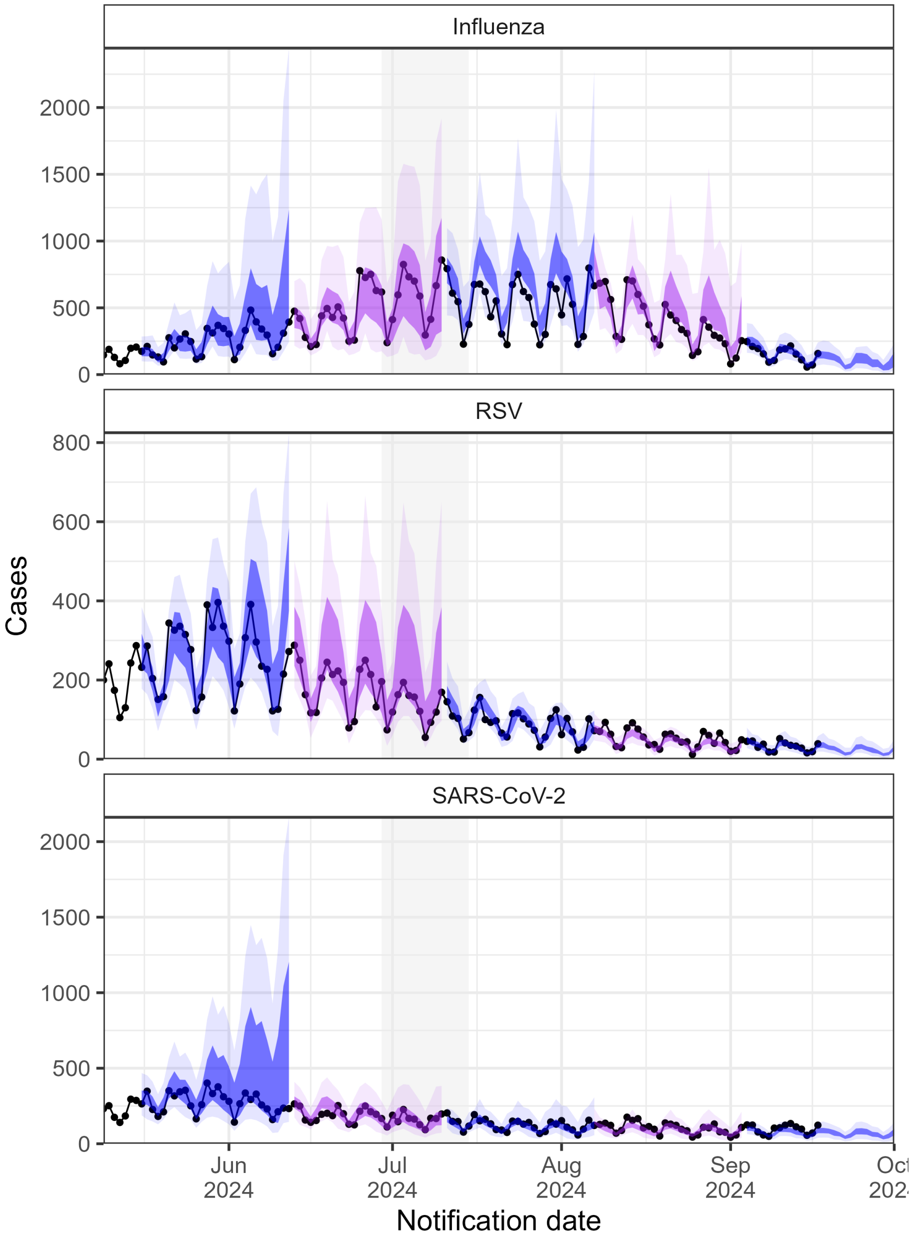

Figure 19: Four week ahead forecasts for ensemble-mixture. The vertical shading represents the winter two-week school holidays. The lightest shading represents the 95% prediction interval and the darker the 50% prediction interval. Alternating colours are used to make it easier to visualise a new forecast date.

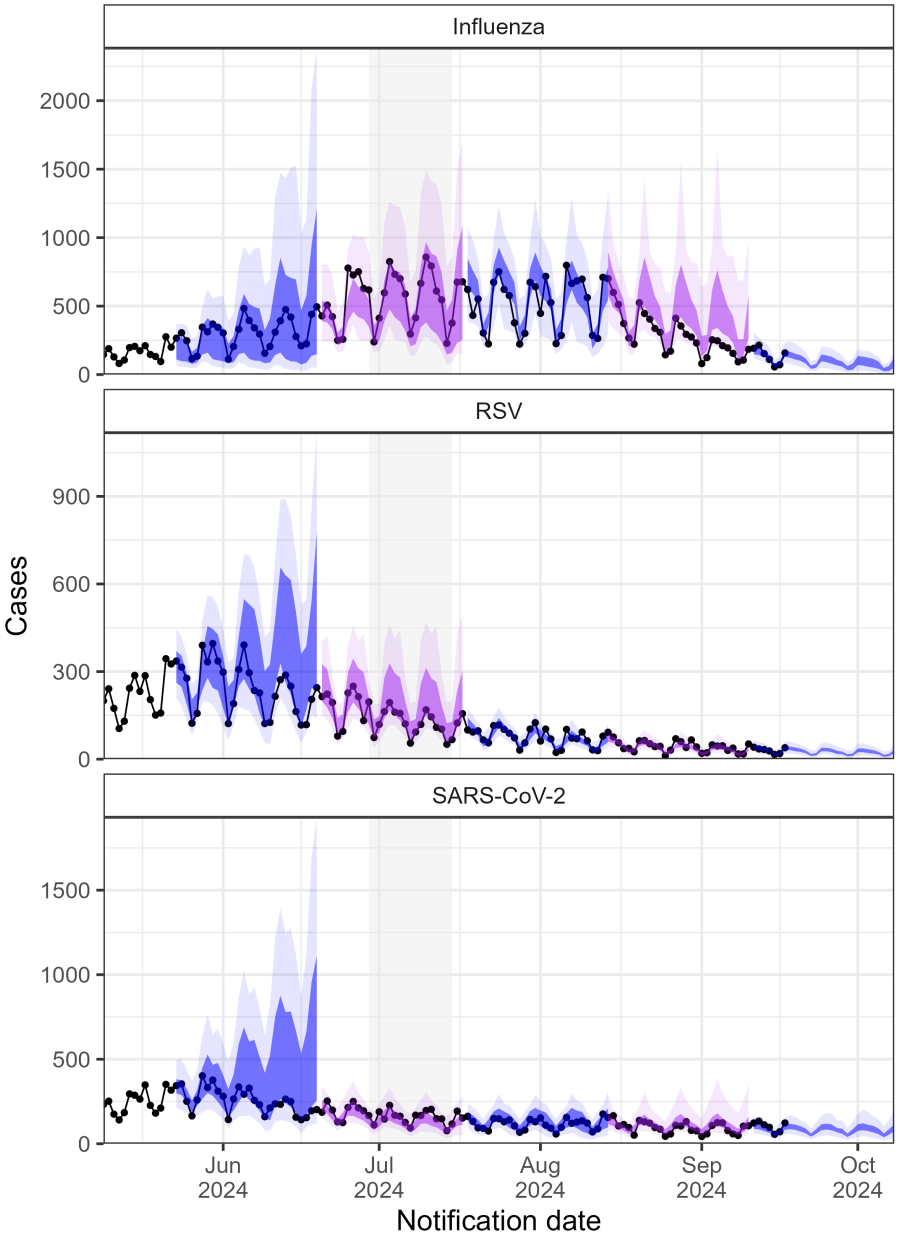

Figure 20: Four week ahead forecasts for ensemble-mixture. The vertical shading represents the winter two-week school holidays. The lightest shading represents the 95% prediction interval and the darker the 50% prediction interval. Alternating colours are used to make it easier to visualise a new forecast date.

##### 3.1.6 ENSEMBLE-QUANTILE

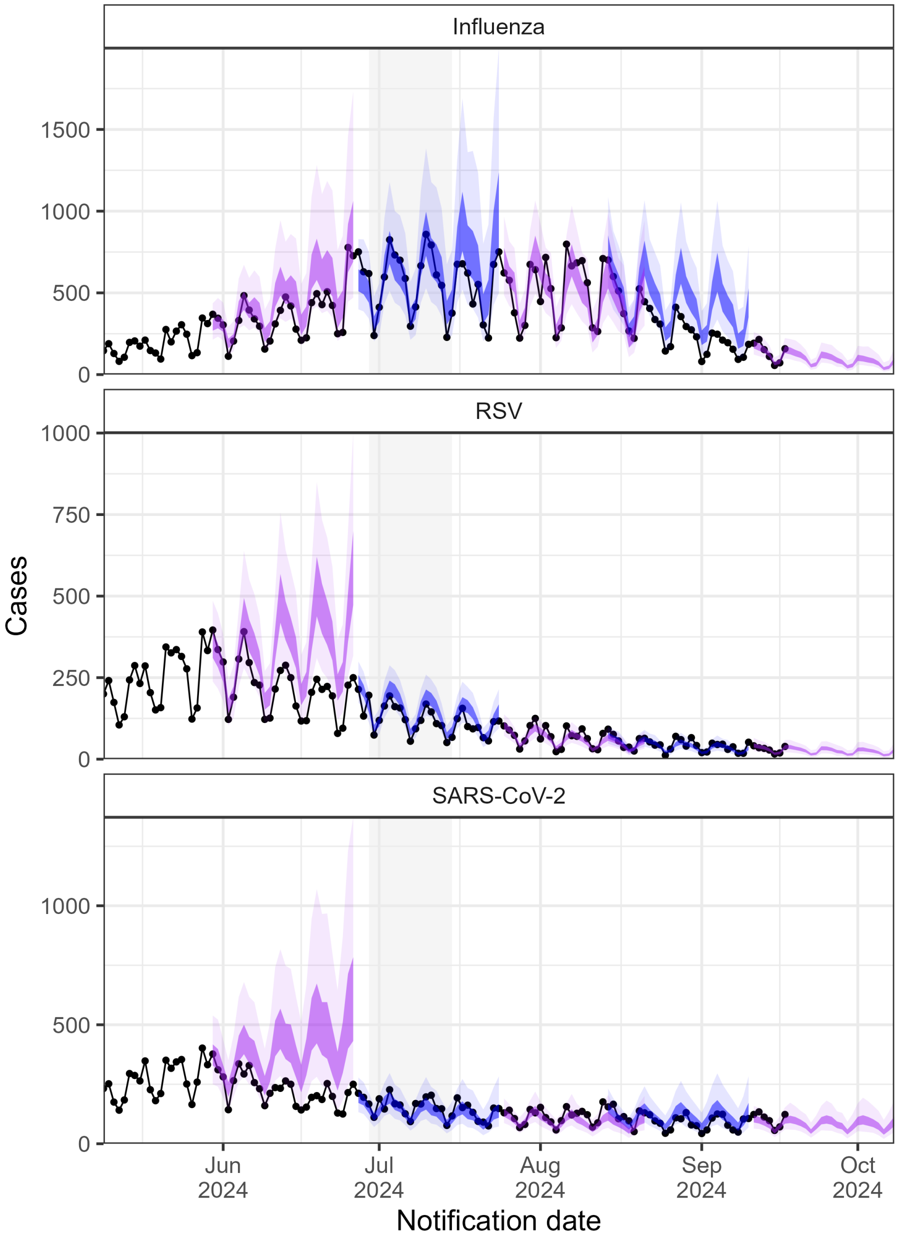

Figure 21: Four week ahead forecasts for ensemble-quantile. The vertical shading represents the winter two-week school holidays. The lightest shading represents the 95% prediction interval and the darker the 50% prediction interval. Alternating colours are used to make it easier to visualise a new forecast date.

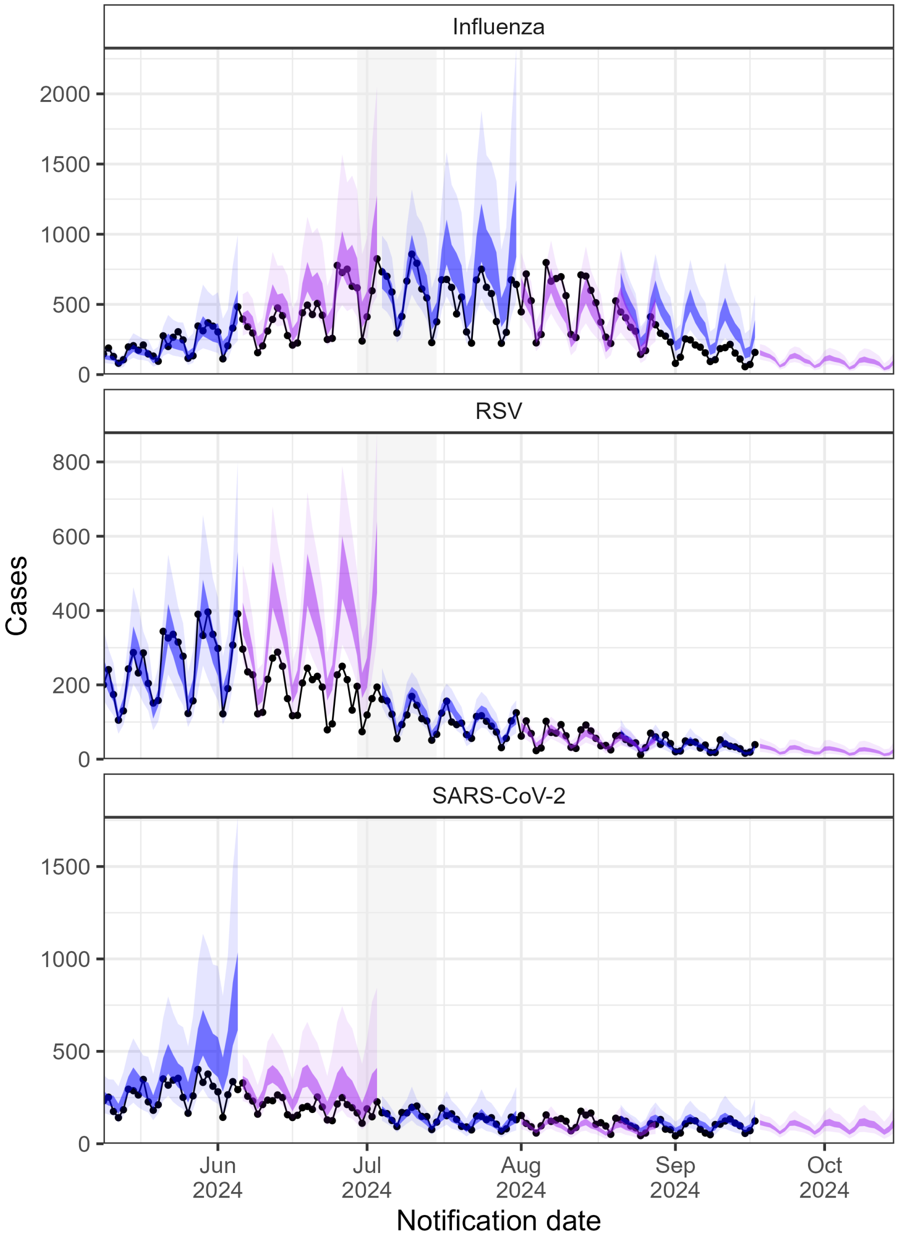

Figure 22: Four week ahead forecasts for ensemble-quantile. The vertical shading represents the winter two-week school holidays. The lightest shading represents the 95% prediction interval and the darker the 50% prediction interval. Alternating colours are used to make it easier to visualise a new forecast date.

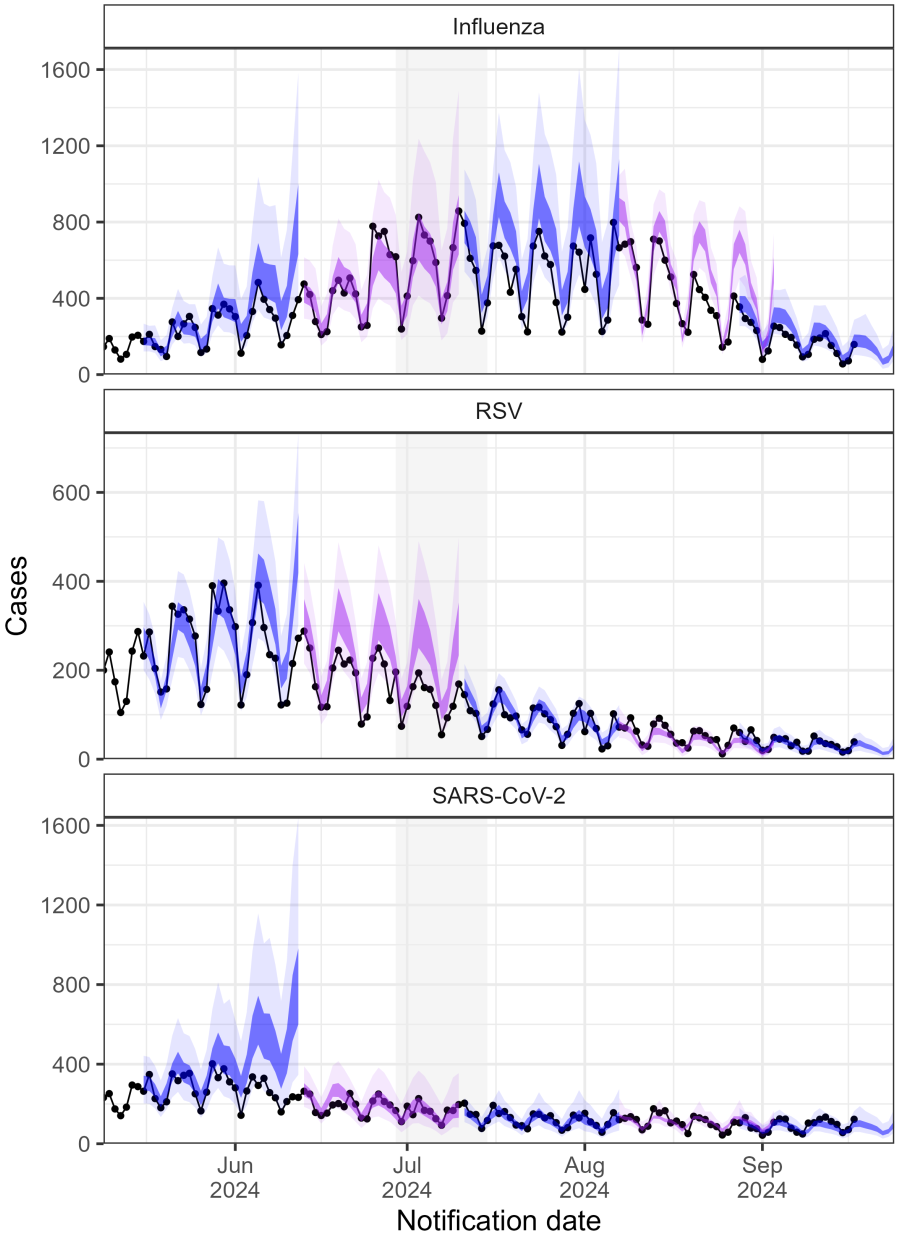

Figure 23: Four week ahead forecasts for ensemble-quantile. The vertical shading represents the winter two-week school holidays. The lightest shading represents the 95% prediction interval and the darker the 50% prediction interval. Alternating colours are used to make it easier to visualise a new forecast date.

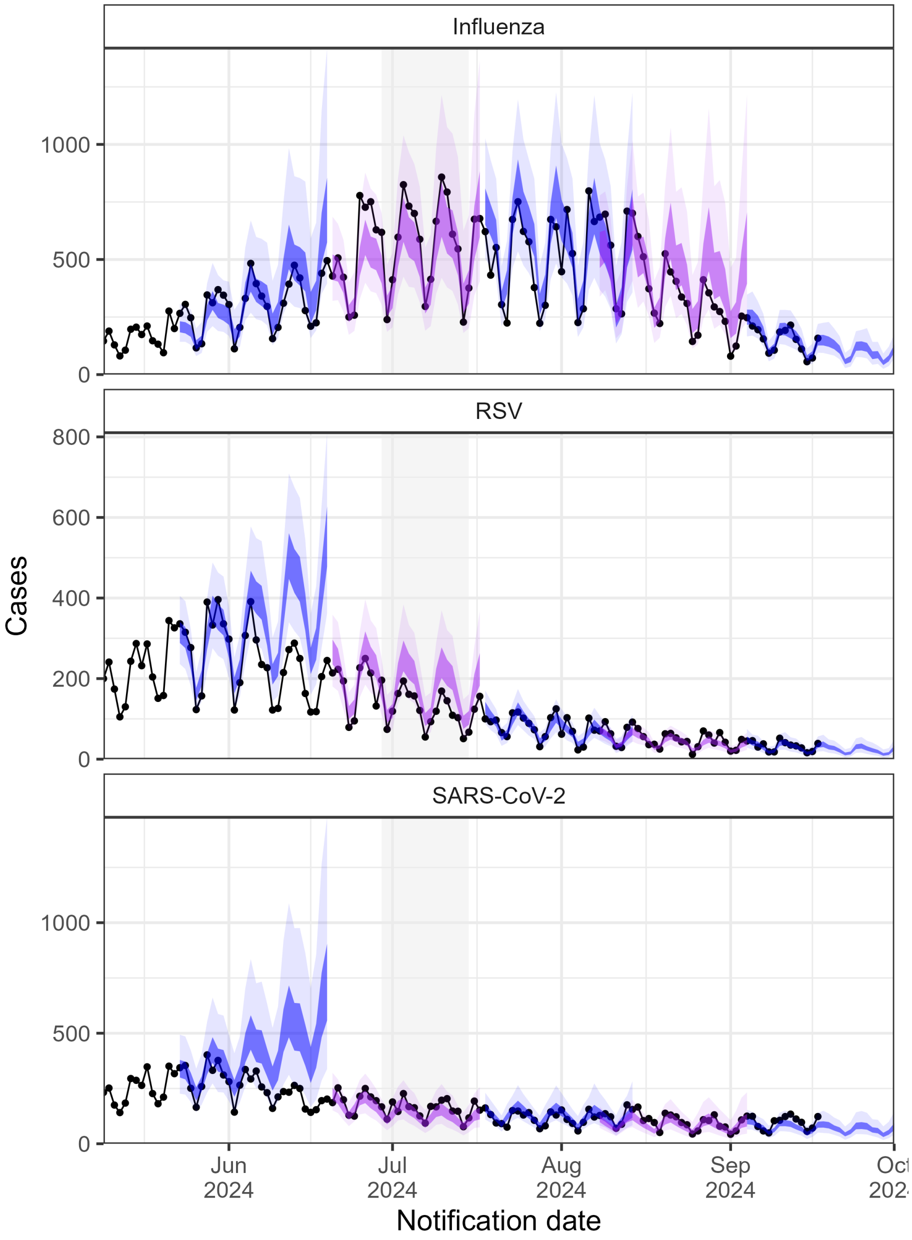

Figure 24: Four week ahead forecasts for ensemble-quantile. The vertical shading represents the winter two-week school holidays. The lightest shading represents the 95% prediction interval and the darker the 50% prediction interval. Alternating colours are used to make it easier to visualise a new forecast date.

#### 3.2 Forecast date

Tables 4 through 22 provide full metrics for each forecasts date.

| Model | Path | Scale | Bias | Dss | Crps | Op | Up | Disp | LS |
|---|---|---|---|---|---|---|---|---|---|
| Baseline | COV | Log | -0.68 | -2.18 | 0.13 | 0.00 | 0.08 | 0.05 | -0.17 |
| DTSG-RV | COV | Log | 0.61 | -1.71 | 0.18 | 0.11 | 0.00 | 0.07 | 0.10 |
| EM | COV | Log | 0.48 | -1.57 | 0.19 | 0.08 | 0.00 | 0.11 | 0.55 |
| JCU | COV | Log | 0.74 | -1.39 | 0.22 | 0.15 | 0.00 | 0.06 | 0.19 |
| EQ | COV | Log | 0.91 | -0.09 | 0.24 | 0.20 | 0.00 | 0.04 | 0.74 |
| DSTG-tft | COV | Log | -0.43 | 13.69 | 0.29 | 0.04 | 0.23 | 0.02 | 10.44 |
| UOM | COV | Log | 0.97 | 6.52 | 0.52 | 0.48 | 0.00 | 0.04 | 2.96 |
| Baseline | COV | Nat | -0.68 | 12.58 | 224.49 | 0.00 | 153.54 | 70.95 | 7.35 |
| DTSG-RV | COV | Nat | 0.61 | 13.93 | 441.52 | 249.43 | 0.00 | 192.09 | 7.65 |
| DSTG-tft | COV | Nat | -0.43 | 39.58 | 476.05 | 66.17 | 379.29 | 30.59 | 14.14 |
| EM | COV | Nat | 0.48 | 14.24 | 492.39 | 170.78 | 0.00 | 321.61 | 8.02 |
| JCU | COV | Nat | 0.74 | 14.18 | 563.11 | 363.59 | 0.00 | 199.53 | 7.76 |
| EQ | COV | Nat | 0.91 | 14.81 | 613.31 | 492.25 | 0.00 | 121.06 | 8.29 |
| UOM | COV | Nat | 0.97 | 17.67 | 1674.81 | 1488.84 | 0.00 | 185.97 | 10.58 |
| DSTG-tft | FLU | Log | 0.17 | -4.60 | 0.03 | 0.02 | 0.00 | 0.01 | -1.33 |
| EQ | FLU | Log | -0.36 | -2.56 | 0.08 | 0.00 | 0.03 | 0.05 | -0.43 |
| DTSG-RV | FLU | Log | -0.32 | -2.07 | 0.10 | 0.00 | 0.02 | 0.08 | -0.08 |
| EM | FLU | Log | -0.14 | -0.95 | 0.14 | 0.00 | 0.01 | 0.13 | -0.19 |
| UOM | FLU | Log | 0.58 | -1.01 | 0.24 | 0.18 | 0.00 | 0.06 | 0.36 |
| Baseline | FLU | Log | -0.90 | 0.77 | 0.33 | 0.00 | 0.28 | 0.05 | 1.19 |
| JCU | FLU | Log | -1.00 | 82.99 | 1.09 | 0.00 | 1.06 | 0.03 | 20.00 |
| DSTG-tft | FLU | Nat | 0.17 | 9.96 | 46.27 | 24.71 | 2.01 | 19.55 | 5.96 |
| EQ | FLU | Nat | -0.36 | 11.88 | 109.58 | 0.01 | 30.72 | 78.85 | 6.88 |
| DTSG-RV | FLU | Nat | -0.32 | 12.33 | 152.40 | 0.00 | 32.65 | 119.75 | 7.22 |
| EM | FLU | Nat | -0.14 | 13.14 | 184.43 | 1.53 | 7.67 | 175.23 | 7.06 |
| Baseline | FLU | Nat | -0.90 | 16.45 | 454.89 | 0.00 | 403.19 | 51.69 | 8.49 |
| UOM | FLU | Nat | 0.58 | 13.71 | 583.74 | 420.57 | 1.94 | 161.23 | 7.67 |
| JCU | FLU | Nat | -1.00 | 357.37 | 1040.43 | 0.00 | 1027.46 | 12.97 | 20.00 |
| EQ | RSV | Log | -0.05 | -4.16 | 0.03 | 0.00 | 0.00 | 0.03 | -1.12 |
| UOM | RSV | Log | 0.26 | -4.06 | 0.03 | 0.01 | 0.00 | 0.03 | -1.23 |
| JCU | RSV | Log | 0.42 | -3.80 | 0.05 | 0.03 | 0.00 | 0.03 | -0.99 |
| EM | RSV | Log | 0.02 | -3.16 | 0.06 | 0.00 | 0.00 | 0.05 | -0.71 |
| DTSG-RV | RSV | Log | 0.36 | -2.89 | 0.07 | 0.02 | 0.00 | 0.05 | -0.49 |
| Baseline | RSV | Log | 0.00 | -2.32 | 0.08 | 0.00 | 0.00 | 0.07 | -0.25 |
| DSTG-tft | RSV | Log | -0.97 | 29.37 | 0.26 | 0.00 | 0.25 | 0.01 | 14.71 |
| EQ | RSV | Nat | -0.05 | 10.79 | 58.34 | 1.93 | 5.61 | 50.80 | 6.34 |
| UOM | RSV | Nat | 0.26 | 11.00 | 62.12 | 14.45 | 0.00 | 47.67 | 6.24 |
| EM | RSV | Nat | 0.02 | 11.81 | 99.87 | 2.90 | 1.35 | 95.63 | 6.76 |
| JCU | RSV | Nat | 0.42 | 11.33 | 111.88 | 52.12 | 0.00 | 59.76 | 6.48 |
| Baseline | RSV | Nat | 0.00 | 12.83 | 140.91 | 6.50 | 5.48 | 128.94 | 7.21 |
| DTSG-RV | RSV | Nat | 0.36 | 12.37 | 145.38 | 39.85 | 0.00 | 105.53 | 6.98 |
| DSTG-tft | RSV | Nat | -0.97 | 60.43 | 413.97 | 0.00 | 397.67 | 16.30 | 16.51 |

Table 1: Forecast metrics for 2024-05-09 . Where EM is ensemble mixture EQ is ensemble quantile JCU is JCU-renewal UOM is UOM-seirode. Nat is for the natural scale. Op and Up are over and under prediction respectivly. Disp is dispersion and LS is the log score.

| Model | Path | Scale | Bias | Dss | Crps | Op | Up | Disp | LS |
|---|---|---|---|---|---|---|---|---|---|
| Baseline | COV | Log | -0.47 | -2.33 | 0.10 | 0.00 | 0.06 | 0.04 | -0.25 |
| DTSG-RV | COV | Log | 0.55 | -1.57 | 0.19 | 0.14 | 0.00 | 0.06 | 0.14 |
| EM | COV | Log | 0.44 | -1.58 | 0.21 | 0.09 | 0.00 | 0.12 | 0.82 |
| JCU | COV | Log | 0.72 | -1.01 | 0.27 | 0.21 | 0.00 | 0.06 | 0.39 |
| EQ | COV | Log | 0.84 | 0.36 | 0.28 | 0.24 | 0.00 | 0.04 | 0.88 |
| DSTG-tft | COV | Log | -0.50 | 20.82 | 0.29 | 0.02 | 0.25 | 0.02 | 11.93 |
| UOM | COV | Log | 0.97 | 10.72 | 0.63 | 0.59 | 0.00 | 0.04 | 10.66 |
| Baseline | COV | Nat | -0.47 | 12.64 | 189.56 | 3.60 | 108.66 | 77.30 | 7.30 |
| DTSG-RV | COV | Nat | 0.55 | 13.76 | 450.80 | 296.27 | 0.00 | 154.53 | 7.68 |
| DSTG-tft | COV | Nat | -0.50 | 48.25 | 478.89 | 49.64 | 405.58 | 23.66 | 16.07 |
| EM | COV | Nat | 0.44 | 14.33 | 542.69 | 204.52 | 0.00 | 338.17 | 8.15 |
| EQ | COV | Nat | 0.84 | 14.84 | 692.20 | 563.95 | 0.00 | 128.25 | 8.35 |
| JCU | COV | Nat | 0.72 | 14.39 | 694.52 | 480.25 | 0.00 | 214.27 | 7.96 |
| UOM | COV | Nat | 0.97 | 19.23 | 1988.57 | 1792.27 | 0.00 | 196.30 | 13.37 |
| DSTG-tft | FLU | Log | 0.22 | -4.88 | 0.03 | 0.02 | 0.00 | 0.01 | -1.53 |
| DTSG-RV | FLU | Log | 0.16 | -2.19 | 0.10 | 0.02 | 0.00 | 0.08 | -0.11 |
| EM | FLU | Log | 0.12 | -1.89 | 0.10 | 0.01 | 0.00 | 0.09 | -0.38 |
| EQ | FLU | Log | 0.50 | -2.51 | 0.13 | 0.08 | 0.00 | 0.04 | -0.40 |
| Baseline | FLU | Log | -0.59 | -1.54 | 0.18 | 0.00 | 0.13 | 0.05 | 0.14 |
| JCU | FLU | Log | -0.90 | 1.55 | 0.22 | 0.00 | 0.18 | 0.04 | 2.02 |
| UOM | FLU | Log | 0.99 | 7.65 | 0.55 | 0.50 | 0.00 | 0.05 | 7.93 |
| DSTG-tft | FLU | Nat | 0.22 | 10.07 | 58.53 | 33.65 | 3.40 | 21.48 | 5.94 |
| DTSG-RV | FLU | Nat | 0.16 | 13.19 | 220.86 | 33.28 | 0.06 | 187.52 | 7.37 |
| EM | FLU | Nat | 0.12 | 13.70 | 248.12 | 17.41 | 0.81 | 229.90 | 7.20 |
| Baseline | FLU | Nat | -0.59 | 13.14 | 304.91 | 0.22 | 228.77 | 75.93 | 7.61 |
| EQ | FLU | Nat | 0.50 | 12.66 | 318.40 | 202.42 | 0.00 | 115.99 | 7.09 |
| JCU | FLU | Nat | -0.90 | 17.56 | 340.69 | 0.00 | 289.00 | 51.69 | 9.59 |
| UOM | FLU | Nat | 0.99 | 17.92 | 1769.33 | 1558.38 | 0.00 | 210.95 | 12.46 |
| Baseline | RSV | Log | -0.06 | -2.26 | 0.10 | 0.02 | 0.01 | 0.07 | -0.22 |
| EM | RSV | Log | 0.03 | -2.74 | 0.10 | 0.04 | 0.00 | 0.05 | -0.52 |
| UOM | RSV | Log | 0.24 | -1.97 | 0.11 | 0.09 | 0.00 | 0.03 | -0.37 |
| EQ | RSV | Log | -0.01 | -1.87 | 0.11 | 0.08 | 0.01 | 0.03 | -0.17 |
| DTSG-RV | RSV | Log | 0.42 | -1.60 | 0.14 | 0.11 | 0.00 | 0.03 | 0.48 |
| JCU | RSV | Log | 0.44 | -0.88 | 0.17 | 0.14 | 0.00 | 0.03 | 1.29 |
| DSTG-tft | RSV | Log | -0.99 | 23.69 | 0.23 | 0.00 | 0.22 | 0.01 | 15.00 |
| Baseline | RSV | Nat | -0.06 | 12.89 | 179.02 | 33.39 | 16.36 | 129.27 | 7.26 |
| EM | RSV | Nat | 0.03 | 12.28 | 191.05 | 81.50 | 5.01 | 104.54 | 6.91 |
| EQ | RSV | Nat | -0.01 | 12.52 | 203.66 | 138.62 | 14.99 | 50.05 | 7.33 |
| UOM | RSV | Nat | 0.24 | 12.55 | 208.58 | 155.46 | 1.24 | 51.88 | 7.08 |
| DTSG-RV | RSV | Nat | 0.42 | 12.78 | 286.54 | 218.55 | 0.12 | 67.87 | 7.57 |
| JCU | RSV | Nat | 0.44 | 13.17 | 339.50 | 269.16 | 0.67 | 69.68 | 8.03 |
| DSTG-tft | RSV | Nat | -0.99 | 50.04 | 378.83 | 0.00 | 361.75 | 17.07 | 16.83 |

Table 2: Forecast metrics for 2024-05-16 . Where EM is ensemble mixture EQ is ensemble quantile JCU is JCU-renewal UOM is UOM-seirode. Nat is for the natural scale. Op and Up are over and under prediction respectivly. Disp is dispersion and LS is the log score.

| Model | Path | Scale | Bias | Dss | Crps | Op | Up | Disp | LS |
|---|---|---|---|---|---|---|---|---|---|
| Baseline | COV | Log | 0.30 | -2.10 | 0.14 | 0.09 | 0.01 | 0.04 | -0.11 |
| DSTG-tft | COV | Log | -0.97 | 16.44 | 0.27 | 0.00 | 0.25 | 0.02 | 11.43 |
| EM | COV | Log | 0.38 | -1.25 | 0.27 | 0.14 | 0.00 | 0.13 | 1.17 |
| JCU | COV | Log | 0.70 | -0.35 | 0.33 | 0.27 | 0.00 | 0.06 | 0.71 |
| DTSG-RV | COV | Log | 0.78 | 0.89 | 0.36 | 0.30 | 0.00 | 0.06 | 1.34 |
| EQ | COV | Log | 0.82 | 2.78 | 0.38 | 0.34 | 0.00 | 0.04 | 1.90 |
| UOM | COV | Log | 0.99 | 21.83 | 0.77 | 0.74 | 0.00 | 0.04 | 12.29 |
| Baseline | COV | Nat | 0.30 | 12.90 | 256.10 | 153.14 | 10.32 | 92.64 | 7.35 |
| DSTG-tft | COV | Nat | -0.97 | 40.68 | 391.95 | 0.00 | 370.78 | 21.17 | 15.56 |
| EM | COV | Nat | 0.38 | 14.59 | 644.65 | 292.03 | 0.00 | 352.62 | 8.28 |
| JCU | COV | Nat | 0.70 | 14.56 | 743.13 | 543.36 | 0.00 | 199.76 | 8.17 |
| DTSG-RV | COV | Nat | 0.78 | 15.13 | 798.38 | 631.07 | 0.00 | 167.31 | 8.68 |
| EQ | COV | Nat | 0.82 | 15.90 | 844.34 | 715.86 | 0.00 | 128.48 | 9.04 |
| UOM | COV | Nat | 0.99 | 23.03 | 2233.20 | 2060.12 | 0.00 | 173.08 | 15.73 |
| DSTG-tft | FLU | Log | 0.06 | -4.89 | 0.03 | 0.02 | 0.00 | 0.01 | -1.53 |
| DTSG-RV | FLU | Log | 0.05 | -2.37 | 0.09 | 0.02 | 0.00 | 0.07 | -0.26 |
| EQ | FLU | Log | -0.01 | -2.26 | 0.11 | 0.03 | 0.02 | 0.06 | -0.21 |
| Baseline | FLU | Log | -0.73 | -1.67 | 0.17 | 0.00 | 0.11 | 0.05 | 0.09 |
| EM | FLU | Log | 0.01 | -0.43 | 0.17 | 0.01 | 0.00 | 0.16 | -0.05 |
| UOM | FLU | Log | 0.95 | 7.74 | 0.51 | 0.47 | 0.00 | 0.04 | 10.63 |
| JCU | FLU | Log | -1.00 | 186.56 | 1.40 | 0.00 | 1.37 | 0.03 | 20.00 |
| DSTG-tft | FLU | Nat | 0.06 | 10.36 | 63.48 | 35.93 | 4.73 | 22.82 | 6.12 |
| DTSG-RV | FLU | Nat | 0.05 | 13.22 | 217.52 | 40.64 | 9.11 | 167.77 | 7.40 |
| EQ | FLU | Nat | -0.01 | 13.12 | 257.82 | 80.02 | 32.69 | 145.12 | 7.44 |
| Baseline | FLU | Nat | -0.73 | 13.21 | 318.16 | 0.00 | 228.24 | 89.92 | 7.74 |
| EM | FLU | Nat | 0.01 | 14.46 | 330.31 | 12.97 | 1.28 | 316.06 | 7.52 |
| JCU | FLU | Nat | -1.00 | 911.07 | 1602.33 | 0.00 | 1589.49 | 12.84 | 20.00 |
| UOM | FLU | Nat | 0.95 | 17.84 | 1806.65 | 1607.73 | 0.00 | 198.92 | 14.34 |
| Baseline | RSV | Log | 0.56 | -1.34 | 0.21 | 0.14 | 0.00 | 0.07 | 0.25 |
| EM | RSV | Log | 0.37 | -1.61 | 0.22 | 0.14 | 0.00 | 0.09 | 1.59 |
| UOM | RSV | Log | 0.67 | 3.95 | 0.26 | 0.23 | 0.00 | 0.02 | 2.00 |
| EQ | RSV | Log | 0.55 | 1.76 | 0.26 | 0.23 | 0.00 | 0.03 | 1.08 |
| DSTG-tft | RSV | Log | -1.00 | 37.17 | 0.33 | 0.00 | 0.31 | 0.01 | 20.00 |
| JCU | RSV | Log | 0.82 | 6.93 | 0.37 | 0.34 | 0.00 | 0.03 | 9.57 |
| DTSG-RV | RSV | Log | 1.00 | 25.88 | 0.53 | 0.50 | 0.00 | 0.02 | 12.82 |
| Baseline | RSV | Nat | 0.56 | 13.94 | 410.61 | 248.03 | 0.00 | 162.58 | 7.70 |
| EM | RSV | Nat | 0.37 | 13.70 | 454.75 | 261.19 | 0.00 | 193.56 | 8.27 |
| DSTG-tft | RSV | Nat | -1.00 | 74.61 | 484.07 | 0.00 | 468.18 | 15.89 | 20.00 |
| UOM | RSV | Nat | 0.67 | 16.48 | 490.68 | 432.50 | 0.00 | 58.18 | 9.00 |
| EQ | RSV | Nat | 0.55 | 15.08 | 497.51 | 419.76 | 0.33 | 77.42 | 8.49 |
| JCU | RSV | Nat | 0.82 | 17.23 | 779.38 | 692.69 | 0.00 | 86.69 | 12.91 |
| DTSG-RV | RSV | Nat | 1.00 | 25.57 | 1231.63 | 1147.52 | 0.00 | 84.11 | 15.92 |

Table 3: Forecast metrics for 2024-05-23 . Where EM is ensemble mixture EQ is ensemble quantile JCU is JCU-renewal UOM is UOM-seirode. Nat is for the natural scale. Op and Up are over and under prediction respectivly. Disp is dispersion and LS is the log score.

| Model | Path | Scale | Bias | Dss | Crps | Op | Up | Disp | LS |
|---|---|---|---|---|---|---|---|---|---|
| DSTG-tft | COV | Log | -0.97 | 7.70 | 0.21 | 0.00 | 0.19 | 0.02 | 9.84 |
| Baseline | COV | Log | 0.85 | 0.46 | 0.28 | 0.23 | 0.00 | 0.04 | 1.14 |
| EM | COV | Log | 0.49 | -0.77 | 0.32 | 0.20 | 0.00 | 0.12 | 1.72 |
| EQ | COV | Log | 0.96 | 5.41 | 0.46 | 0.42 | 0.00 | 0.04 | 2.94 |
| DTSG-RV | COV | Log | 0.95 | 3.44 | 0.47 | 0.42 | 0.00 | 0.05 | 2.33 |
| JCU | COV | Log | 0.96 | 2.33 | 0.49 | 0.42 | 0.00 | 0.06 | 1.87 |
| UOM | COV | Log | 1.00 | 12.31 | 0.74 | 0.69 | 0.00 | 0.06 | 12.85 |
| DSTG-tft | COV | Nat | -0.97 | 26.09 | 277.36 | 0.00 | 256.53 | 20.83 | 14.25 |
| Baseline | COV | Nat | 0.85 | 14.68 | 508.96 | 407.02 | 0.00 | 101.94 | 8.47 |
| EM | COV | Nat | 0.49 | 14.96 | 709.18 | 403.63 | 0.00 | 305.55 | 8.47 |
| EQ | COV | Nat | 0.96 | 17.58 | 943.04 | 822.41 | 0.00 | 120.64 | 10.11 |
| DTSG-RV | COV | Nat | 0.95 | 16.50 | 975.15 | 819.64 | 0.00 | 155.51 | 9.63 |
| JCU | COV | Nat | 0.96 | 16.16 | 1067.52 | 855.47 | 0.00 | 212.05 | 9.21 |
| UOM | COV | Nat | 1.00 | 20.08 | 1816.93 | 1580.86 | 0.00 | 236.07 | 14.80 |
| JCU | FLU | Log | 0.11 | -3.17 | 0.07 | 0.02 | 0.01 | 0.04 | -0.70 |
| DTSG-RV | FLU | Log | 0.20 | -2.09 | 0.10 | 0.02 | 0.00 | 0.08 | -0.09 |
| EM | FLU | Log | 0.26 | -1.95 | 0.11 | 0.02 | 0.00 | 0.09 | -0.35 |
| Baseline | FLU | Log | -0.47 | -2.37 | 0.12 | 0.00 | 0.07 | 0.05 | -0.23 |
| DSTG-tft | FLU | Log | -0.27 | 29.80 | 0.15 | 0.01 | 0.13 | 0.01 | 3.98 |
| EQ | FLU | Log | 0.67 | -1.56 | 0.15 | 0.11 | 0.00 | 0.04 | 0.11 |
| UOM | FLU | Log | 0.99 | 12.36 | 0.54 | 0.50 | 0.00 | 0.04 | 15.15 |
| JCU | FLU | Nat | 0.11 | 12.59 | 171.95 | 50.23 | 16.18 | 105.54 | 7.13 |
| Baseline | FLU | Nat | -0.47 | 13.00 | 295.28 | 0.00 | 182.99 | 112.29 | 7.58 |
| DTSG-RV | FLU | Nat | 0.20 | 14.02 | 309.22 | 56.09 | 2.12 | 251.02 | 7.73 |
| EM | FLU | Nat | 0.26 | 14.30 | 369.93 | 48.92 | 0.01 | 321.00 | 7.51 |
| DSTG-tft | FLU | Nat | -0.27 | 68.43 | 385.92 | 28.00 | 335.09 | 22.83 | 9.77 |
| EQ | FLU | Nat | 0.67 | 14.12 | 442.50 | 305.94 | 0.00 | 136.56 | 7.95 |
| UOM | FLU | Nat | 0.99 | 20.69 | 2109.76 | 1910.58 | 0.00 | 199.18 | 17.05 |
| EM | RSV | Log | 0.45 | -1.11 | 0.30 | 0.21 | 0.00 | 0.10 | 3.61 |
| DSTG-tft | RSV | Log | -1.00 | 32.44 | 0.33 | 0.00 | 0.32 | 0.01 | 20.00 |
| EQ | RSV | Log | 0.75 | 3.44 | 0.36 | 0.32 | 0.00 | 0.04 | 1.69 |
| Baseline | RSV | Log | 0.84 | 0.60 | 0.37 | 0.31 | 0.00 | 0.07 | 1.21 |
| UOM | RSV | Log | 0.91 | 13.18 | 0.44 | 0.41 | 0.00 | 0.03 | 13.82 |
| DTSG-RV | RSV | Log | 0.92 | 5.05 | 0.52 | 0.47 | 0.00 | 0.05 | 5.57 |
| JCU | RSV | Log | 0.96 | 22.38 | 0.58 | 0.55 | 0.00 | 0.03 | 14.85 |
| DSTG-tft | RSV | Nat | -1.00 | 63.80 | 431.81 | 0.00 | 416.92 | 14.90 | 20.00 |
| EM | RSV | Nat | 0.45 | 14.31 | 597.30 | 390.46 | 0.00 | 206.85 | 9.23 |
| EQ | RSV | Nat | 0.75 | 16.32 | 654.77 | 560.80 | 0.00 | 93.97 | 9.03 |
| Baseline | RSV | Nat | 0.84 | 15.10 | 723.96 | 545.28 | 0.00 | 178.67 | 8.52 |
| UOM | RSV | Nat | 0.91 | 21.81 | 838.05 | 767.64 | 0.00 | 70.41 | 15.50 |
| DTSG-RV | RSV | Nat | 0.92 | 16.95 | 1078.12 | 915.72 | 0.00 | 162.40 | 10.91 |
| JCU | RSV | Nat | 0.96 | 24.58 | 1220.95 | 1128.60 | 0.00 | 92.35 | 16.75 |

Table 4: Forecast metrics for 2024-05-30 . Where EM is ensemble mixture EQ is ensemble quantile JCU is JCU-renewal UOM is UOM-seirode. Nat is for the natural scale. Op and Up are over and under prediction respectivly. Disp is dispersion and LS is the log score.

| Model | Path | Scale | Bias | Dss | Crps | Op | Up | Disp | LS |
|---|---|---|---|---|---|---|---|---|---|
| DSTG-tft | COV | Log | 0.01 | 25.22 | 0.19 | 0.09 | 0.09 | 0.01 | 13.52 |
| EM | COV | Log | 0.66 | -0.73 | 0.22 | 0.13 | 0.00 | 0.08 | 0.87 |
| DTSG-RV | COV | Log | 0.79 | -0.98 | 0.22 | 0.15 | 0.00 | 0.06 | 0.34 |
| JCU | COV | Log | 0.84 | -0.39 | 0.26 | 0.19 | 0.00 | 0.07 | 0.73 |
| EQ | COV | Log | 0.98 | 4.37 | 0.29 | 0.26 | 0.00 | 0.03 | 6.28 |
| Baseline | COV | Log | 0.98 | 2.95 | 0.37 | 0.33 | 0.00 | 0.04 | 2.30 |
| UOM | COV | Log | 0.99 | 3.31 | 0.45 | 0.40 | 0.00 | 0.05 | 3.14 |
| DSTG-tft | COV | Nat | 0.01 | 38.24 | 264.25 | 152.98 | 98.21 | 13.05 | 16.66 |
| DTSG-RV | COV | Nat | 0.79 | 14.00 | 365.50 | 237.03 | 0.00 | 128.47 | 7.63 |
| EM | COV | Nat | 0.66 | 14.23 | 369.31 | 210.47 | 0.00 | 158.84 | 8.02 |
| JCU | COV | Nat | 0.84 | 14.48 | 447.43 | 308.17 | 0.00 | 139.26 | 7.97 |
| EQ | COV | Nat | 0.98 | 17.17 | 489.37 | 419.67 | 0.00 | 69.71 | 11.66 |
| Baseline | COV | Nat | 0.98 | 16.18 | 638.71 | 541.77 | 0.00 | 96.94 | 9.49 |
| UOM | COV | Nat | 0.99 | 16.41 | 827.81 | 695.24 | 0.00 | 132.56 | 10.24 |
| DTSG-RV | FLU | Log | 0.29 | -2.02 | 0.10 | 0.02 | 0.00 | 0.07 | -0.12 |
| EQ | FLU | Log | 0.83 | -0.72 | 0.17 | 0.13 | 0.00 | 0.04 | 0.56 |
| EM | FLU | Log | 0.42 | -1.43 | 0.17 | 0.05 | 0.00 | 0.12 | 0.51 |
| Baseline | FLU | Log | -0.59 | -1.50 | 0.19 | 0.00 | 0.14 | 0.05 | 0.19 |
| JCU | FLU | Log | 0.95 | 2.34 | 0.25 | 0.21 | 0.00 | 0.04 | 2.07 |
| DSTG-tft | FLU | Log | -0.56 | 89.82 | 0.34 | 0.01 | 0.32 | 0.01 | 9.92 |
| UOM | FLU | Log | 1.00 | 12.76 | 0.52 | 0.49 | 0.00 | 0.03 | 17.13 |
| DTSG-RV | FLU | Nat | 0.29 | 14.38 | 323.16 | 59.86 | 0.11 | 263.19 | 7.87 |
| EQ | FLU | Nat | 0.83 | 15.28 | 547.31 | 385.11 | 0.00 | 162.20 | 8.54 |
| Baseline | FLU | Nat | -0.59 | 14.31 | 557.90 | 0.00 | 438.29 | 119.61 | 8.18 |
| EM | FLU | Nat | 0.42 | 15.00 | 609.06 | 155.83 | 0.00 | 453.22 | 8.46 |
| JCU | FLU | Nat | 0.95 | 17.56 | 888.62 | 710.22 | 0.00 | 178.40 | 9.97 |
| DSTG-tft | FLU | Nat | -0.56 | 207.33 | 934.35 | 12.28 | 899.42 | 22.65 | 13.80 |
| UOM | FLU | Nat | 1.00 | 21.97 | 2213.96 | 2012.86 | 0.00 | 201.11 | 18.58 |
| DSTG-tft | RSV | Log | -1.00 | 13.47 | 0.21 | 0.00 | 0.20 | 0.01 | 11.31 |
| EM | RSV | Log | 0.50 | -0.47 | 0.38 | 0.28 | 0.00 | 0.09 | 2.98 |
| Baseline | RSV | Log | 0.96 | 1.56 | 0.43 | 0.37 | 0.00 | 0.07 | 1.72 |
| EQ | RSV | Log | 0.99 | 6.70 | 0.47 | 0.43 | 0.00 | 0.04 | 3.11 |
| UOM | RSV | Log | 1.00 | 20.50 | 0.58 | 0.55 | 0.00 | 0.03 | 18.85 |
| JCU | RSV | Log | 1.00 | 30.22 | 0.67 | 0.64 | 0.00 | 0.03 | 20.00 |
| DTSG-RV | RSV | Log | 1.00 | 18.24 | 0.72 | 0.68 | 0.00 | 0.04 | 14.30 |
| DSTG-tft | RSV | Nat | -1.00 | 33.51 | 238.07 | 0.00 | 223.99 | 14.08 | 14.92 |
| EM | RSV | Nat | 0.50 | 14.87 | 697.15 | 500.31 | 0.00 | 196.85 | 8.80 |
| Baseline | RSV | Nat | 0.96 | 15.48 | 753.85 | 592.45 | 0.00 | 161.40 | 8.84 |
| EQ | RSV | Nat | 0.99 | 18.31 | 793.05 | 698.81 | 0.00 | 94.24 | 10.20 |
| UOM | RSV | Nat | 1.00 | 25.98 | 1041.20 | 962.55 | 0.00 | 78.65 | 19.25 |
| JCU | RSV | Nat | 1.00 | 28.99 | 1263.53 | 1179.88 | 0.00 | 83.65 | 20.00 |
| DTSG-RV | RSV | Nat | 1.00 | 22.54 | 1422.12 | 1299.30 | 0.00 | 122.81 | 16.06 |

Table 5: Forecast metrics for 2024-06-06 . Where EM is ensemble mixture EQ is ensemble quantile JCU is JCU-renewal UOM is UOM-seirode. Nat is for the natural scale. Op and Up are over and under prediction respectivly. Disp is dispersion and LS is the log score.

| Model | Path | Scale | Bias | Dss | Crps | Op | Up | Disp | LS |
|---|---|---|---|---|---|---|---|---|---|
| EQ | COV | Log | 0.19 | -3.13 | 0.06 | 0.03 | 0.00 | 0.03 | -0.71 |
| JCU | COV | Log | 0.25 | -2.54 | 0.08 | 0.02 | 0.00 | 0.06 | -0.37 |
| DTSG-RV | COV | Log | -0.14 | -2.41 | 0.09 | 0.00 | 0.02 | 0.07 | -0.26 |
| EM | COV | Log | 0.07 | -2.47 | 0.09 | 0.02 | 0.01 | 0.06 | 0.01 |
| UOM | COV | Log | 0.67 | -1.47 | 0.14 | 0.10 | 0.00 | 0.04 | 0.24 |
| DSTG-tft | COV | Log | -0.49 | 17.22 | 0.20 | 0.03 | 0.16 | 0.01 | 15.23 |
| Baseline | COV | Log | 0.97 | 1.37 | 0.30 | 0.26 | 0.00 | 0.04 | 1.59 |
| EQ | COV | Nat | 0.19 | 11.07 | 80.35 | 42.69 | 2.76 | 34.90 | 6.42 |
| DTSG-RV | COV | Nat | -0.14 | 11.81 | 99.58 | 5.12 | 18.84 | 75.61 | 6.87 |
| JCU | COV | Nat | 0.25 | 12.01 | 100.91 | 21.26 | 0.00 | 79.65 | 6.77 |
| EM | COV | Nat | 0.07 | 11.95 | 113.80 | 29.26 | 7.36 | 77.17 | 7.14 |
| UOM | COV | Nat | 0.67 | 12.91 | 204.71 | 149.61 | 0.00 | 55.09 | 7.38 |
| DSTG-tft | COV | Nat | -0.49 | 36.61 | 220.33 | 46.58 | 162.27 | 11.48 | 17.04 |
| Baseline | COV | Nat | 0.97 | 15.05 | 454.14 | 374.19 | 0.00 | 79.95 | 8.69 |
| JCU | FLU | Log | 0.11 | -3.53 | 0.05 | 0.01 | 0.00 | 0.04 | -0.84 |
| EQ | FLU | Log | -0.09 | -3.42 | 0.05 | 0.00 | 0.01 | 0.04 | -0.78 |
| EM | FLU | Log | -0.15 | -1.84 | 0.13 | 0.00 | 0.01 | 0.11 | 0.21 |
| DTSG-RV | FLU | Log | -0.74 | -0.80 | 0.27 | 0.00 | 0.18 | 0.08 | 0.53 |
| Baseline | FLU | Log | -0.93 | 0.76 | 0.32 | 0.00 | 0.27 | 0.05 | 1.24 |
| UOM | FLU | Log | 1.00 | 5.04 | 0.34 | 0.31 | 0.00 | 0.03 | 4.10 |
| DSTG-tft | FLU | Log | -0.98 | 78.13 | 0.38 | 0.00 | 0.36 | 0.01 | 14.88 |
| EQ | FLU | Nat | -0.09 | 12.80 | 181.78 | 10.36 | 29.94 | 141.47 | 7.38 |
| JCU | FLU | Nat | 0.11 | 12.86 | 183.16 | 29.33 | 11.13 | 142.69 | 7.32 |
| EM | FLU | Nat | -0.15 | 14.38 | 438.30 | 0.43 | 51.93 | 385.94 | 8.35 |
| DTSG-RV | FLU | Nat | -0.74 | 14.92 | 797.12 | 0.00 | 602.73 | 194.39 | 8.69 |
| Baseline | FLU | Nat | -0.93 | 17.76 | 968.06 | 0.00 | 852.35 | 115.71 | 9.43 |
| DSTG-tft | FLU | Nat | -0.98 | 170.77 | 1150.14 | 0.00 | 1124.22 | 25.92 | 16.84 |
| UOM | FLU | Nat | 1.00 | 18.90 | 1502.39 | 1337.91 | 0.00 | 164.48 | 12.20 |
| EM | RSV | Log | 0.46 | -1.48 | 0.22 | 0.12 | 0.00 | 0.10 | 1.55 |
| DTSG-RV | RSV | Log | 0.83 | -0.78 | 0.24 | 0.18 | 0.00 | 0.06 | 0.44 |
| EQ | RSV | Log | 0.91 | 0.44 | 0.24 | 0.20 | 0.00 | 0.04 | 0.92 |
| DSTG-tft | RSV | Log | -1.00 | 23.14 | 0.26 | 0.00 | 0.25 | 0.01 | 18.63 |
| Baseline | RSV | Log | 0.82 | -0.18 | 0.31 | 0.25 | 0.00 | 0.06 | 0.80 |
| UOM | RSV | Log | 0.99 | 4.55 | 0.35 | 0.32 | 0.00 | 0.04 | 2.74 |
| JCU | RSV | Log | 1.00 | 22.36 | 0.52 | 0.50 | 0.00 | 0.03 | 20.00 |
| DSTG-tft | RSV | Nat | -1.00 | 47.80 | 267.55 | 0.00 | 256.34 | 11.21 | 20.00 |
| EM | RSV | Nat | 0.46 | 13.23 | 298.99 | 153.87 | 0.00 | 145.12 | 8.40 |
| EQ | RSV | Nat | 0.91 | 14.20 | 307.76 | 250.00 | 0.00 | 57.76 | 7.99 |
| DTSG-RV | RSV | Nat | 0.83 | 13.65 | 317.66 | 222.84 | 0.00 | 94.82 | 7.49 |
| Baseline | RSV | Nat | 0.82 | 13.98 | 436.68 | 317.46 | 0.00 | 119.22 | 7.85 |
| UOM | RSV | Nat | 0.99 | 17.12 | 491.76 | 421.47 | 0.00 | 70.29 | 9.72 |
| JCU | RSV | Nat | 1.00 | 26.90 | 781.18 | 725.76 | 0.00 | 55.42 | 20.00 |

Table 6: Forecast metrics for 2024-06-13 . Where EM is ensemble mixture EQ is ensemble quantile JCU is JCU-renewal UOM is UOM-seirode. Nat is for the natural scale. Op and Up are over and under prediction respectivly. Disp is dispersion and LS is the log score.

| Model | Path | Scale | Bias | Dss | Crps | Op | Up | Disp | LS |
|---|---|---|---|---|---|---|---|---|---|
| DTSG-RV | COV | Log | 0.00 | -3.45 | 0.05 | 0.00 | 0.01 | 0.04 | -0.75 |
| UOM | COV | Log | -0.23 | -3.20 | 0.06 | 0.00 | 0.02 | 0.04 | -0.71 |
| EQ | COV | Log | -0.76 | -2.37 | 0.10 | 0.00 | 0.08 | 0.03 | -0.34 |
| EM | COV | Log | -0.47 | -2.46 | 0.11 | 0.00 | 0.05 | 0.06 | -0.26 |
| JCU | COV | Log | -0.66 | -1.75 | 0.16 | 0.00 | 0.09 | 0.07 | 0.02 |
| Baseline | COV | Log | 0.75 | -1.55 | 0.17 | 0.13 | 0.00 | 0.04 | 0.19 |
| DSTG-tft | COV | Log | -1.00 | 29.75 | 0.29 | 0.00 | 0.28 | 0.01 | 15.78 |
| DTSG-RV | COV | Nat | 0.00 | 10.76 | 58.03 | 4.84 | 6.18 | 47.01 | 6.32 |
| UOM | COV | Nat | -0.23 | 10.77 | 69.01 | 2.92 | 19.34 | 46.75 | 6.36 |
| EQ | COV | Nat | -0.76 | 11.75 | 106.72 | 0.00 | 80.83 | 25.89 | 6.72 |
| EM | COV | Nat | -0.47 | 11.33 | 109.27 | 0.00 | 51.66 | 57.60 | 6.80 |
| JCU | COV | Nat | -0.66 | 11.88 | 154.18 | 0.00 | 96.78 | 57.41 | 7.07 |
| Baseline | COV | Nat | 0.75 | 12.73 | 227.84 | 163.05 | 0.00 | 64.79 | 7.25 |
| DSTG-tft | COV | Nat | -1.00 | 57.11 | 291.27 | 0.00 | 281.05 | 10.21 | 17.59 |
| EQ | FLU | Log | -0.55 | -2.71 | 0.08 | 0.00 | 0.05 | 0.04 | -0.48 |
| JCU | FLU | Log | -0.48 | -1.96 | 0.10 | 0.01 | 0.06 | 0.03 | -0.08 |
| EM | FLU | Log | -0.31 | -1.75 | 0.15 | 0.00 | 0.04 | 0.11 | 0.44 |
| DTSG-RV | FLU | Log | -0.73 | -1.07 | 0.20 | 0.00 | 0.13 | 0.07 | 0.37 |
| UOM | FLU | Log | 0.95 | 0.48 | 0.21 | 0.18 | 0.00 | 0.03 | 2.33 |
| Baseline | FLU | Log | -0.89 | 0.05 | 0.25 | 0.00 | 0.20 | 0.05 | 0.93 |
| DSTG-tft | FLU | Log | -1.00 | 148.79 | 0.60 | 0.00 | 0.58 | 0.01 | 20.00 |
| EQ | FLU | Nat | -0.55 | 13.62 | 307.64 | 0.00 | 173.91 | 133.73 | 7.79 |
| JCU | FLU | Nat | -0.48 | 14.69 | 376.39 | 33.29 | 215.47 | 127.62 | 8.19 |
| EM | FLU | Nat | -0.31 | 14.43 | 499.18 | 0.00 | 137.17 | 362.01 | 8.71 |
| DTSG-RV | FLU | Nat | -0.73 | 15.09 | 661.18 | 0.00 | 465.75 | 195.43 | 8.63 |
| Baseline | FLU | Nat | -0.89 | 16.70 | 832.95 | 0.00 | 696.24 | 136.70 | 9.20 |
| UOM | FLU | Nat | 0.95 | 16.21 | 993.15 | 840.75 | 0.00 | 152.40 | 10.24 |
| DSTG-tft | FLU | Nat | -1.00 | 380.76 | 1702.93 | 0.00 | 1678.16 | 24.77 | 20.00 |
| DTSG-RV | RSV | Log | 0.56 | -2.35 | 0.12 | 0.06 | 0.00 | 0.05 | -0.32 |
| EM | RSV | Log | 0.37 | -1.76 | 0.15 | 0.05 | 0.00 | 0.10 | 0.38 |
| DSTG-tft | RSV | Log | -0.99 | 8.57 | 0.17 | 0.00 | 0.15 | 0.01 | 10.65 |
| UOM | RSV | Log | 0.93 | -0.24 | 0.19 | 0.16 | 0.00 | 0.04 | 0.55 |
| EQ | RSV | Log | 0.86 | -0.58 | 0.20 | 0.16 | 0.00 | 0.04 | 0.53 |
| Baseline | RSV | Log | 0.89 | 1.08 | 0.39 | 0.33 | 0.00 | 0.06 | 1.53 |
| JCU | RSV | Log | 1.00 | 35.45 | 0.59 | 0.57 | 0.00 | 0.02 | 20.00 |
| DTSG-RV | RSV | Nat | 0.56 | 11.84 | 121.33 | 60.47 | 0.00 | 60.86 | 6.59 |
| DSTG-tft | RSV | Nat | -0.99 | 25.63 | 161.87 | 0.00 | 151.08 | 10.79 | 14.00 |
| EM | RSV | Nat | 0.37 | 12.63 | 170.43 | 51.86 | 0.00 | 118.58 | 7.19 |
| EQ | RSV | Nat | 0.86 | 13.09 | 208.90 | 162.40 | 0.00 | 46.50 | 7.44 |
| UOM | RSV | Nat | 0.93 | 13.48 | 212.86 | 165.96 | 0.00 | 46.90 | 7.44 |
| Baseline | RSV | Nat | 0.89 | 14.33 | 477.57 | 371.95 | 0.00 | 105.62 | 8.35 |
| JCU | RSV | Nat | 1.00 | 32.52 | 781.69 | 735.64 | 0.00 | 46.05 | 20.00 |

Table 7: Forecast metrics for 2024-06-20 . Where EM is ensemble mixture EQ is ensemble quantile JCU is JCU-renewal UOM is UOM-seirode. Nat is for the natural scale. Op and Up are over and under prediction respectivly. Disp is dispersion and LS is the log score.

| Model | Path | Scale | Bias | Dss | Crps | Op | Up | Disp | LS |
|---|---|---|---|---|---|---|---|---|---|
| JCU | COV | Log | 0.05 | -2.75 | 0.07 | 0.00 | 0.00 | 0.06 | -0.47 |
| EQ | COV | Log | -0.66 | -3.37 | 0.07 | 0.00 | 0.04 | 0.03 | -0.78 |
| UOM | COV | Log | -0.66 | -2.86 | 0.08 | 0.00 | 0.05 | 0.03 | -0.67 |
| EM | COV | Log | -0.55 | -2.81 | 0.09 | 0.00 | 0.05 | 0.04 | -0.31 |
| DTSG-RV | COV | Log | -0.60 | -2.30 | 0.12 | 0.00 | 0.06 | 0.05 | -0.21 |
| DSTG-tft | COV | Log | -0.99 | 7.30 | 0.16 | 0.00 | 0.14 | 0.01 | 2.66 |
| Baseline | COV | Log | 0.84 | -0.60 | 0.22 | 0.18 | 0.00 | 0.04 | 0.59 |
| EQ | COV | Nat | -0.66 | 10.38 | 66.57 | 0.00 | 43.21 | 23.36 | 6.17 |
| JCU | COV | Nat | 0.05 | 11.42 | 71.46 | 3.10 | 0.49 | 67.87 | 6.48 |
| UOM | COV | Nat | -0.66 | 10.91 | 75.04 | 0.00 | 45.00 | 30.05 | 6.29 |
| EM | COV | Nat | -0.55 | 11.01 | 82.49 | 0.00 | 45.58 | 36.91 | 6.64 |
| DTSG-RV | COV | Nat | -0.60 | 11.22 | 106.89 | 0.00 | 61.30 | 45.59 | 6.74 |
| DSTG-tft | COV | Nat | -0.99 | 23.66 | 150.17 | 0.00 | 139.82 | 10.35 | 9.90 |
| Baseline | COV | Nat | 0.84 | 13.15 | 260.37 | 200.82 | 0.00 | 59.54 | 7.54 |
| EQ | FLU | Log | 0.18 | -2.96 | 0.09 | 0.05 | 0.01 | 0.04 | -0.51 |
| Baseline | FLU | Log | -0.44 | -2.22 | 0.10 | 0.00 | 0.05 | 0.05 | -0.19 |
| DTSG-RV | FLU | Log | 0.31 | -1.90 | 0.15 | 0.08 | 0.00 | 0.07 | -0.03 |
| EM | FLU | Log | 0.18 | -1.69 | 0.16 | 0.05 | 0.00 | 0.11 | 0.45 |
| JCU | FLU | Log | 0.53 | -1.10 | 0.20 | 0.16 | 0.00 | 0.04 | 0.34 |
| UOM | FLU | Log | 0.89 | 1.05 | 0.22 | 0.19 | 0.00 | 0.03 | 2.20 |
| DSTG-tft | FLU | Log | -1.00 | 207.02 | 0.73 | 0.00 | 0.72 | 0.01 | 20.00 |
| Baseline | FLU | Nat | -0.44 | 14.24 | 382.71 | 2.92 | 203.38 | 176.41 | 8.08 |
| EQ | FLU | Nat | 0.18 | 13.75 | 393.33 | 194.80 | 22.38 | 176.15 | 7.77 |
| EM | FLU | Nat | 0.18 | 14.83 | 626.82 | 206.86 | 6.82 | 413.13 | 8.79 |
| DTSG-RV | FLU | Nat | 0.31 | 15.11 | 715.40 | 326.72 | 8.86 | 379.82 | 8.26 |
| JCU | FLU | Nat | 0.53 | 15.17 | 953.55 | 730.89 | 6.74 | 215.92 | 8.61 |
| UOM | FLU | Nat | 0.89 | 16.40 | 975.19 | 829.12 | 0.00 | 146.07 | 10.42 |
| DSTG-tft | FLU | Nat | -1.00 | 657.27 | 1889.57 | 0.00 | 1866.07 | 23.50 | 20.00 |
| DSTG-tft | RSV | Log | -0.27 | -4.69 | 0.03 | 0.00 | 0.02 | 0.01 | -1.38 |
| EM | RSV | Log | 0.58 | -2.52 | 0.10 | 0.05 | 0.00 | 0.05 | -0.63 |
| DTSG-RV | RSV | Log | 0.78 | -2.69 | 0.10 | 0.07 | 0.00 | 0.03 | -0.40 |
| UOM | RSV | Log | 0.80 | -2.17 | 0.11 | 0.08 | 0.00 | 0.03 | -0.21 |
| EQ | RSV | Log | 0.89 | -1.18 | 0.13 | 0.11 | 0.00 | 0.02 | 0.21 |
| JCU | RSV | Log | 1.00 | 8.84 | 0.33 | 0.30 | 0.00 | 0.03 | 10.46 |
| Baseline | RSV | Log | 0.94 | 2.00 | 0.44 | 0.38 | 0.00 | 0.06 | 1.95 |
| DSTG-tft | RSV | Nat | -0.27 | 8.75 | 29.48 | 1.40 | 17.21 | 10.88 | 5.34 |
| DTSG-RV | RSV | Nat | 0.78 | 10.91 | 83.23 | 59.28 | 0.00 | 23.95 | 6.32 |
| EM | RSV | Nat | 0.58 | 11.29 | 87.03 | 38.09 | 0.00 | 48.94 | 6.11 |
| UOM | RSV | Nat | 0.80 | 11.47 | 106.77 | 73.68 | 0.00 | 33.09 | 6.50 |
| EQ | RSV | Nat | 0.89 | 12.15 | 111.83 | 88.51 | 0.00 | 23.32 | 6.93 |
| JCU | RSV | Nat | 1.00 | 19.04 | 320.48 | 289.73 | 0.00 | 30.75 | 14.31 |
| Baseline | RSV | Nat | 0.94 | 14.54 | 477.23 | 383.01 | 0.00 | 94.22 | 8.57 |

Table 8: Forecast metrics for 2024-06-27 . Where EM is ensemble mixture EQ is ensemble quantile JCU is JCU-renewal UOM is UOM-seirode. Nat is for the natural scale. Op and Up are over and under prediction respectivly. Disp is dispersion and LS is the log score.

| Model | Path | Scale | Bias | Dss | Crps | Op | Up | Disp | LS |
|---|---|---|---|---|---|---|---|---|---|
| EQ | COV | Log | -0.22 | -4.11 | 0.04 | 0.00 | 0.01 | 0.03 | -1.11 |
| EM | COV | Log | -0.35 | -2.89 | 0.06 | 0.00 | 0.02 | 0.04 | -0.51 |
| DSTG-tft | COV | Log | -0.85 | -0.78 | 0.08 | 0.00 | 0.07 | 0.01 | 0.17 |
| UOM | COV | Log | -0.78 | -2.76 | 0.08 | 0.00 | 0.06 | 0.03 | -0.55 |
| DTSG-RV | COV | Log | -0.28 | -2.47 | 0.09 | 0.00 | 0.02 | 0.07 | -0.30 |
| JCU | COV | Log | 0.49 | -2.42 | 0.11 | 0.05 | 0.00 | 0.06 | -0.25 |
| Baseline | COV | Log | 0.94 | 0.98 | 0.29 | 0.25 | 0.00 | 0.04 | 1.38 |
| EQ | COV | Nat | -0.22 | 9.55 | 33.79 | 0.00 | 8.78 | 25.01 | 5.73 |
| EM | COV | Nat | -0.35 | 10.96 | 55.11 | 0.00 | 17.90 | 37.22 | 6.33 |
| DTSG-RV | COV | Nat | -0.28 | 11.04 | 70.71 | 0.00 | 16.49 | 54.23 | 6.53 |
| UOM | COV | Nat | -0.78 | 10.84 | 72.00 | 0.00 | 49.30 | 22.69 | 6.31 |
| DSTG-tft | COV | Nat | -0.85 | 13.43 | 73.33 | 0.00 | 63.16 | 10.16 | 7.03 |
| JCU | COV | Nat | 0.49 | 11.78 | 113.29 | 44.73 | 0.00 | 68.56 | 6.61 |
| Baseline | COV | Nat | 0.94 | 14.06 | 318.08 | 260.47 | 0.00 | 57.62 | 8.20 |
| Baseline | FLU | Log | 0.44 | -2.46 | 0.11 | 0.06 | 0.00 | 0.05 | -0.33 |
| EQ | FLU | Log | 0.57 | -2.12 | 0.16 | 0.11 | 0.00 | 0.05 | -0.01 |
| EM | FLU | Log | 0.40 | -1.08 | 0.23 | 0.09 | 0.00 | 0.14 | 1.08 |
| DTSG-RV | FLU | Log | 0.67 | -1.18 | 0.23 | 0.16 | 0.00 | 0.08 | 0.32 |
| UOM | FLU | Log | 0.96 | 3.35 | 0.25 | 0.22 | 0.00 | 0.02 | 6.71 |
| JCU | FLU | Log | 0.96 | 5.52 | 0.47 | 0.43 | 0.00 | 0.04 | 5.81 |
| DSTG-tft | FLU | Log | -1.00 | 333.33 | 1.00 | 0.00 | 0.99 | 0.01 | 20.00 |
| Baseline | FLU | Nat | 0.44 | 14.37 | 465.02 | 237.34 | 1.27 | 226.40 | 7.92 |
| EQ | FLU | Nat | 0.57 | 14.70 | 680.89 | 447.53 | 0.00 | 233.37 | 8.24 |
| EM | FLU | Nat | 0.40 | 15.53 | 909.61 | 368.20 | 0.00 | 541.41 | 9.46 |
| UOM | FLU | Nat | 0.96 | 17.95 | 1058.71 | 939.52 | 0.00 | 119.19 | 12.84 |
| DTSG-RV | FLU | Nat | 0.67 | 15.90 | 1104.32 | 659.55 | 0.00 | 444.77 | 8.58 |
| DSTG-tft | FLU | Nat | -1.00 | 1878.00 | 2137.37 | 0.00 | 2118.47 | 18.90 | 20.00 |
| JCU | FLU | Nat | 0.96 | 19.07 | 2435.60 | 2118.52 | 0.00 | 317.08 | 11.92 |
| DSTG-tft | RSV | Log | 0.68 | -4.22 | 0.04 | 0.03 | 0.00 | 0.01 | -1.15 |
| DTSG-RV | RSV | Log | 0.51 | -3.57 | 0.05 | 0.03 | 0.00 | 0.03 | -0.88 |
| EM | RSV | Log | 0.64 | -3.29 | 0.07 | 0.04 | 0.00 | 0.03 | -0.80 |
| UOM | RSV | Log | 0.52 | -3.24 | 0.07 | 0.04 | 0.00 | 0.03 | -0.69 |
| EQ | RSV | Log | 0.81 | -2.94 | 0.07 | 0.05 | 0.00 | 0.02 | -0.57 |
| JCU | RSV | Log | 0.87 | -0.72 | 0.13 | 0.11 | 0.00 | 0.03 | 0.58 |
| Baseline | RSV | Log | 0.98 | 3.04 | 0.50 | 0.44 | 0.00 | 0.06 | 2.60 |
| DSTG-tft | RSV | Nat | 0.68 | 8.97 | 30.63 | 20.11 | 0.00 | 10.52 | 5.41 |
| DTSG-RV | RSV | Nat | 0.51 | 9.73 | 41.21 | 20.45 | 0.00 | 20.76 | 5.68 |
| EM | RSV | Nat | 0.64 | 10.06 | 50.79 | 28.55 | 0.00 | 22.24 | 5.76 |
| EQ | RSV | Nat | 0.81 | 10.27 | 57.62 | 42.51 | 0.00 | 15.11 | 5.98 |
| UOM | RSV | Nat | 0.52 | 10.03 | 58.17 | 32.64 | 0.00 | 25.52 | 5.87 |
| JCU | RSV | Nat | 0.87 | 12.28 | 106.49 | 84.02 | 0.00 | 22.47 | 7.14 |
| Baseline | RSV | Nat | 0.98 | 14.86 | 476.31 | 391.39 | 0.00 | 84.93 | 9.00 |

Table 9: Forecast metrics for 2024-07-04 . Where EM is ensemble mixture EQ is ensemble quantile JCU is JCU-renewal UOM is UOM-seirode. Nat is for the natural scale. Op and Up are over and under prediction respectivly. Disp is dispersion and LS is the log score.

| Model | Path | Scale | Bias | Dss | Crps | Op | Up | Disp | LS |
|---|---|---|---|---|---|---|---|---|---|
| DSTG-tft | COV | Log | -0.61 | -4.48 | 0.04 | 0.00 | 0.02 | 0.01 | -1.30 |
| EQ | COV | Log | -0.34 | -4.08 | 0.04 | 0.00 | 0.01 | 0.03 | -1.10 |
| EM | COV | Log | -0.40 | -2.90 | 0.06 | 0.00 | 0.02 | 0.04 | -0.74 |
| JCU | COV | Log | 0.31 | -2.62 | 0.08 | 0.02 | 0.00 | 0.06 | -0.41 |
| DTSG-RV | COV | Log | -0.37 | -2.36 | 0.10 | 0.00 | 0.03 | 0.07 | -0.18 |
| UOM | COV | Log | -0.90 | -2.11 | 0.10 | 0.00 | 0.08 | 0.03 | 0.07 |
| Baseline | COV | Log | 0.88 | 0.05 | 0.25 | 0.21 | 0.00 | 0.04 | 0.92 |
| DSTG-tft | COV | Nat | -0.61 | 9.00 | 32.57 | 0.00 | 22.50 | 10.06 | 5.46 |
| EQ | COV | Nat | -0.34 | 9.34 | 33.46 | 0.00 | 12.04 | 21.41 | 5.66 |
| EM | COV | Nat | -0.40 | 10.65 | 46.88 | 0.00 | 14.98 | 31.90 | 6.01 |
| JCU | COV | Nat | 0.31 | 11.29 | 74.18 | 15.36 | 0.00 | 58.82 | 6.36 |
| DTSG-RV | COV | Nat | -0.37 | 10.91 | 75.17 | 0.00 | 26.01 | 49.16 | 6.58 |
| UOM | COV | Nat | -0.90 | 11.49 | 83.73 | 0.00 | 65.00 | 18.74 | 6.85 |
| Baseline | COV | Nat | 0.88 | 13.30 | 249.44 | 198.05 | 0.00 | 51.39 | 7.68 |
| DSTG-tft | FLU | Log | 0.89 | 2.97 | 0.11 | 0.10 | 0.00 | 0.01 | 4.43 |
| Baseline | FLU | Log | 0.76 | -1.71 | 0.16 | 0.11 | 0.00 | 0.05 | 0.01 |
| EM | FLU | Log | 0.85 | -1.35 | 0.17 | 0.13 | 0.00 | 0.04 | 0.73 |
| DTSG-RV | FLU | Log | 0.60 | -1.60 | 0.17 | 0.09 | 0.00 | 0.08 | 0.15 |
| EQ | FLU | Log | 0.98 | 2.72 | 0.21 | 0.18 | 0.00 | 0.02 | 2.35 |
| UOM | FLU | Log | 0.96 | 4.78 | 0.22 | 0.20 | 0.00 | 0.02 | 4.08 |
| JCU | FLU | Log | 0.97 | 2.27 | 0.26 | 0.22 | 0.00 | 0.03 | 2.15 |
| DSTG-tft | FLU | Nat | 0.89 | 18.37 | 443.70 | 392.94 | 0.00 | 50.77 | 11.67 |
| Baseline | FLU | Nat | 0.76 | 15.06 | 672.70 | 440.13 | 0.00 | 232.58 | 8.23 |
| EM | FLU | Nat | 0.85 | 15.35 | 694.50 | 506.88 | 0.00 | 187.62 | 8.91 |
| DTSG-RV | FLU | Nat | 0.60 | 15.47 | 746.10 | 368.84 | 0.00 | 377.26 | 8.36 |
| EQ | FLU | Nat | 0.98 | 18.01 | 850.61 | 737.42 | 0.00 | 113.20 | 10.42 |
| UOM | FLU | Nat | 0.96 | 19.36 | 901.66 | 808.92 | 0.00 | 92.74 | 11.56 |
| JCU | FLU | Nat | 0.97 | 17.81 | 1092.94 | 913.37 | 0.00 | 179.57 | 10.28 |
| EQ | RSV | Log | 0.36 | -4.40 | 0.04 | 0.01 | 0.00 | 0.02 | -1.24 |
| UOM | RSV | Log | 0.07 | -4.00 | 0.04 | 0.01 | 0.00 | 0.03 | -1.01 |
| EM | RSV | Log | 0.27 | -3.67 | 0.05 | 0.01 | 0.00 | 0.04 | -0.83 |
| DTSG-RV | RSV | Log | -0.53 | -3.65 | 0.06 | 0.00 | 0.03 | 0.03 | -0.94 |
| DSTG-tft | RSV | Log | 0.83 | -2.26 | 0.08 | 0.06 | 0.00 | 0.01 | -0.18 |
| JCU | RSV | Log | 0.69 | -0.99 | 0.10 | 0.07 | 0.00 | 0.03 | 0.53 |
| Baseline | RSV | Log | 0.93 | 1.44 | 0.41 | 0.35 | 0.00 | 0.06 | 1.68 |
| EQ | RSV | Nat | 0.36 | 8.50 | 23.45 | 10.21 | 0.04 | 13.20 | 5.16 |
| UOM | RSV | Nat | 0.07 | 8.80 | 26.24 | 4.87 | 1.35 | 20.02 | 5.39 |
| EM | RSV | Nat | 0.27 | 9.22 | 28.85 | 6.57 | 0.00 | 22.28 | 5.58 |
| DTSG-RV | RSV | Nat | -0.53 | 8.95 | 31.45 | 0.00 | 16.48 | 14.97 | 5.46 |
| DSTG-tft | RSV | Nat | 0.83 | 10.39 | 44.45 | 34.44 | 0.00 | 10.01 | 6.20 |
| JCU | RSV | Nat | 0.69 | 11.59 | 73.52 | 55.47 | 0.00 | 18.04 | 6.92 |
| Baseline | RSV | Nat | 0.93 | 13.65 | 318.77 | 252.56 | 0.00 | 66.22 | 8.00 |

Table 10: Forecast metrics for 2024-07-11 . Where EM is ensemble mixture EQ is ensemble quantile JCU is JCU-renewal UOM is UOM-seirode. Nat is for the natural scale. Op and Up are over and under prediction respectivly. Disp is dispersion and LS is the log score.

| Model | Path | Scale | Bias | Dss | Crps | Op | Up | Disp | LS |
|---|---|---|---|---|---|---|---|---|---|
| EQ | COV | Log | 0.32 | -3.40 | 0.06 | 0.02 | 0.00 | 0.03 | -0.76 |
| JCU | COV | Log | -0.02 | -2.66 | 0.08 | 0.00 | 0.02 | 0.06 | -0.42 |
| EM | COV | Log | -0.03 | -2.27 | 0.10 | 0.00 | 0.01 | 0.09 | 0.01 |
| DTSG-RV | COV | Log | -0.25 | -2.33 | 0.11 | 0.00 | 0.04 | 0.06 | -0.24 |
| UOM | COV | Log | -0.84 | -0.57 | 0.14 | 0.00 | 0.11 | 0.02 | 1.85 |
| Baseline | COV | Log | 0.95 | 0.76 | 0.25 | 0.21 | 0.00 | 0.04 | 1.33 |
| DSTG-tft | COV | Log | 1.00 | 99.97 | 0.39 | 0.38 | 0.00 | 0.01 | 20.00 |
| EQ | COV | Nat | 0.32 | 10.13 | 51.92 | 20.43 | 3.52 | 27.97 | 5.96 |
| JCU | COV | Nat | -0.02 | 10.76 | 63.04 | 3.12 | 12.15 | 47.76 | 6.29 |
| DTSG-RV | COV | Nat | -0.25 | 10.90 | 79.95 | 0.64 | 34.19 | 45.12 | 6.47 |
| EM | COV | Nat | -0.03 | 11.31 | 84.18 | 1.54 | 9.59 | 73.05 | 6.71 |
| UOM | COV | Nat | -0.84 | 14.00 | 103.53 | 0.00 | 87.31 | 16.21 | 9.63 |
| Baseline | COV | Nat | 0.95 | 13.85 | 249.49 | 200.54 | 0.00 | 48.96 | 8.03 |
| DSTG-tft | COV | Nat | 1.00 | 76.96 | 396.91 | 385.27 | 0.00 | 11.64 | 20.00 |
| EQ | FLU | Log | 0.42 | -2.12 | 0.09 | 0.06 | 0.01 | 0.02 | -0.17 |
| JCU | FLU | Log | 0.60 | -1.90 | 0.09 | 0.06 | 0.00 | 0.03 | 0.04 |
| EM | FLU | Log | 0.35 | -2.52 | 0.09 | 0.06 | 0.01 | 0.03 | -0.05 |
| DTSG-RV | FLU | Log | -0.13 | -2.21 | 0.10 | 0.00 | 0.02 | 0.08 | -0.17 |
| DSTG-tft | FLU | Log | 0.50 | 4.39 | 0.12 | 0.10 | 0.01 | 0.01 | 7.32 |
| Baseline | FLU | Log | 0.72 | -1.88 | 0.14 | 0.09 | 0.00 | 0.05 | -0.09 |
| UOM | FLU | Log | 0.42 | 1.03 | 0.14 | 0.11 | 0.02 | 0.02 | 0.93 |
| DTSG-RV | FLU | Nat | -0.13 | 14.13 | 331.17 | 5.11 | 76.77 | 249.29 | 8.02 |
| EQ | FLU | Nat | 0.42 | 14.15 | 346.17 | 228.92 | 32.67 | 84.59 | 8.02 |
| EM | FLU | Nat | 0.35 | 13.97 | 359.61 | 210.35 | 26.17 | 123.08 | 8.14 |
| JCU | FLU | Nat | 0.60 | 14.49 | 362.98 | 231.04 | 0.00 | 131.94 | 8.23 |
| DSTG-tft | FLU | Nat | 0.50 | 19.30 | 461.70 | 378.70 | 32.34 | 50.66 | 12.89 |
| UOM | FLU | Nat | 0.42 | 16.64 | 561.37 | 420.08 | 59.43 | 81.86 | 9.16 |
| Baseline | FLU | Nat | 0.72 | 14.92 | 562.72 | 336.90 | 0.00 | 225.82 | 8.13 |
| DSTG-tft | RSV | Log | 0.05 | -4.37 | 0.04 | 0.01 | 0.02 | 0.02 | -1.30 |
| DTSG-RV | RSV | Log | -0.17 | -3.73 | 0.05 | 0.00 | 0.01 | 0.03 | -0.92 |
| EQ | RSV | Log | -0.23 | -3.92 | 0.05 | 0.00 | 0.03 | 0.02 | -1.03 |
| EM | RSV | Log | -0.18 | -3.80 | 0.05 | 0.00 | 0.02 | 0.03 | -0.95 |
| JCU | RSV | Log | -0.22 | -3.66 | 0.06 | 0.00 | 0.03 | 0.03 | -0.85 |
| UOM | RSV | Log | -0.37 | -2.99 | 0.07 | 0.00 | 0.04 | 0.03 | -0.32 |
| Baseline | RSV | Log | 0.93 | 1.02 | 0.38 | 0.32 | 0.00 | 0.06 | 1.46 |
| DSTG-tft | RSV | Nat | 0.05 | 8.24 | 20.32 | 4.59 | 7.25 | 8.48 | 4.98 |
| DTSG-RV | RSV | Nat | -0.17 | 8.74 | 22.75 | 0.11 | 5.34 | 17.30 | 5.36 |
| EQ | RSV | Nat | -0.23 | 8.67 | 23.81 | 1.21 | 12.58 | 10.02 | 5.25 |
| EM | RSV | Nat | -0.18 | 8.67 | 24.53 | 1.19 | 9.49 | 13.85 | 5.33 |
| JCU | RSV | Nat | -0.22 | 8.80 | 28.57 | 2.07 | 12.53 | 13.98 | 5.42 |
| UOM | RSV | Nat | -0.37 | 9.65 | 34.72 | 0.56 | 19.04 | 15.13 | 6.01 |
| Baseline | RSV | Nat | 0.93 | 13.17 | 255.01 | 199.80 | 0.00 | 55.21 | 7.70 |

Table 11: Forecast metrics for 2024-07-18 . Where EM is ensemble mixture EQ is ensemble quantile JCU is JCU-renewal UOM is UOM-seirode. Nat is for the natural scale. Op and Up are over and under prediction respectivly. Disp is dispersion and LS is the log score.

| Model | Path | Scale | Bias | Dss | Crps | Op | Up | Disp | LS |
|---|---|---|---|---|---|---|---|---|---|
| DSTG-tft | COV | Log | -0.37 | -2.31 | 0.07 | 0.01 | 0.05 | 0.01 | -0.33 |
| Baseline | COV | Log | 0.52 | -2.93 | 0.08 | 0.04 | 0.00 | 0.04 | -0.60 |
| EM | COV | Log | -0.64 | -2.22 | 0.13 | 0.00 | 0.08 | 0.05 | -0.07 |
| JCU | COV | Log | -0.50 | -2.13 | 0.14 | 0.00 | 0.08 | 0.06 | -0.12 |
| EQ | COV | Log | -0.79 | -0.31 | 0.15 | 0.00 | 0.13 | 0.02 | 0.80 |
| UOM | COV | Log | -0.95 | 1.26 | 0.19 | 0.00 | 0.17 | 0.02 | 4.43 |
| DTSG-RV | COV | Log | -0.74 | -1.11 | 0.22 | 0.00 | 0.17 | 0.05 | 0.38 |
| DSTG-tft | COV | Nat | -0.37 | 11.35 | 54.15 | 4.35 | 40.03 | 9.77 | 6.39 |
| Baseline | COV | Nat | 0.52 | 10.82 | 69.15 | 30.60 | 0.00 | 38.55 | 6.12 |
| EM | COV | Nat | -0.64 | 10.89 | 95.04 | 0.00 | 64.88 | 30.15 | 6.67 |
| JCU | COV | Nat | -0.50 | 10.86 | 98.85 | 0.00 | 59.12 | 39.73 | 6.58 |
| EQ | COV | Nat | -0.79 | 14.03 | 112.40 | 0.00 | 96.94 | 15.46 | 7.51 |
| UOM | COV | Nat | -0.95 | 16.45 | 138.67 | 0.00 | 122.87 | 15.80 | 11.09 |
| DTSG-RV | COV | Nat | -0.74 | 12.27 | 148.33 | 0.00 | 118.66 | 29.67 | 7.10 |
| JCU | FLU | Log | 0.16 | -3.46 | 0.06 | 0.02 | 0.01 | 0.03 | -0.78 |
| EQ | FLU | Log | -0.04 | -3.29 | 0.07 | 0.02 | 0.02 | 0.02 | -0.72 |
| EM | FLU | Log | 0.02 | -2.75 | 0.07 | 0.01 | 0.01 | 0.04 | -0.62 |
| Baseline | FLU | Log | 0.36 | -2.80 | 0.09 | 0.04 | 0.00 | 0.05 | -0.49 |
| UOM | FLU | Log | -0.09 | -2.15 | 0.10 | 0.04 | 0.03 | 0.03 | -0.21 |
| DSTG-tft | FLU | Log | 0.53 | 3.42 | 0.10 | 0.08 | 0.00 | 0.01 | 4.38 |
| DTSG-RV | FLU | Log | -0.51 | -1.90 | 0.14 | 0.00 | 0.07 | 0.07 | -0.04 |
| JCU | FLU | Nat | 0.16 | 12.89 | 207.02 | 62.64 | 28.56 | 115.82 | 7.37 |
| EQ | FLU | Nat | -0.04 | 13.02 | 232.30 | 66.44 | 84.62 | 81.25 | 7.43 |
| EM | FLU | Nat | 0.02 | 13.39 | 238.73 | 51.56 | 44.85 | 142.33 | 7.53 |
| Baseline | FLU | Nat | 0.36 | 13.77 | 321.19 | 136.65 | 0.00 | 184.54 | 7.66 |
| UOM | FLU | Nat | -0.09 | 14.07 | 341.23 | 135.82 | 116.60 | 88.81 | 7.94 |
| DSTG-tft | FLU | Nat | 0.53 | 18.23 | 371.66 | 309.19 | 13.99 | 48.48 | 10.50 |
| DTSG-RV | FLU | Nat | -0.51 | 14.01 | 434.06 | 0.00 | 245.77 | 188.29 | 8.10 |
| DTSG-RV | RSV | Log | 0.19 | -3.74 | 0.05 | 0.01 | 0.00 | 0.04 | -0.89 |
| EQ | RSV | Log | -0.70 | -2.70 | 0.08 | 0.00 | 0.06 | 0.02 | -0.57 |
| EM | RSV | Log | -0.55 | -3.07 | 0.09 | 0.00 | 0.05 | 0.03 | -0.34 |
| UOM | RSV | Log | -0.64 | -2.39 | 0.09 | 0.00 | 0.07 | 0.03 | -0.17 |
| JCU | RSV | Log | -0.75 | -2.15 | 0.13 | 0.00 | 0.10 | 0.03 | -0.16 |
| DSTG-tft | RSV | Log | -0.98 | 3.04 | 0.17 | 0.00 | 0.15 | 0.02 | 5.48 |
| Baseline | RSV | Log | 0.90 | 0.83 | 0.35 | 0.29 | 0.00 | 0.06 | 1.31 |
| DTSG-RV | RSV | Nat | 0.19 | 8.61 | 20.92 | 4.44 | 0.33 | 16.14 | 5.24 |
| EQ | RSV | Nat | -0.70 | 9.62 | 33.62 | 0.00 | 25.20 | 8.41 | 5.55 |
| EM | RSV | Nat | -0.55 | 8.97 | 35.37 | 0.00 | 21.76 | 13.61 | 5.79 |
| UOM | RSV | Nat | -0.64 | 9.98 | 37.56 | 0.00 | 26.29 | 11.27 | 5.98 |
| JCU | RSV | Nat | -0.75 | 10.08 | 48.86 | 0.00 | 37.84 | 11.02 | 5.96 |
| DSTG-tft | RSV | Nat | -0.98 | 17.19 | 67.44 | 0.00 | 60.74 | 6.70 | 10.16 |
| Baseline | RSV | Nat | 0.90 | 12.65 | 195.45 | 153.87 | 0.00 | 41.58 | 7.41 |

Table 12: Forecast metrics for 2024-07-25 . Where EM is ensemble mixture EQ is ensemble quantile JCU is JCU-renewal UOM is UOM-seirode. Nat is for the natural scale. Op and Up are over and under prediction respectivly. Disp is dispersion and LS is the log score.

| Model | Path | Scale | Bias | Dss | Crps | Op | Up | Disp | LS |
|---|---|---|---|---|---|---|---|---|---|
| DSTG-tft | COV | Log | -0.12 | -2.44 | 0.07 | 0.02 | 0.04 | 0.01 | -0.33 |
| Baseline | COV | Log | 0.46 | -2.56 | 0.11 | 0.07 | 0.00 | 0.04 | -0.38 |
| EM | COV | Log | -0.61 | -1.82 | 0.13 | 0.00 | 0.08 | 0.05 | -0.07 |
| EQ | COV | Log | -0.83 | -0.17 | 0.15 | 0.00 | 0.12 | 0.03 | 0.96 |
| JCU | COV | Log | -0.71 | -1.39 | 0.18 | 0.00 | 0.12 | 0.06 | 0.18 |
| UOM | COV | Log | -0.90 | 1.71 | 0.18 | 0.00 | 0.16 | 0.02 | 5.23 |
| DTSG-RV | COV | Log | -0.73 | -1.15 | 0.21 | 0.00 | 0.14 | 0.07 | 0.33 |
| DSTG-tft | COV | Nat | -0.12 | 11.01 | 52.73 | 10.45 | 32.36 | 9.92 | 6.33 |
| Baseline | COV | Nat | 0.46 | 10.86 | 86.12 | 50.27 | 0.01 | 35.85 | 6.27 |
| EM | COV | Nat | -0.61 | 11.19 | 91.65 | 0.00 | 59.21 | 32.44 | 6.59 |
| EQ | COV | Nat | -0.83 | 13.88 | 106.99 | 0.00 | 91.13 | 15.87 | 7.62 |
| JCU | COV | Nat | -0.71 | 11.54 | 119.97 | 0.00 | 84.44 | 35.53 | 6.79 |
| UOM | COV | Nat | -0.90 | 16.94 | 130.71 | 0.00 | 115.73 | 14.98 | 12.45 |
| DTSG-RV | COV | Nat | -0.73 | 11.73 | 133.84 | 0.00 | 97.39 | 36.45 | 6.97 |
| EQ | FLU | Log | -0.36 | -2.72 | 0.08 | 0.01 | 0.05 | 0.03 | -0.43 |
| UOM | FLU | Log | -0.44 | -2.70 | 0.08 | 0.00 | 0.04 | 0.04 | -0.30 |
| EM | FLU | Log | -0.26 | -2.36 | 0.09 | 0.00 | 0.03 | 0.07 | -0.28 |
| DTSG-RV | FLU | Log | -0.46 | -1.63 | 0.14 | 0.00 | 0.07 | 0.07 | 0.15 |
| DSTG-tft | FLU | Log | 0.68 | 9.47 | 0.15 | 0.14 | 0.00 | 0.01 | 5.55 |
| JCU | FLU | Log | -0.84 | 0.69 | 0.19 | 0.00 | 0.15 | 0.04 | 1.79 |
| Baseline | FLU | Log | 0.42 | -1.30 | 0.19 | 0.14 | 0.00 | 0.05 | 0.35 |
| UOM | FLU | Nat | -0.44 | 13.28 | 256.41 | 0.38 | 152.89 | 103.14 | 7.75 |
| EQ | FLU | Nat | -0.36 | 13.47 | 257.03 | 12.56 | 163.79 | 80.68 | 7.60 |
| EM | FLU | Nat | -0.26 | 13.55 | 279.04 | 2.61 | 89.38 | 187.04 | 7.74 |
| DTSG-RV | FLU | Nat | -0.46 | 14.26 | 423.61 | 1.20 | 235.69 | 186.72 | 8.18 |
| DSTG-tft | FLU | Nat | 0.68 | 22.02 | 458.17 | 411.14 | 2.39 | 44.64 | 11.68 |
| JCU | FLU | Nat | -0.84 | 17.68 | 562.23 | 0.00 | 474.90 | 87.33 | 10.26 |
| Baseline | FLU | Nat | 0.42 | 14.56 | 606.90 | 413.82 | 1.60 | 191.49 | 8.34 |
| EQ | RSV | Log | -0.42 | -3.77 | 0.05 | 0.00 | 0.03 | 0.02 | -0.95 |
| EM | RSV | Log | -0.37 | -3.11 | 0.08 | 0.00 | 0.03 | 0.04 | -0.25 |
| JCU | RSV | Log | -0.49 | -3.07 | 0.08 | 0.00 | 0.05 | 0.03 | -0.60 |
| UOM | RSV | Log | -0.74 | -2.05 | 0.10 | 0.00 | 0.08 | 0.03 | 0.05 |
| DTSG-RV | RSV | Log | 0.71 | -2.37 | 0.11 | 0.07 | 0.00 | 0.03 | -0.24 |
| DSTG-tft | RSV | Log | -0.96 | 2.42 | 0.17 | 0.00 | 0.15 | 0.02 | 2.74 |
| Baseline | RSV | Log | 0.94 | 1.27 | 0.39 | 0.33 | 0.00 | 0.06 | 1.60 |
| EQ | RSV | Nat | -0.42 | 8.08 | 20.12 | 0.12 | 11.47 | 8.53 | 5.02 |
| EM | RSV | Nat | -0.37 | 8.76 | 27.88 | 0.01 | 12.46 | 15.41 | 5.72 |
| JCU | RSV | Nat | -0.49 | 8.69 | 28.79 | 0.09 | 17.54 | 11.17 | 5.36 |
| UOM | RSV | Nat | -0.74 | 10.03 | 37.34 | 0.00 | 28.35 | 8.98 | 6.05 |
| DTSG-RV | RSV | Nat | 0.71 | 9.77 | 44.39 | 30.30 | 0.00 | 14.08 | 5.73 |
| DSTG-tft | RSV | Nat | -0.96 | 15.97 | 60.01 | 0.00 | 54.02 | 5.99 | 9.43 |
| Baseline | RSV | Nat | 0.94 | 12.68 | 194.83 | 154.05 | 0.00 | 40.78 | 7.52 |

Table 13: Forecast metrics for 2024-08-01 . Where EM is ensemble mixture EQ is ensemble quantile JCU is JCU-renewal UOM is UOM-seirode. Nat is for the natural scale. Op and Up are over and under prediction respectivly. Disp is dispersion and LS is the log score.

| Model | Path | Scale | Bias | Dss | Crps | Op | Up | Disp | LS |
|---|---|---|---|---|---|---|---|---|---|
| DSTG-tft | COV | Log | 0.13 | -0.95 | 0.09 | 0.06 | 0.02 | 0.01 | 1.47 |
| JCU | COV | Log | -0.33 | -2.14 | 0.10 | 0.00 | 0.03 | 0.06 | -0.16 |
| EQ | COV | Log | -0.27 | -0.79 | 0.10 | 0.03 | 0.05 | 0.02 | 0.58 |
| EM | COV | Log | -0.23 | -1.33 | 0.11 | 0.03 | 0.05 | 0.03 | 0.69 |
| Baseline | COV | Log | 0.43 | -2.02 | 0.14 | 0.10 | 0.00 | 0.04 | -0.08 |
| DTSG-RV | COV | Log | -0.59 | -1.60 | 0.15 | 0.00 | 0.08 | 0.07 | 0.08 |
| UOM | COV | Log | -0.83 | 1.57 | 0.16 | 0.00 | 0.13 | 0.02 | 2.90 |
| DSTG-tft | COV | Nat | 0.13 | 11.71 | 62.23 | 40.10 | 12.16 | 9.98 | 7.83 |
| JCU | COV | Nat | -0.33 | 11.05 | 69.44 | 0.43 | 27.43 | 41.58 | 6.41 |
| EQ | COV | Nat | -0.27 | 12.27 | 74.10 | 20.32 | 40.15 | 13.63 | 7.17 |
| EM | COV | Nat | -0.23 | 11.50 | 74.74 | 20.05 | 34.14 | 20.54 | 7.16 |
| DTSG-RV | COV | Nat | -0.59 | 11.26 | 95.81 | 0.00 | 58.79 | 37.01 | 6.65 |
| Baseline | COV | Nat | 0.43 | 11.19 | 108.84 | 70.27 | 2.80 | 35.78 | 6.49 |
| UOM | COV | Nat | -0.83 | 16.63 | 112.00 | 0.00 | 98.27 | 13.73 | 10.43 |
| UOM | FLU | Log | -0.37 | -2.53 | 0.09 | 0.01 | 0.05 | 0.04 | -0.06 |
| JCU | FLU | Log | 0.38 | -0.49 | 0.20 | 0.15 | 0.02 | 0.03 | 0.65 |
| EM | FLU | Log | 0.60 | 20.17 | 0.23 | 0.18 | 0.01 | 0.04 | 5.31 |
| EQ | FLU | Log | 0.66 | 24.38 | 0.25 | 0.22 | 0.01 | 0.02 | 5.78 |
| DTSG-RV | FLU | Log | 0.46 | -0.90 | 0.28 | 0.20 | 0.00 | 0.08 | 0.47 |
| DSTG-tft | FLU | Log | 0.97 | 42.00 | 0.30 | 0.28 | 0.00 | 0.01 | 10.64 |
| Baseline | FLU | Log | 0.68 | 3.14 | 0.41 | 0.36 | 0.00 | 0.05 | 5.67 |
| UOM | FLU | Nat | -0.37 | 13.17 | 247.32 | 7.89 | 157.66 | 81.76 | 7.75 |
| JCU | FLU | Nat | 0.38 | 14.78 | 488.59 | 317.88 | 70.39 | 100.32 | 8.42 |
| EM | FLU | Nat | 0.60 | 27.31 | 604.10 | 443.94 | 25.81 | 134.35 | 11.26 |
| EQ | FLU | Nat | 0.66 | 29.52 | 630.80 | 521.46 | 38.41 | 70.93 | 11.61 |
| DTSG-RV | FLU | Nat | 0.46 | 15.11 | 764.12 | 457.70 | 16.36 | 290.06 | 8.30 |
| DSTG-tft | FLU | Nat | 0.97 | 40.12 | 775.61 | 731.96 | 0.00 | 43.65 | 14.71 |
| Baseline | FLU | Nat | 0.68 | 16.40 | 1143.00 | 945.77 | 1.98 | 195.25 | 10.48 |
| DTSG-RV | RSV | Log | -0.19 | -3.59 | 0.04 | 0.00 | 0.01 | 0.03 | -0.91 |
| UOM | RSV | Log | -0.94 | -1.06 | 0.13 | 0.00 | 0.11 | 0.03 | 0.51 |
| EM | RSV | Log | -0.77 | 0.40 | 0.14 | 0.00 | 0.11 | 0.03 | 1.29 |
| EQ | RSV | Log | -0.93 | 1.45 | 0.15 | 0.00 | 0.13 | 0.02 | 3.65 |
| JCU | RSV | Log | -0.89 | -1.13 | 0.15 | 0.00 | 0.12 | 0.03 | 0.36 |
| DSTG-tft | RSV | Log | -0.99 | 3.76 | 0.20 | 0.00 | 0.18 | 0.02 | 3.08 |
| Baseline | RSV | Log | 0.90 | 1.11 | 0.39 | 0.33 | 0.00 | 0.06 | 1.54 |
| DTSG-RV | RSV | Nat | -0.19 | 8.02 | 14.97 | 0.11 | 3.53 | 11.33 | 4.94 |
| UOM | RSV | Nat | -0.94 | 10.90 | 42.66 | 0.00 | 35.13 | 7.53 | 6.41 |
| EM | RSV | Nat | -0.77 | 13.06 | 44.10 | 0.00 | 35.49 | 8.60 | 7.36 |
| JCU | RSV | Nat | -0.89 | 10.60 | 47.75 | 0.00 | 39.22 | 8.53 | 6.23 |
| EQ | RSV | Nat | -0.93 | 14.54 | 47.94 | 0.00 | 41.99 | 5.95 | 9.00 |
| DSTG-tft | RSV | Nat | -0.99 | 17.61 | 62.77 | 0.00 | 57.13 | 5.63 | 9.36 |
| Baseline | RSV | Nat | 0.90 | 12.33 | 174.41 | 136.26 | 0.00 | 38.15 | 7.34 |

Table 14: Forecast metrics for 2024-08-08 . Where EM is ensemble mixture EQ is ensemble quantile JCU is JCU-renewal UOM is UOM-seirode. Nat is for the natural scale. Op and Up are over and under prediction respectivly. Disp is dispersion and LS is the log score.

| Model | Path | Scale | Bias | Dss | Crps | Op | Up | Disp | LS |
|---|---|---|---|---|---|---|---|---|---|
| EM | COV | Log | 0.25 | -2.28 | 0.10 | 0.04 | 0.00 | 0.06 | -0.30 |
| UOM | COV | Log | -0.74 | -0.99 | 0.11 | 0.00 | 0.08 | 0.02 | 1.37 |
| EQ | COV | Log | 0.55 | -1.92 | 0.13 | 0.09 | 0.00 | 0.03 | -0.05 |
| JCU | COV | Log | 0.42 | -2.04 | 0.13 | 0.07 | 0.00 | 0.07 | -0.11 |
| DSTG-tft | COV | Log | 0.67 | 8.93 | 0.16 | 0.15 | 0.00 | 0.01 | 8.24 |
| DTSG-RV | COV | Log | 0.66 | -1.41 | 0.19 | 0.13 | 0.00 | 0.06 | 0.18 |
| Baseline | COV | Log | 0.81 | -0.29 | 0.22 | 0.18 | 0.00 | 0.04 | 0.76 |
| UOM | COV | Nat | -0.74 | 12.84 | 70.92 | 0.00 | 58.30 | 12.62 | 8.66 |
| EM | COV | Nat | 0.25 | 11.22 | 73.08 | 27.04 | 1.39 | 44.65 | 6.21 |
| EQ | COV | Nat | 0.55 | 11.10 | 89.27 | 61.77 | 3.12 | 24.38 | 6.45 |
| JCU | COV | Nat | 0.42 | 11.51 | 102.76 | 45.57 | 0.18 | 57.01 | 6.41 |
| DSTG-tft | COV | Nat | 0.67 | 18.93 | 114.03 | 103.27 | 0.74 | 10.02 | 12.67 |
| DTSG-RV | COV | Nat | 0.66 | 12.10 | 152.34 | 92.54 | 0.00 | 59.80 | 6.71 |
| Baseline | COV | Nat | 0.81 | 12.45 | 163.47 | 129.07 | 0.00 | 34.39 | 7.24 |
| UOM | FLU | Log | 0.18 | -3.17 | 0.08 | 0.03 | 0.02 | 0.03 | -0.60 |
| EM | FLU | Log | 0.74 | -0.52 | 0.30 | 0.22 | 0.00 | 0.07 | 0.42 |
| JCU | FLU | Log | 0.84 | 1.58 | 0.32 | 0.28 | 0.00 | 0.04 | 1.40 |
| EQ | FLU | Log | 0.91 | 6.86 | 0.40 | 0.36 | 0.00 | 0.03 | 7.73 |
| DSTG-tft | FLU | Log | 1.00 | 129.48 | 0.56 | 0.55 | 0.00 | 0.01 | 20.00 |
| DTSG-RV | FLU | Log | 0.95 | 5.09 | 0.59 | 0.53 | 0.00 | 0.06 | 3.88 |
| Baseline | FLU | Log | 0.99 | 13.06 | 0.69 | 0.64 | 0.00 | 0.04 | 11.85 |
| UOM | FLU | Nat | 0.18 | 12.05 | 152.42 | 46.79 | 49.23 | 56.40 | 6.97 |
| JCU | FLU | Nat | 0.84 | 15.68 | 663.02 | 546.29 | 0.00 | 116.73 | 9.00 |
| EM | FLU | Nat | 0.74 | 14.97 | 663.67 | 445.72 | 0.00 | 217.95 | 8.02 |
| EQ | FLU | Nat | 0.91 | 18.34 | 850.36 | 747.20 | 0.00 | 103.16 | 12.79 |
| DSTG-tft | FLU | Nat | 1.00 | 80.01 | 1350.93 | 1308.12 | 0.00 | 42.81 | 20.00 |
| DTSG-RV | FLU | Nat | 0.95 | 17.48 | 1503.90 | 1260.11 | 0.00 | 243.79 | 10.69 |
| Baseline | FLU | Nat | 0.99 | 20.17 | 1808.77 | 1616.55 | 0.00 | 192.23 | 14.98 |
| EQ | RSV | Log | -0.65 | -3.41 | 0.07 | 0.00 | 0.04 | 0.03 | -0.84 |
| DTSG-RV | RSV | Log | 0.52 | -3.31 | 0.07 | 0.03 | 0.00 | 0.03 | -0.70 |
| EM | RSV | Log | -0.52 | -3.01 | 0.08 | 0.00 | 0.04 | 0.04 | -0.19 |
| JCU | RSV | Log | -0.71 | -2.45 | 0.11 | 0.00 | 0.08 | 0.04 | -0.31 |
| UOM | RSV | Log | -0.92 | -1.36 | 0.12 | 0.00 | 0.10 | 0.02 | 0.37 |
| DSTG-tft | RSV | Log | -0.97 | 0.57 | 0.15 | 0.00 | 0.13 | 0.02 | 1.26 |
| Baseline | RSV | Log | 0.92 | 0.90 | 0.36 | 0.30 | 0.00 | 0.06 | 1.42 |
| EQ | RSV | Nat | -0.65 | 7.88 | 18.83 | 0.00 | 11.85 | 6.99 | 4.89 |
| DTSG-RV | RSV | Nat | 0.52 | 8.42 | 22.04 | 10.22 | 0.00 | 11.82 | 5.04 |
| EM | RSV | Nat | -0.52 | 8.27 | 23.60 | 0.00 | 12.75 | 10.85 | 5.54 |
| JCU | RSV | Nat | -0.71 | 8.74 | 30.15 | 0.00 | 20.63 | 9.52 | 5.43 |
| UOM | RSV | Nat | -0.92 | 10.31 | 34.85 | 0.00 | 28.41 | 6.43 | 6.17 |
| DSTG-tft | RSV | Nat | -0.97 | 12.79 | 43.43 | 0.00 | 38.20 | 5.23 | 7.02 |
| Baseline | RSV | Nat | 0.92 | 11.96 | 138.53 | 109.16 | 0.00 | 29.38 | 7.10 |

Table 15: Forecast metrics for 2024-08-15 . Where EM is ensemble mixture EQ is ensemble quantile JCU is JCU-renewal UOM is UOM-seirode. Nat is for the natural scale. Op and Up are over and under prediction respectivly. Disp is dispersion and LS is the log score.

| Model | Path | Scale | Bias | Dss | Crps | Op | Up | Disp | LS |
|---|---|---|---|---|---|---|---|---|---|
| UOM | COV | Log | -0.67 | -2.57 | 0.08 | 0.00 | 0.06 | 0.02 | -0.14 |
| EM | COV | Log | 0.38 | -2.29 | 0.11 | 0.04 | 0.00 | 0.07 | -0.06 |
| EQ | COV | Log | 0.80 | -1.71 | 0.12 | 0.09 | 0.00 | 0.03 | 0.09 |
| JCU | COV | Log | 0.54 | -1.82 | 0.12 | 0.06 | 0.00 | 0.06 | -0.03 |
| DTSG-RV | COV | Log | 0.65 | -1.63 | 0.14 | 0.09 | 0.00 | 0.06 | 0.08 |
| Baseline | COV | Log | 0.91 | 0.62 | 0.22 | 0.18 | 0.00 | 0.04 | 1.22 |
| DSTG-tft | COV | Log | 1.00 | 34.35 | 0.30 | 0.28 | 0.00 | 0.01 | 20.00 |
| UOM | COV | Nat | -0.67 | 10.60 | 52.16 | 0.00 | 40.39 | 11.77 | 6.38 |
| EM | COV | Nat | 0.38 | 11.07 | 80.61 | 27.33 | 0.00 | 53.28 | 6.41 |
| EQ | COV | Nat | 0.80 | 11.26 | 81.17 | 60.62 | 0.00 | 20.55 | 6.55 |
| JCU | COV | Nat | 0.54 | 11.49 | 87.28 | 41.14 | 0.00 | 46.14 | 6.45 |
| DTSG-RV | COV | Nat | 0.65 | 11.69 | 106.71 | 61.40 | 0.00 | 45.31 | 6.57 |
| Baseline | COV | Nat | 0.91 | 13.25 | 168.78 | 133.77 | 0.00 | 35.01 | 7.69 |
| DSTG-tft | COV | Nat | 1.00 | 36.29 | 224.39 | 214.01 | 0.00 | 10.38 | 20.00 |
| UOM | FLU | Log | 0.79 | -1.91 | 0.12 | 0.09 | 0.00 | 0.03 | -0.13 |
| EM | FLU | Log | 0.94 | 0.49 | 0.36 | 0.27 | 0.00 | 0.09 | 0.99 |
| JCU | FLU | Log | 1.00 | 6.32 | 0.39 | 0.35 | 0.00 | 0.04 | 5.10 |
| DTSG-RV | FLU | Log | 0.95 | 1.73 | 0.46 | 0.39 | 0.00 | 0.07 | 1.71 |
| EQ | FLU | Log | 1.00 | 11.57 | 0.49 | 0.45 | 0.00 | 0.03 | 12.02 |
| Baseline | FLU | Log | 1.00 | 16.89 | 0.79 | 0.75 | 0.00 | 0.05 | 15.27 |
| DSTG-tft | FLU | Log | 1.00 | 252.34 | 0.83 | 0.82 | 0.00 | 0.01 | 20.00 |
| UOM | FLU | Nat | 0.79 | 12.69 | 172.30 | 121.20 | 0.00 | 51.10 | 7.14 |
| EM | FLU | Nat | 0.94 | 15.27 | 659.26 | 439.81 | 0.00 | 219.45 | 8.22 |
| JCU | FLU | Nat | 1.00 | 18.40 | 669.49 | 591.49 | 0.00 | 77.99 | 11.63 |
| DTSG-RV | FLU | Nat | 0.95 | 15.95 | 868.31 | 677.52 | 0.00 | 190.79 | 8.94 |
| EQ | FLU | Nat | 1.00 | 20.91 | 869.79 | 786.64 | 0.00 | 83.16 | 15.00 |
| Baseline | FLU | Nat | 1.00 | 21.28 | 1693.76 | 1531.73 | 0.00 | 162.03 | 16.22 |
| DSTG-tft | FLU | Nat | 1.00 | 121.90 | 1784.09 | 1742.26 | 0.00 | 41.84 | 20.00 |
| EQ | RSV | Log | -0.40 | -3.87 | 0.05 | 0.00 | 0.02 | 0.03 | -0.97 |
| JCU | RSV | Log | -0.24 | -3.51 | 0.05 | 0.00 | 0.01 | 0.04 | -0.87 |
| EM | RSV | Log | -0.41 | -3.29 | 0.06 | 0.00 | 0.02 | 0.04 | -0.59 |
| DTSG-RV | RSV | Log | 0.30 | -3.01 | 0.06 | 0.01 | 0.00 | 0.05 | -0.57 |
| DSTG-tft | RSV | Log | -0.83 | -2.18 | 0.10 | 0.00 | 0.07 | 0.02 | -0.21 |
| UOM | RSV | Log | -0.86 | -2.16 | 0.10 | 0.00 | 0.08 | 0.02 | -0.18 |
| Baseline | RSV | Log | 0.94 | 1.21 | 0.37 | 0.32 | 0.00 | 0.06 | 1.59 |
| EQ | RSV | Nat | -0.40 | 7.17 | 11.97 | 0.00 | 5.13 | 6.84 | 4.61 |
| JCU | RSV | Nat | -0.24 | 7.53 | 11.98 | 0.07 | 2.65 | 9.25 | 4.70 |
| EM | RSV | Nat | -0.41 | 7.86 | 15.14 | 0.00 | 6.05 | 9.10 | 4.99 |
| DTSG-RV | RSV | Nat | 0.30 | 8.45 | 17.67 | 4.06 | 0.00 | 13.60 | 5.02 |
| DSTG-tft | RSV | Nat | -0.83 | 9.05 | 23.96 | 0.00 | 18.95 | 5.01 | 5.38 |
| UOM | RSV | Nat | -0.86 | 9.02 | 25.37 | 0.00 | 19.77 | 5.60 | 5.42 |
| Baseline | RSV | Nat | 0.94 | 11.84 | 124.92 | 99.32 | 0.00 | 25.60 | 7.08 |

Table 16: Forecast metrics for 2024-08-22 . Where EM is ensemble mixture EQ is ensemble quantile JCU is JCU-renewal UOM is UOM-seirode. Nat is for the natural scale. Op and Up are over and under prediction respectivly. Disp is dispersion and LS is the log score.

| Model | Path | Scale | Bias | Dss | Crps | Op | Up | Disp | LS |
|---|---|---|---|---|---|---|---|---|---|
| Baseline | COV | Log | 0.30 | -3.49 | 0.05 | 0.02 | 0.00 | 0.03 | -0.84 |
| EQ | COV | Log | -0.35 | -3.31 | 0.07 | 0.00 | 0.04 | 0.03 | -0.78 |
| EM | COV | Log | -0.17 | -2.47 | 0.09 | 0.00 | 0.03 | 0.06 | -0.08 |
| UOM | COV | Log | -0.67 | -2.05 | 0.09 | 0.00 | 0.07 | 0.02 | 0.11 |
| JCU | COV | Log | -0.34 | -2.31 | 0.13 | 0.00 | 0.07 | 0.05 | -0.19 |
| DSTG-tft | COV | Log | 1.00 | 4.89 | 0.15 | 0.14 | 0.00 | 0.01 | 6.32 |
| DTSG-RV | COV | Log | -0.67 | -1.30 | 0.22 | 0.00 | 0.15 | 0.07 | 0.28 |
| Baseline | COV | Nat | 0.30 | 9.63 | 33.93 | 9.95 | 0.13 | 23.85 | 5.65 |
| EQ | COV | Nat | -0.35 | 9.59 | 46.37 | 2.26 | 27.46 | 16.65 | 5.69 |
| EM | COV | Nat | -0.17 | 10.16 | 54.98 | 0.84 | 16.77 | 37.38 | 6.51 |
| UOM | COV | Nat | -0.67 | 11.26 | 58.51 | 0.00 | 47.07 | 11.43 | 6.65 |
| JCU | COV | Nat | -0.34 | 10.38 | 75.74 | 2.12 | 44.12 | 29.50 | 6.30 |
| DSTG-tft | COV | Nat | 1.00 | 16.40 | 107.68 | 97.80 | 0.00 | 9.88 | 11.83 |
| DTSG-RV | COV | Nat | -0.67 | 11.29 | 119.97 | 0.00 | 89.94 | 30.03 | 6.76 |
| DTSG-RV | FLU | Log | 0.46 | -2.20 | 0.11 | 0.04 | 0.00 | 0.07 | -0.20 |
| UOM | FLU | Log | 0.90 | -1.41 | 0.14 | 0.10 | 0.00 | 0.03 | 0.04 |
| EM | FLU | Log | 0.83 | -1.72 | 0.16 | 0.12 | 0.00 | 0.03 | 0.42 |
| EQ | FLU | Log | 0.98 | 1.93 | 0.18 | 0.16 | 0.00 | 0.02 | 1.35 |
| JCU | FLU | Log | 0.96 | 1.80 | 0.19 | 0.17 | 0.00 | 0.03 | 2.04 |
| DSTG-tft | FLU | Log | 1.00 | 43.48 | 0.24 | 0.23 | 0.00 | 0.01 | 14.20 |
| Baseline | FLU | Log | 1.00 | 10.29 | 0.57 | 0.53 | 0.00 | 0.04 | 5.71 |
| DTSG-RV | FLU | Nat | 0.46 | 12.53 | 146.07 | 52.38 | 0.00 | 93.68 | 6.93 |
| UOM | FLU | Nat | 0.90 | 12.89 | 185.98 | 135.15 | 0.00 | 50.84 | 7.16 |
| EM | FLU | Nat | 0.83 | 12.87 | 221.73 | 171.66 | 0.00 | 50.07 | 7.53 |
| EQ | FLU | Nat | 0.98 | 15.45 | 249.84 | 217.92 | 0.00 | 31.92 | 8.50 |
| JCU | FLU | Nat | 0.96 | 15.31 | 275.18 | 233.66 | 0.00 | 41.52 | 9.04 |
| DSTG-tft | FLU | Nat | 1.00 | 44.81 | 356.19 | 340.93 | 0.00 | 15.26 | 16.48 |
| Baseline | FLU | Nat | 1.00 | 19.16 | 946.26 | 845.28 | 0.00 | 100.98 | 12.46 |
| DTSG-RV | RSV | Log | -0.06 | -3.40 | 0.04 | 0.00 | 0.00 | 0.04 | -0.80 |
| JCU | RSV | Log | -0.45 | -3.61 | 0.05 | 0.00 | 0.02 | 0.03 | -0.90 |
| EQ | RSV | Log | -0.71 | -3.25 | 0.07 | 0.00 | 0.05 | 0.02 | -0.73 |
| EM | RSV | Log | -0.60 | -3.12 | 0.08 | 0.00 | 0.04 | 0.03 | -0.49 |
| UOM | RSV | Log | -0.90 | -2.03 | 0.11 | 0.00 | 0.08 | 0.02 | -0.15 |
| DSTG-tft | RSV | Log | -0.97 | 0.03 | 0.15 | 0.00 | 0.13 | 0.02 | 1.12 |
| Baseline | RSV | Log | 0.80 | -0.61 | 0.24 | 0.19 | 0.00 | 0.05 | 0.69 |
| DTSG-RV | RSV | Nat | -0.06 | 7.66 | 11.06 | 0.04 | 0.56 | 10.45 | 4.72 |
| JCU | RSV | Nat | -0.45 | 7.25 | 12.00 | 0.00 | 4.89 | 7.11 | 4.62 |
| EQ | RSV | Nat | -0.71 | 7.63 | 17.09 | 0.00 | 11.64 | 5.44 | 4.79 |
| EM | RSV | Nat | -0.60 | 7.72 | 18.19 | 0.00 | 10.63 | 7.56 | 5.03 |
| UOM | RSV | Nat | -0.90 | 9.05 | 25.74 | 0.00 | 20.62 | 5.12 | 5.37 |
| DSTG-tft | RSV | Nat | -0.97 | 11.58 | 35.94 | 0.00 | 31.28 | 4.66 | 6.69 |
| Baseline | RSV | Nat | 0.80 | 10.46 | 71.42 | 53.10 | 0.00 | 18.32 | 6.18 |

Table 17: Forecast metrics for 2024-08-29 . Where EM is ensemble mixture EQ is ensemble quantile JCU is JCU-renewal UOM is UOM-seirode. Nat is for the natural scale. Op and Up are over and under prediction respectivly. Disp is dispersion and LS is the log score.

| Model | Path | Scale | Bias | Dss | Crps | Op | Up | Disp | LS |
|---|---|---|---|---|---|---|---|---|---|
| Baseline | COV | Log | -0.43 | -3.80 | 0.05 | 0.00 | 0.02 | 0.03 | -0.93 |
| DSTG-tft | COV | Log | -0.56 | -1.83 | 0.09 | 0.00 | 0.07 | 0.02 | -0.07 |
| UOM | COV | Log | -0.92 | -0.36 | 0.13 | 0.00 | 0.11 | 0.02 | 0.87 |
| EM | COV | Log | -0.82 | -1.73 | 0.15 | 0.00 | 0.11 | 0.04 | 0.31 |
| EQ | COV | Log | -0.94 | 1.47 | 0.18 | 0.00 | 0.16 | 0.02 | 2.75 |
| DTSG-RV | COV | Log | -0.88 | -0.74 | 0.22 | 0.00 | 0.17 | 0.05 | 0.46 |
| JCU | COV | Log | -0.93 | 0.14 | 0.25 | 0.00 | 0.21 | 0.04 | 1.09 |
| Baseline | COV | Nat | -0.43 | 9.08 | 32.58 | 0.00 | 14.09 | 18.49 | 5.59 |
| DSTG-tft | COV | Nat | -0.56 | 11.58 | 57.51 | 0.00 | 46.77 | 10.73 | 6.45 |
| UOM | COV | Nat | -0.92 | 13.34 | 82.91 | 0.00 | 71.91 | 11.00 | 7.40 |
| EM | COV | Nat | -0.82 | 11.33 | 92.18 | 0.00 | 70.51 | 21.66 | 6.89 |
| EQ | COV | Nat | -0.94 | 15.98 | 109.63 | 0.00 | 97.67 | 11.96 | 9.77 |
| DTSG-RV | COV | Nat | -0.88 | 12.26 | 129.73 | 0.00 | 106.02 | 23.71 | 6.98 |
| JCU | COV | Nat | -0.93 | 13.62 | 145.16 | 0.00 | 125.69 | 19.46 | 7.75 |
| EQ | FLU | Log | 0.51 | -3.96 | 0.05 | 0.03 | 0.00 | 0.02 | -1.05 |
| EM | FLU | Log | 0.31 | -3.36 | 0.05 | 0.02 | 0.00 | 0.03 | -0.90 |
| DSTG-tft | FLU | Log | 0.45 | -3.11 | 0.05 | 0.04 | 0.00 | 0.01 | -0.48 |
| DTSG-RV | FLU | Log | 0.30 | -3.16 | 0.06 | 0.01 | 0.00 | 0.05 | -0.64 |
| JCU | FLU | Log | -0.40 | -3.76 | 0.06 | 0.00 | 0.03 | 0.02 | -0.97 |
| UOM | FLU | Log | 0.88 | -1.70 | 0.13 | 0.10 | 0.00 | 0.04 | -0.10 |
| Baseline | FLU | Log | 1.00 | 4.96 | 0.38 | 0.34 | 0.00 | 0.04 | 3.43 |
| JCU | FLU | Nat | -0.40 | 10.08 | 54.62 | 0.00 | 30.90 | 23.72 | 6.00 |
| EQ | FLU | Nat | 0.51 | 10.11 | 55.94 | 31.30 | 0.00 | 24.65 | 5.92 |
| EM | FLU | Nat | 0.31 | 10.80 | 57.91 | 19.89 | 0.00 | 38.01 | 6.08 |
| DSTG-tft | FLU | Nat | 0.45 | 10.64 | 61.27 | 48.84 | 0.08 | 12.35 | 6.48 |
| DTSG-RV | FLU | Nat | 0.30 | 11.02 | 65.74 | 14.49 | 0.00 | 51.25 | 6.34 |
| UOM | FLU | Nat | 0.88 | 12.41 | 157.93 | 110.34 | 0.00 | 47.58 | 6.89 |
| Baseline | FLU | Nat | 1.00 | 16.78 | 499.85 | 437.39 | 0.00 | 62.46 | 10.22 |
| JCU | RSV | Log | 0.01 | -4.16 | 0.03 | 0.00 | 0.00 | 0.03 | -1.14 |
| EQ | RSV | Log | -0.41 | -4.18 | 0.04 | 0.00 | 0.01 | 0.02 | -1.17 |
| DTSG-RV | RSV | Log | 0.17 | -3.53 | 0.04 | 0.00 | 0.00 | 0.04 | -0.81 |
| EM | RSV | Log | -0.37 | -3.59 | 0.05 | 0.00 | 0.01 | 0.04 | -0.79 |
| UOM | RSV | Log | -0.74 | -3.36 | 0.07 | 0.00 | 0.04 | 0.02 | -0.84 |
| DSTG-tft | RSV | Log | -0.93 | -1.57 | 0.12 | 0.00 | 0.10 | 0.02 | 0.05 |
| Baseline | RSV | Log | 0.92 | -0.07 | 0.25 | 0.20 | 0.00 | 0.04 | 0.89 |
| JCU | RSV | Nat | 0.01 | 6.73 | 6.74 | 0.02 | 0.00 | 6.72 | 4.29 |
| EQ | RSV | Nat | -0.41 | 6.53 | 8.23 | 0.00 | 2.83 | 5.39 | 4.25 |
| DTSG-RV | RSV | Nat | 0.17 | 7.49 | 10.32 | 0.88 | 0.00 | 9.44 | 4.63 |
| EM | RSV | Nat | -0.37 | 7.18 | 10.57 | 0.00 | 3.01 | 7.56 | 4.64 |
| UOM | RSV | Nat | -0.74 | 7.33 | 14.41 | 0.00 | 9.55 | 4.85 | 4.59 |
| DSTG-tft | RSV | Nat | -0.93 | 9.37 | 25.08 | 0.00 | 20.53 | 4.55 | 5.50 |
| Baseline | RSV | Nat | 0.92 | 10.69 | 66.68 | 52.47 | 0.00 | 14.21 | 6.30 |

Table 18: Forecast metrics for 2024-09-05 . Where EM is ensemble mixture EQ is ensemble quantile JCU is JCU-renewal UOM is UOM-seirode. Nat is for the natural scale. Op and Up are over and under prediction respectivly. Disp is dispersion and LS is the log score.

| Model | Path | Scale | Bias | Dss | Crps | Op | Up | Disp | LS |
|---|---|---|---|---|---|---|---|---|---|
| Baseline | COV | Log | -0.53 | -3.96 | 0.05 | 0.00 | 0.02 | 0.02 | -1.01 |
| DSTG-tft | COV | Log | -0.83 | -3.28 | 0.07 | 0.00 | 0.05 | 0.02 | -0.74 |
| JCU | COV | Log | -0.81 | -2.16 | 0.12 | 0.00 | 0.08 | 0.03 | -0.17 |
| EM | COV | Log | -0.88 | -2.02 | 0.12 | 0.00 | 0.09 | 0.03 | -0.06 |
| EQ | COV | Log | -0.98 | 0.19 | 0.14 | 0.00 | 0.12 | 0.02 | 1.11 |
| DTSG-RV | COV | Log | -0.88 | -1.42 | 0.16 | 0.00 | 0.12 | 0.04 | 0.17 |
| UOM | COV | Log | -1.00 | 2.20 | 0.18 | 0.00 | 0.16 | 0.02 | 2.27 |
| Baseline | COV | Nat | -0.53 | 9.02 | 31.72 | 0.00 | 15.33 | 16.39 | 5.56 |
| DSTG-tft | COV | Nat | -0.83 | 9.75 | 45.58 | 0.00 | 34.72 | 10.86 | 5.82 |
| JCU | COV | Nat | -0.81 | 10.75 | 77.67 | 0.00 | 56.86 | 20.82 | 6.40 |
| EM | COV | Nat | -0.88 | 11.01 | 79.59 | 0.00 | 61.85 | 17.74 | 6.51 |
| EQ | COV | Nat | -0.98 | 13.87 | 94.35 | 0.00 | 83.04 | 11.31 | 7.76 |
| DTSG-RV | COV | Nat | -0.88 | 11.56 | 100.77 | 0.00 | 79.80 | 20.98 | 6.74 |
| UOM | COV | Nat | -1.00 | 16.55 | 115.27 | 0.00 | 104.57 | 10.70 | 8.96 |
| EQ | FLU | Log | -0.24 | -4.68 | 0.03 | 0.00 | 0.00 | 0.02 | -1.39 |
| JCU | FLU | Log | -0.62 | -4.10 | 0.05 | 0.00 | 0.03 | 0.02 | -1.14 |
| EM | FLU | Log | -0.30 | -3.41 | 0.05 | 0.00 | 0.01 | 0.04 | -0.59 |
| DTSG-RV | FLU | Log | -0.45 | -2.89 | 0.07 | 0.00 | 0.03 | 0.05 | -0.56 |
| DSTG-tft | FLU | Log | -0.95 | -0.74 | 0.10 | 0.00 | 0.08 | 0.01 | 0.42 |
| UOM | FLU | Log | 0.82 | -2.30 | 0.11 | 0.07 | 0.00 | 0.04 | -0.45 |
| Baseline | FLU | Log | 0.96 | 0.12 | 0.19 | 0.16 | 0.00 | 0.03 | 0.93 |
| EQ | FLU | Nat | -0.24 | 8.99 | 23.49 | 0.00 | 3.20 | 20.29 | 5.47 |
| JCU | FLU | Nat | -0.62 | 9.47 | 42.67 | 0.00 | 24.64 | 18.03 | 5.72 |
| EM | FLU | Nat | -0.30 | 10.32 | 45.53 | 0.00 | 10.73 | 34.80 | 6.27 |
| DTSG-RV | FLU | Nat | -0.45 | 10.55 | 63.16 | 0.00 | 23.90 | 39.26 | 6.30 |
| DSTG-tft | FLU | Nat | -0.95 | 13.12 | 86.67 | 0.00 | 75.97 | 10.70 | 7.28 |
| UOM | FLU | Nat | 0.82 | 11.65 | 113.39 | 72.47 | 0.00 | 40.92 | 6.43 |
| Baseline | FLU | Nat | 0.96 | 13.58 | 205.10 | 168.97 | 0.00 | 36.13 | 7.78 |
| EQ | RSV | Log | 0.30 | -4.57 | 0.03 | 0.00 | 0.00 | 0.02 | -1.40 |
| JCU | RSV | Log | 0.14 | -4.24 | 0.03 | 0.00 | 0.00 | 0.03 | -1.21 |
| EM | RSV | Log | 0.16 | -3.79 | 0.04 | 0.00 | 0.00 | 0.04 | -0.88 |
| DTSG-RV | RSV | Log | 0.17 | -3.41 | 0.04 | 0.00 | 0.00 | 0.04 | -0.83 |
| UOM | RSV | Log | -0.60 | -3.84 | 0.05 | 0.00 | 0.02 | 0.02 | -1.10 |
| DSTG-tft | RSV | Log | 0.94 | -1.61 | 0.10 | 0.08 | 0.00 | 0.02 | 0.02 |
| Baseline | RSV | Log | 0.85 | -1.78 | 0.14 | 0.10 | 0.00 | 0.03 | 0.06 |
| EQ | RSV | Nat | 0.30 | 6.24 | 5.89 | 1.02 | 0.00 | 4.87 | 3.96 |
| JCU | RSV | Nat | 0.14 | 6.54 | 6.19 | 0.38 | 0.00 | 5.81 | 4.15 |
| EM | RSV | Nat | 0.16 | 6.99 | 8.37 | 0.64 | 0.00 | 7.73 | 4.48 |
| DTSG-RV | RSV | Nat | 0.17 | 7.41 | 9.58 | 0.70 | 0.00 | 8.88 | 4.53 |
| UOM | RSV | Nat | -0.60 | 6.67 | 9.78 | 0.00 | 5.04 | 4.74 | 4.26 |
| DSTG-tft | RSV | Nat | 0.94 | 9.04 | 22.42 | 18.35 | 0.00 | 4.07 | 5.39 |
| Baseline | RSV | Nat | 0.85 | 9.14 | 32.82 | 23.61 | 0.00 | 9.20 | 5.41 |

Table 19: Forecast metrics for 2024-09-12 . Where EM is ensemble mixture EQ is ensemble quantile JCU is JCU-renewal UOM is UOM-seirode. Nat is for the natural scale. Op and Up are over and under prediction respectivly. Disp is dispersion and LS is the log score.

##### 3.3 Forecast horizon

| Model | Path | Scale | Week | Bias | Dss | Crps | Op | Up | Disp | LS |
|---|---|---|---|---|---|---|---|---|---|---|
| EM | COV | Log | 1.00 | -0.04 | -3.07 | 0.08 | 0.02 | 0.02 | 0.04 | -0.54 |
| JCU | COV | Log | 1.00 | 0.10 | -2.99 | 0.08 | 0.03 | 0.02 | 0.03 | -0.58 |
| EQ | COV | Log | 1.00 | 0.00 | -2.66 | 0.08 | 0.03 | 0.03 | 0.02 | -0.39 |
| DTSG-RV | COV | Log | 1.00 | -0.02 | -2.86 | 0.08 | 0.02 | 0.03 | 0.04 | -0.52 |
| Baseline | COV | Log | 1.00 | 0.32 | -2.62 | 0.09 | 0.05 | 0.01 | 0.03 | -0.38 |
| DSTG-tft | COV | Log | 1.00 | 0.05 | 3.77 | 0.10 | 0.06 | 0.03 | 0.01 | 4.30 |
| UOM | COV | Log | 1.00 | -0.14 | -0.50 | 0.14 | 0.06 | 0.05 | 0.02 | 1.17 |
| EM | COV | Log | 2.00 | -0.14 | -2.10 | 0.13 | 0.04 | 0.04 | 0.06 | 0.15 |
| EQ | COV | Log | 2.00 | -0.02 | -0.68 | 0.14 | 0.07 | 0.05 | 0.03 | 1.36 |
| JCU | COV | Log | 2.00 | 0.14 | -1.61 | 0.15 | 0.06 | 0.04 | 0.05 | 0.10 |
| DTSG-RV | COV | Log | 2.00 | -0.01 | -1.53 | 0.16 | 0.06 | 0.05 | 0.05 | 0.16 |
| Baseline | COV | Log | 2.00 | 0.46 | -1.05 | 0.16 | 0.11 | 0.01 | 0.04 | 0.41 |
| DSTG-tft | COV | Log | 2.00 | -0.35 | 18.97 | 0.19 | 0.07 | 0.10 | 0.01 | 7.98 |
| UOM | COV | Log | 2.00 | -0.18 | 1.59 | 0.22 | 0.13 | 0.06 | 0.03 | 2.88 |
| EM | COV | Log | 3.00 | -0.04 | -1.51 | 0.16 | 0.05 | 0.03 | 0.08 | 0.45 |
| EQ | COV | Log | 3.00 | 0.15 | -0.26 | 0.19 | 0.11 | 0.04 | 0.03 | 0.70 |
| JCU | COV | Log | 3.00 | 0.20 | -1.00 | 0.21 | 0.10 | 0.03 | 0.07 | 0.39 |
| DTSG-RV | COV | Log | 3.00 | 0.03 | -0.66 | 0.23 | 0.10 | 0.06 | 0.07 | 0.54 |
| DSTG-tft | COV | Log | 3.00 | -0.36 | 22.89 | 0.23 | 0.07 | 0.14 | 0.01 | 10.17 |
| Baseline | COV | Log | 3.00 | 0.62 | -0.04 | 0.24 | 0.18 | 0.01 | 0.05 | 0.88 |
| UOM | COV | Log | 3.00 | -0.19 | 3.68 | 0.31 | 0.21 | 0.06 | 0.03 | 5.07 |
| EM | COV | Log | 4.00 | -0.03 | -0.95 | 0.20 | 0.07 | 0.02 | 0.10 | 0.99 |
| DSTG-tft | COV | Log | 4.00 | -0.34 | 17.94 | 0.24 | 0.06 | 0.16 | 0.01 | 10.92 |
| EQ | COV | Log | 4.00 | 0.16 | 0.64 | 0.25 | 0.18 | 0.03 | 0.04 | 1.02 |
| DTSG-RV | COV | Log | 4.00 | 0.04 | -0.14 | 0.28 | 0.14 | 0.05 | 0.08 | 0.82 |
| JCU | COV | Log | 4.00 | 0.23 | -0.40 | 0.28 | 0.15 | 0.02 | 0.10 | 0.68 |
| Baseline | COV | Log | 4.00 | 0.77 | 0.62 | 0.30 | 0.24 | 0.00 | 0.05 | 1.18 |
| UOM | COV | Log | 4.00 | -0.22 | 5.28 | 0.42 | 0.31 | 0.07 | 0.04 | 4.24 |
| DTSG-RV | COV | Nat | 1.00 | -0.02 | 11.12 | 94.06 | 30.52 | 19.63 | 43.92 | 6.46 |
| JCU | COV | Nat | 1.00 | 0.10 | 11.00 | 95.21 | 40.81 | 16.36 | 38.04 | 6.40 |
| EM | COV | Nat | 1.00 | -0.04 | 10.91 | 95.36 | 35.00 | 15.89 | 44.47 | 6.42 |
| EQ | COV | Nat | 1.00 | 0.00 | 11.29 | 101.14 | 54.57 | 20.86 | 25.70 | 6.58 |
| Baseline | COV | Nat | 1.00 | 0.32 | 11.26 | 109.32 | 61.94 | 12.55 | 34.83 | 6.59 |
| DSTG-tft | COV | Nat | 1.00 | 0.05 | 16.26 | 124.36 | 75.79 | 33.30 | 15.28 | 9.94 |
| UOM | COV | Nat | 1.00 | -0.14 | 13.71 | 194.48 | 121.34 | 39.75 | 33.40 | 8.48 |
| EM | COV | Nat | 2.00 | -0.14 | 11.90 | 166.67 | 57.13 | 27.91 | 81.62 | 7.07 |
| EQ | COV | Nat | 2.00 | -0.02 | 13.16 | 179.61 | 107.81 | 36.05 | 35.76 | 7.97 |
| DTSG-RV | COV | Nat | 2.00 | -0.01 | 12.32 | 191.04 | 91.99 | 34.86 | 64.20 | 7.11 |
| JCU | COV | Nat | 2.00 | 0.14 | 12.38 | 197.04 | 103.43 | 29.62 | 63.98 | 7.07 |
| Baseline | COV | Nat | 2.00 | 0.46 | 12.65 | 207.77 | 132.86 | 23.56 | 51.36 | 7.35 |
| DSTG-tft | COV | Nat | 2.00 | -0.35 | 30.98 | 220.73 | 72.68 | 134.20 | 13.85 | 12.60 |
| UOM | COV | Nat | 2.00 | -0.18 | 14.76 | 380.63 | 288.44 | 44.18 | 48.01 | 9.17 |
| EM | COV | Nat | 3.00 | -0.04 | 12.51 | 241.61 | 84.54 | 24.03 | 133.04 | 7.28 |
| DSTG-tft | COV | Nat | 3.00 | -0.36 | 37.09 | 254.97 | 61.39 | 180.79 | 12.80 | 13.84 |
| EQ | COV | Nat | 3.00 | 0.15 | 13.31 | 285.73 | 201.94 | 31.70 | 52.09 | 7.54 |
| Baseline | COV | Nat | 3.00 | 0.62 | 13.48 | 298.12 | 210.65 | 20.72 | 66.75 | 7.77 |
| DTSG-RV | COV | Nat | 3.00 | 0.03 | 13.00 | 302.91 | 170.48 | 42.78 | 89.66 | 7.45 |
| JCU | COV | Nat | 3.00 | 0.20 | 12.87 | 321.94 | 190.72 | 24.37 | 106.84 | 7.31 |
| UOM | COV | Nat | 3.00 | -0.19 | 15.42 | 665.03 | 541.16 | 48.20 | 75.66 | 10.45 |
| DSTG-tft | COV | Nat | 4.00 | -0.34 | 36.29 | 253.47 | 49.56 | 191.53 | 12.37 | 14.44 |
| EM | COV | Nat | 4.00 | -0.03 | 13.09 | 350.90 | 136.59 | 19.77 | 194.54 | 7.59 |
| Baseline | COV | Nat | 4.00 | 0.77 | 14.00 | 375.48 | 284.19 | 8.16 | 83.14 | 8.04 |
| DTSG-RV | COV | Nat | 4.00 | 0.04 | 13.25 | 436.22 | 278.64 | 36.59 | 120.99 | 7.66 |
| EQ | COV | Nat | 4.00 | 0.16 | 13.44 | 461.56 | 364.30 | 22.26 | 75.00 | 7.77 |
| JCU | COV | Nat | 4.00 | 0.23 | 13.49 | 503.18 | 314.32 | 19.04 | 169.82 | 7.55 |
| UOM | COV | Nat | 4.00 | -0.22 | 15.74 | 1076.24 | 898.90 | 51.62 | 125.72 | 10.37 |

Table 20: Where EM is ensemble mixture EQ is ensemble quantile JCU is JCU-renewal UOM is UOM-seirode. Nat is for the natural scale. Op and Up are over and under prediction respectivly. Disp is dispersion and LS is the log score.

| Model | Path | Scale | Week | Bias | Dss | Crps | Op | Up | Disp | LS |
|---|---|---|---|---|---|---|---|---|---|---|
| DTSG-RV | FLU | Log | 1.00 | 0.01 | -2.71 | 0.09 | 0.02 | 0.02 | 0.04 | -0.45 |
| EM | FLU | Log | 1.00 | 0.23 | -2.62 | 0.09 | 0.04 | 0.01 | 0.05 | -0.35 |
| EQ | FLU | Log | 1.00 | 0.23 | -2.22 | 0.09 | 0.05 | 0.02 | 0.02 | -0.27 |
| Baseline | FLU | Log | 1.00 | 0.11 | -1.46 | 0.12 | 0.07 | 0.03 | 0.03 | 0.15 |
| UOM | FLU | Log | 1.00 | 0.58 | -0.32 | 0.14 | 0.10 | 0.01 | 0.03 | 0.81 |
| DSTG-tft | FLU | Log | 1.00 | 0.25 | 17.00 | 0.16 | 0.09 | 0.06 | 0.01 | 7.56 |
| JCU | FLU | Log | 1.00 | 0.01 | 18.37 | 0.19 | 0.05 | 0.12 | 0.02 | 2.66 |
| EQ | FLU | Log | 2.00 | 0.34 | -0.10 | 0.14 | 0.09 | 0.01 | 0.03 | 0.64 |
| EM | FLU | Log | 2.00 | 0.30 | -1.34 | 0.14 | 0.06 | 0.01 | 0.07 | 0.29 |
| DTSG-RV | FLU | Log | 2.00 | 0.12 | -1.39 | 0.16 | 0.07 | 0.04 | 0.06 | 0.24 |
| UOM | FLU | Log | 2.00 | 0.66 | 1.88 | 0.21 | 0.17 | 0.01 | 0.03 | 3.43 |
| DSTG-tft | FLU | Log | 2.00 | 0.16 | 47.88 | 0.24 | 0.12 | 0.11 | 0.01 | 9.57 |
| Baseline | FLU | Log | 2.00 | 0.07 | 1.17 | 0.25 | 0.14 | 0.06 | 0.05 | 2.10 |
| JCU | FLU | Log | 2.00 | 0.15 | 18.32 | 0.29 | 0.10 | 0.16 | 0.03 | 3.69 |
| EQ | FLU | Log | 3.00 | 0.43 | 4.16 | 0.19 | 0.14 | 0.01 | 0.04 | 2.82 |
| EM | FLU | Log | 3.00 | 0.31 | 2.51 | 0.20 | 0.09 | 0.01 | 0.10 | 1.48 |
| DTSG-RV | FLU | Log | 3.00 | 0.16 | -0.50 | 0.24 | 0.11 | 0.04 | 0.09 | 0.68 |
| UOM | FLU | Log | 3.00 | 0.65 | 3.55 | 0.29 | 0.25 | 0.01 | 0.04 | 5.83 |
| Baseline | FLU | Log | 3.00 | 0.06 | 2.91 | 0.35 | 0.20 | 0.09 | 0.06 | 3.31 |
| JCU | FLU | Log | 3.00 | 0.20 | 13.16 | 0.36 | 0.14 | 0.18 | 0.04 | 3.78 |
| DSTG-tft | FLU | Log | 3.00 | 0.09 | 91.32 | 0.37 | 0.15 | 0.21 | 0.01 | 9.49 |
| EQ | FLU | Log | 4.00 | 0.52 | 7.56 | 0.25 | 0.19 | 0.00 | 0.05 | 3.76 |
| EM | FLU | Log | 4.00 | 0.31 | 5.34 | 0.25 | 0.11 | 0.00 | 0.13 | 1.63 |
| DTSG-RV | FLU | Log | 4.00 | 0.23 | 0.05 | 0.30 | 0.17 | 0.03 | 0.11 | 1.06 |
| UOM | FLU | Log | 4.00 | 0.65 | 4.57 | 0.39 | 0.34 | 0.01 | 0.04 | 6.52 |
| Baseline | FLU | Log | 4.00 | 0.06 | 3.70 | 0.43 | 0.25 | 0.11 | 0.06 | 4.41 |
| JCU | FLU | Log | 4.00 | 0.29 | 11.75 | 0.45 | 0.18 | 0.21 | 0.05 | 3.60 |
| DSTG-tft | FLU | Log | 4.00 | -0.09 | 165.79 | 0.57 | 0.18 | 0.37 | 0.01 | 11.98 |
| DTSG-RV | FLU | Nat | 1.00 | 0.01 | 12.84 | 230.91 | 67.19 | 50.73 | 112.98 | 7.32 |
| EQ | FLU | Nat | 1.00 | 0.23 | 13.19 | 244.31 | 141.21 | 39.31 | 63.79 | 7.51 |
| EM | FLU | Nat | 1.00 | 0.23 | 12.98 | 246.96 | 105.67 | 23.04 | 118.26 | 7.44 |
| Baseline | FLU | Nat | 1.00 | 0.11 | 13.79 | 306.64 | 146.90 | 71.54 | 88.19 | 7.92 |
| JCU | FLU | Nat | 1.00 | 0.01 | 73.89 | 360.69 | 153.21 | 154.15 | 53.33 | 9.66 |
| UOM | FLU | Nat | 1.00 | 0.58 | 14.70 | 398.16 | 286.01 | 37.47 | 74.68 | 8.49 |
| DSTG-tft | FLU | Nat | 1.00 | 0.25 | 32.97 | 478.03 | 267.61 | 179.22 | 31.21 | 12.25 |
| EQ | FLU | Nat | 2.00 | 0.34 | 14.95 | 389.19 | 250.58 | 52.08 | 86.53 | 8.44 |
| EM | FLU | Nat | 2.00 | 0.30 | 14.29 | 397.18 | 177.35 | 32.95 | 186.89 | 8.10 |
| DTSG-RV | FLU | Nat | 2.00 | 0.12 | 14.25 | 478.72 | 179.30 | 116.45 | 182.97 | 8.05 |
| Baseline | FLU | Nat | 2.00 | 0.07 | 15.68 | 612.32 | 323.40 | 157.01 | 131.92 | 9.33 |
| JCU | FLU | Nat | 2.00 | 0.15 | 90.52 | 619.36 | 302.77 | 230.95 | 85.65 | 10.66 |
| UOM | FLU | Nat | 2.00 | 0.66 | 16.12 | 646.36 | 515.48 | 34.10 | 96.78 | 10.24 |
| DSTG-tft | FLU | Nat | 2.00 | 0.16 | 72.01 | 687.99 | 324.28 | 333.04 | 30.68 | 13.65 |
| EQ | FLU | Nat | 3.00 | 0.43 | 17.67 | 520.25 | 358.74 | 37.40 | 124.11 | 9.69 |
| EM | FLU | Nat | 3.00 | 0.31 | 17.05 | 549.71 | 235.59 | 24.75 | 289.37 | 8.91 |
| DTSG-RV | FLU | Nat | 3.00 | 0.16 | 15.13 | 703.20 | 297.44 | 133.19 | 272.57 | 8.50 |
| JCU | FLU | Nat | 3.00 | 0.20 | 78.50 | 818.27 | 454.39 | 229.45 | 134.43 | 10.36 |
| Baseline | FLU | Nat | 3.00 | 0.06 | 16.73 | 884.38 | 475.54 | 235.07 | 173.77 | 10.08 |
| DSTG-tft | FLU | Nat | 3.00 | 0.09 | 192.41 | 938.89 | 350.37 | 556.73 | 31.80 | 13.61 |
| UOM | FLU | Nat | 3.00 | 0.65 | 16.84 | 1048.54 | 881.96 | 30.58 | 136.00 | 11.70 |
| EQ | FLU | Nat | 4.00 | 0.52 | 19.12 | 639.29 | 450.99 | 8.49 | 179.81 | 10.11 |
| EM | FLU | Nat | 4.00 | 0.31 | 18.60 | 689.47 | 250.53 | 8.78 | 430.16 | 9.09 |
| DTSG-RV | FLU | Nat | 4.00 | 0.23 | 15.68 | 872.76 | 400.75 | 92.72 | 379.29 | 8.78 |
| JCU | FLU | Nat | 4.00 | 0.29 | 104.16 | 1076.32 | 622.26 | 251.07 | 202.99 | 10.31 |
| Baseline | FLU | Nat | 4.00 | 0.06 | 17.09 | 1107.30 | 595.69 | 298.23 | 213.38 | 10.55 |
| DSTG-tft | FLU | Nat | 4.00 | -0.09 | 614.18 | 1244.25 | 367.39 | 845.82 | 31.03 | 14.96 |
| UOM | FLU | Nat | 4.00 | 0.65 | 16.93 | 1659.04 | 1436.64 | 19.08 | 203.32 | 12.10 |

Table 21: Where EM is ensemble mixture EQ is ensemble quantile JCU is JCU-renewal UOM is UOM-seirode. Nat is for the natural scale. Op and Up are over and under prediction respectivly. Disp is dispersion and LS is the log score.

| Model | Path | Scale | Week | Bias | Dss | Crps | Op | Up | Disp | LS |
|---|---|---|---|---|---|---|---|---|---|---|
| EQ | RSV | Log | 1.00 | 0.03 | -3.29 | 0.07 | 0.03 | 0.02 | 0.02 | -0.75 |
| EM | RSV | Log | 1.00 | -0.02 | -3.26 | 0.07 | 0.02 | 0.01 | 0.04 | -0.52 |
| DTSG-RV | RSV | Log | 1.00 | 0.35 | -2.62 | 0.08 | 0.05 | 0.00 | 0.03 | -0.12 |
| UOM | RSV | Log | 1.00 | 0.04 | -1.79 | 0.10 | 0.05 | 0.02 | 0.03 | 0.27 |
| JCU | RSV | Log | 1.00 | 0.28 | 2.42 | 0.12 | 0.08 | 0.01 | 0.02 | 2.97 |
| DSTG-tft | RSV | Log | 1.00 | -0.60 | 4.22 | 0.13 | 0.01 | 0.10 | 0.02 | 5.05 |
| Baseline | RSV | Log | 1.00 | 0.67 | -1.99 | 0.13 | 0.09 | 0.00 | 0.04 | -0.10 |
| EQ | RSV | Log | 2.00 | 0.00 | -2.17 | 0.10 | 0.06 | 0.02 | 0.03 | -0.09 |
| EM | RSV | Log | 2.00 | -0.05 | -2.52 | 0.11 | 0.04 | 0.02 | 0.05 | 0.10 |
| DTSG-RV | RSV | Log | 2.00 | 0.41 | -1.19 | 0.13 | 0.09 | 0.00 | 0.04 | 0.90 |
| UOM | RSV | Log | 2.00 | 0.02 | -0.02 | 0.14 | 0.08 | 0.03 | 0.03 | 1.70 |
| JCU | RSV | Log | 2.00 | 0.23 | 4.94 | 0.18 | 0.14 | 0.01 | 0.03 | 5.09 |
| DSTG-tft | RSV | Log | 2.00 | -0.73 | 11.55 | 0.18 | 0.00 | 0.16 | 0.02 | 7.34 |
| Baseline | RSV | Log | 2.00 | 0.76 | 0.01 | 0.27 | 0.21 | 0.00 | 0.06 | 0.92 |
| EM | RSV | Log | 3.00 | -0.02 | -2.02 | 0.15 | 0.06 | 0.03 | 0.06 | 0.72 |
| EQ | RSV | Log | 3.00 | 0.07 | -0.71 | 0.16 | 0.10 | 0.03 | 0.03 | 0.34 |
| UOM | RSV | Log | 3.00 | 0.03 | 1.60 | 0.20 | 0.13 | 0.04 | 0.03 | 2.65 |
| DSTG-tft | RSV | Log | 3.00 | -0.63 | 11.71 | 0.20 | 0.01 | 0.17 | 0.02 | 8.56 |
| DTSG-RV | RSV | Log | 3.00 | 0.43 | 1.91 | 0.20 | 0.16 | 0.00 | 0.04 | 1.83 |
| JCU | RSV | Log | 3.00 | 0.25 | 6.95 | 0.28 | 0.22 | 0.03 | 0.03 | 6.34 |
| Baseline | RSV | Log | 3.00 | 0.82 | 1.55 | 0.41 | 0.34 | 0.00 | 0.07 | 1.77 |
| DSTG-tft | RSV | Log | 4.00 | -0.64 | 9.24 | 0.18 | 0.01 | 0.15 | 0.02 | 6.53 |
| EM | RSV | Log | 4.00 | -0.00 | -1.19 | 0.20 | 0.09 | 0.03 | 0.07 | 1.12 |
| EQ | RSV | Log | 4.00 | 0.16 | 1.58 | 0.24 | 0.17 | 0.04 | 0.03 | 2.01 |
| UOM | RSV | Log | 4.00 | 0.08 | 3.03 | 0.26 | 0.18 | 0.04 | 0.03 | 3.42 |
| DTSG-RV | RSV | Log | 4.00 | 0.48 | 4.10 | 0.30 | 0.24 | 0.01 | 0.05 | 3.47 |
| JCU | RSV | Log | 4.00 | 0.28 | 8.67 | 0.40 | 0.32 | 0.04 | 0.04 | 7.08 |
| Baseline | RSV | Log | 4.00 | 0.88 | 2.96 | 0.57 | 0.49 | 0.00 | 0.08 | 2.49 |
| EQ | RSV | Nat | 1.00 | 0.03 | 9.85 | 69.05 | 38.19 | 8.18 | 22.68 | 5.82 |
| EM | RSV | Nat | 1.00 | -0.02 | 9.95 | 76.65 | 29.50 | 6.18 | 40.97 | 6.04 |
| DTSG-RV | RSV | Nat | 1.00 | 0.35 | 10.37 | 107.00 | 77.14 | 0.98 | 28.89 | 6.24 |
| UOM | RSV | Nat | 1.00 | 0.04 | 11.08 | 110.72 | 77.71 | 7.64 | 25.37 | 6.57 |
| Baseline | RSV | Nat | 1.00 | 0.67 | 11.41 | 119.27 | 72.63 | 0.61 | 46.02 | 6.50 |
| DSTG-tft | RSV | Nat | 1.00 | -0.60 | 20.01 | 132.26 | 3.17 | 118.22 | 10.87 | 9.74 |
| JCU | RSV | Nat | 1.00 | 0.28 | 13.92 | 148.68 | 124.34 | 5.08 | 19.26 | 8.43 |
| EQ | RSV | Nat | 2.00 | 0.00 | 10.80 | 119.46 | 81.99 | 9.83 | 27.65 | 6.49 |
| EM | RSV | Nat | 2.00 | -0.05 | 10.71 | 124.66 | 59.71 | 7.61 | 57.34 | 6.52 |
| UOM | RSV | Nat | 2.00 | 0.02 | 12.25 | 173.71 | 135.24 | 9.21 | 29.26 | 7.50 |
| DSTG-tft | RSV | Nat | 2.00 | -0.73 | 31.82 | 180.61 | 1.93 | 168.70 | 9.98 | 11.60 |
| DTSG-RV | RSV | Nat | 2.00 | 0.41 | 11.32 | 199.99 | 158.00 | 0.74 | 41.24 | 7.01 |
| Baseline | RSV | Nat | 2.00 | 0.76 | 13.04 | 250.22 | 176.39 | 1.90 | 71.92 | 7.47 |
| JCU | RSV | Nat | 2.00 | 0.23 | 14.91 | 251.35 | 218.14 | 5.34 | 27.86 | 9.68 |
| DSTG-tft | RSV | Nat | 3.00 | -0.63 | 31.04 | 180.07 | 3.81 | 166.53 | 9.74 | 12.15 |
| EM | RSV | Nat | 3.00 | -0.02 | 11.22 | 185.04 | 100.25 | 8.75 | 76.05 | 6.83 |
| EQ | RSV | Nat | 3.00 | 0.07 | 11.94 | 198.62 | 153.70 | 10.24 | 34.68 | 6.81 |
| UOM | RSV | Nat | 3.00 | 0.03 | 13.33 | 253.62 | 207.11 | 11.82 | 34.69 | 8.21 |
| DTSG-RV | RSV | Nat | 3.00 | 0.43 | 12.77 | 337.34 | 279.43 | 2.21 | 55.71 | 7.57 |
| Baseline | RSV | Nat | 3.00 | 0.82 | 14.05 | 395.76 | 294.56 | 1.97 | 99.22 | 8.21 |
| JCU | RSV | Nat | 3.00 | 0.25 | 15.54 | 403.19 | 351.93 | 9.55 | 41.71 | 10.43 |
| DSTG-tft | RSV | Nat | 4.00 | -0.64 | 27.57 | 149.38 | 6.30 | 133.69 | 9.39 | 11.15 |
| EM | RSV | Nat | 4.00 | -0.00 | 12.24 | 259.74 | 149.27 | 10.10 | 100.37 | 7.16 |
| EQ | RSV | Nat | 4.00 | 0.16 | 13.64 | 313.65 | 259.55 | 11.22 | 42.88 | 8.08 |
| UOM | RSV | Nat | 4.00 | 0.08 | 14.08 | 348.09 | 293.47 | 13.27 | 41.35 | 8.89 |
| DTSG-RV | RSV | Nat | 4.00 | 0.48 | 13.54 | 518.83 | 442.23 | 2.21 | 74.39 | 8.48 |
| Baseline | RSV | Nat | 4.00 | 0.88 | 14.78 | 549.68 | 418.95 | 0.49 | 130.25 | 8.80 |
| JCU | RSV | Nat | 4.00 | 0.28 | 15.81 | 609.44 | 536.87 | 11.49 | 61.08 | 11.10 |

Table 22: Where EM is ensemble mixture EQ is ensemble quantile JCU is JCU-renewal UOM is UOM-seirode. Nat is for the natural scale. Op and Up are over and under prediction respectivly. Disp is dispersion and LS is the log score.

##### 3.4 Overall

| Model | Path | Scale | Bias | Dss | Crps | Op | Up | Disp | LS |
|---|---|---|---|---|---|---|---|---|---|
| EM | COV | Log | -0.06 | -1.96 | 0.14 | 0.04 | 0.03 | 0.07 | 0.23 |
| EQ | COV | Log | 0.07 | -0.81 | 0.16 | 0.09 | 0.04 | 0.03 | 0.65 |
| JCU | COV | Log | 0.17 | -1.56 | 0.18 | 0.08 | 0.03 | 0.06 | 0.12 |
| DTSG-RV | COV | Log | 0.01 | -1.36 | 0.18 | 0.08 | 0.05 | 0.06 | 0.22 |
| DSTG-tft | COV | Log | -0.24 | 15.56 | 0.19 | 0.07 | 0.11 | 0.01 | 8.19 |
| Baseline | COV | Log | 0.53 | -0.85 | 0.19 | 0.14 | 0.01 | 0.04 | 0.48 |
| UOM | COV | Log | -0.18 | 2.37 | 0.26 | 0.17 | 0.06 | 0.03 | 3.26 |
| EM | COV | Nat | -0.06 | 12.05 | 207.95 | 76.07 | 21.85 | 110.03 | 7.07 |
| DSTG-tft | COV | Nat | -0.24 | 29.68 | 210.37 | 65.50 | 131.23 | 13.64 | 12.60 |
| Baseline | COV | Nat | 0.53 | 12.78 | 241.33 | 167.09 | 16.36 | 57.88 | 7.40 |
| DTSG-RV | COV | Nat | 0.01 | 12.37 | 247.93 | 137.03 | 33.04 | 77.86 | 7.14 |
| EQ | COV | Nat | 0.07 | 12.76 | 248.99 | 175.24 | 27.72 | 46.03 | 7.44 |
| JCU | COV | Nat | 0.17 | 12.38 | 269.71 | 155.84 | 22.33 | 91.54 | 7.06 |
| UOM | COV | Nat | -0.18 | 14.86 | 558.17 | 443.99 | 45.65 | 68.52 | 9.57 |
| EQ | FLU | Log | 0.37 | 2.12 | 0.16 | 0.12 | 0.01 | 0.03 | 1.64 |
| EM | FLU | Log | 0.29 | 0.78 | 0.17 | 0.07 | 0.01 | 0.08 | 0.72 |
| DTSG-RV | FLU | Log | 0.12 | -1.20 | 0.19 | 0.09 | 0.03 | 0.07 | 0.35 |
| UOM | FLU | Log | 0.63 | 2.30 | 0.25 | 0.21 | 0.01 | 0.03 | 4.01 |
| Baseline | FLU | Log | 0.08 | 1.45 | 0.28 | 0.16 | 0.07 | 0.05 | 2.39 |
| JCU | FLU | Log | 0.16 | 15.58 | 0.32 | 0.12 | 0.17 | 0.03 | 3.41 |
| DSTG-tft | FLU | Log | 0.11 | 77.00 | 0.33 | 0.13 | 0.18 | 0.01 | 9.56 |
| EQ | FLU | Nat | 0.37 | 16.10 | 439.37 | 293.37 | 35.04 | 110.96 | 8.88 |
| EM | FLU | Nat | 0.29 | 15.60 | 460.83 | 188.95 | 22.72 | 249.15 | 8.34 |
| DTSG-RV | FLU | Nat | 0.12 | 14.41 | 556.04 | 228.18 | 97.25 | 230.61 | 8.13 |
| JCU | FLU | Nat | 0.16 | 86.20 | 701.91 | 372.03 | 214.34 | 115.54 | 10.24 |
| Baseline | FLU | Nat | 0.08 | 15.74 | 708.56 | 374.68 | 185.05 | 148.83 | 9.41 |
| DSTG-tft | FLU | Nat | 0.11 | 214.58 | 819.08 | 325.09 | 462.82 | 31.17 | 13.56 |
| UOM | FLU | Nat | 0.63 | 16.09 | 908.14 | 752.75 | 30.73 | 124.66 | 10.54 |
| EM | RSV | Log | -0.03 | -2.29 | 0.13 | 0.05 | 0.02 | 0.05 | 0.32 |
| EQ | RSV | Log | 0.06 | -1.26 | 0.14 | 0.09 | 0.03 | 0.03 | 0.32 |
| UOM | RSV | Log | 0.04 | 0.59 | 0.17 | 0.11 | 0.03 | 0.03 | 1.94 |
| DSTG-tft | RSV | Log | -0.65 | 9.07 | 0.17 | 0.01 | 0.15 | 0.02 | 6.83 |
| DTSG-RV | RSV | Log | 0.41 | 0.38 | 0.17 | 0.13 | 0.00 | 0.04 | 1.44 |
| JCU | RSV | Log | 0.26 | 5.60 | 0.24 | 0.19 | 0.02 | 0.03 | 5.27 |
| Baseline | RSV | Log | 0.78 | 0.52 | 0.33 | 0.27 | 0.00 | 0.06 | 1.21 |
| EM | RSV | Nat | -0.03 | 10.98 | 157.40 | 81.98 | 8.07 | 67.35 | 6.61 |
| DSTG-tft | RSV | Nat | -0.65 | 27.45 | 160.21 | 3.72 | 146.47 | 10.03 | 11.12 |
| EQ | RSV | Nat | 0.06 | 11.47 | 169.70 | 128.39 | 9.80 | 31.52 | 6.75 |
| UOM | RSV | Nat | 0.04 | 12.61 | 215.88 | 173.24 | 10.35 | 32.29 | 7.74 |
| DTSG-RV | RSV | Nat | 0.41 | 11.92 | 280.99 | 230.51 | 1.50 | 48.98 | 7.27 |
| Baseline | RSV | Nat | 0.78 | 13.24 | 318.47 | 232.37 | 1.25 | 84.85 | 7.69 |
| JCU | RSV | Nat | 0.26 | 15.00 | 342.21 | 298.02 | 7.70 | 36.48 | 9.85 |

Table 23: Where EM is ensemble mixture EQ is ensemble quantile JCU is JCU-renewal UOM is UOM-seirode. Nat is for the natural scale. Op and Up are over and under prediction respectivly. Disp is dispersion and LS is the log score.

### References
